# Efficacy and Safety of a Booster Regimen of Ad26.COV2.S Vaccine against Covid-19

**DOI:** 10.1101/2022.01.28.22270043

**Authors:** Karin Hardt, An Vandebosch, Jerry Sadoff, Mathieu Le Gars, Carla Truyers, David Lowson, Ilse Van Dromme, Johan Vingerhoets, Tobias Kamphuis, Gert Scheper, Javier Ruiz-Guiñazú, Saul N. Faust, Christoph D. Spinner, Hanneke Schuitemaker, Johan Van Hoof, Macaya Douoguih, Frank Struyf, the ENSEMBLE2 Study Group

## Abstract

**Background:** Despite the availability of effective vaccines against coronavirus disease 2019 (Covid-19), the emergence of variant strains and breakthrough infections pose a challenging new reality. Booster vaccinations are needed to maintain vaccine-induced protection.

**Methods:** ENSEMBLE2 is an ongoing, randomized, double-blind, placebo-controlled, phase 3 pivotal trial including crossover vaccination after emergency authorization of Covid-19 vaccines. Adults aged ≥18 years were randomized to receive Ad26.COV2.S or placebo as a primary dose plus a booster dose at two months. The primary endpoint was vaccine efficacy against the first occurrence of molecularly-confirmed moderate to severe–critical Covid-19 with onset ≥14 days after booster vaccination in the per-protocol population. Key efficacy, safety, and immunogenicity endpoints were also assessed.

**Results:** The double-blind phase enrolled 31,300 participants, 14,492 of whom received 2 doses and were evaluable for efficacy (per-protocol set, Ad26.COV2.S n=7484; placebo n=7008). Baseline demographics and characteristics were balanced. Vaccine efficacy was 75.2% (adjusted 95% CI, 54.6-87.3) against moderate to severe–critical Covid-19 and was similar against symptomatic infection (75.6% [55.5-99.9]). Efficacy was consistent across participants with and without comorbidities, and reached 93.7% (58.5-99.9) in the US. Vaccine efficacy against severe–critical Covid-19 was 100% (32.6-100.0; 0 vs 8 cases). The booster vaccine induced robust humoral responses and exhibited an acceptable safety profile.

**Conclusions:** A homologous Ad26.COV2.S booster administered 2 months after primary single-dose vaccination in adults led to high vaccine efficacy, including against any symptomatic infection and SARS-CoV-2 variants prevalent during the study. (Funding: Janssen Research and Development and others; ENSEMBLE2 ClinicalTrials.gov number, NCT04614948.)

## INTRODUCTION

The emergence of variant strains and breakthrough infections^1^ requires the improvement and prolongation of vaccine-induced protection against severe acute respiratory syndrome coronavirus-2 (SARS-CoV-2) infection and coronavirus disease 2019 (Covid-19). The initial World Health Organization target profile recommended that Covid-19 vaccines should be highly efficacious as a single dose, with booster doses administered for long-term protection.^2^ The Ad26.COV2.S vaccine is a recombinant, replication-incompetent human adenovirus type 26 (Ad26) vector encoding a prefusion conformation-stabilized, full-length, membrane-bound SARS-CoV-2 spike protein.^3^ We report here the results of the double-blind portion of the phase 3 ENSEMBLE2 trial investigating the efficacy, safety, and immunogenicity of Ad26.COV2.S administered as primary vaccination plus a booster dose after a 56-day (2-month) interval.

## METHODS

### Trial Design and Oversight

ENSEMBLE2 is an ongoing, randomized, double-blind, placebo-controlled, phase 3 trial (including crossover vaccination after emergency authorization of Covid-19 vaccines) conducted in 10 countries: Belgium, Brazil, Colombia, France, Germany, The Philippines, South Africa, Spain, the United Kingdom, and the United States. Objectives and endpoints are in **Table S1**.

The sponsor was responsible for trial design, conduct, data analysis, and data interpretation. Trial investigators collected data and contributed to interpretation. The protocol and amendments were approved by ethics committees and institutional review boards per local regulations.

All relevant ethical guidelines have been followed, all necessary institutional review board (IRB) and/or ethics committee approvals have been obtained, all necessary patient/participant consent has been obtained, and the appropriate institutional forms archived. The COV3009 (ENSEMBLE2) study was reviewed and approved by local ethics committees and IRBs (Algemeen Ziekenhuis Sint-Jan, Brugge, Belgium; Comissão Nacional de Ética em Pesquisa, Brasília, Brazil; Instituto Nacional de Vigilancia de Medicamentos y Alimentos (Colombia), Bogota, Colombia; Comité de Protection des Personnes Ile de France III, Paris, France; Ethik-Kommission der Fakultaet f. Medizin der Technischen Universitaet Muenchen, Muenchen, Germany; Single Joint Research Ethics Board, Manila City, Philippines; Pharma Ethics, Pretoria, South Africa; Hospital Universitario La Paz, Madrid, Spain; UK NHS Research Ethics Service, Health Research Authority, London, United Kingdom; Copernicus Group IRB, Cary, North Carolina, United States.

All data were available to the authors, who vouch for data accuracy, completeness, and adherence to the study protocol. All participants provided written informed consent. This trial adheres to the International Conference for Harmonisation guidelines on Good Clinical Practice and Declaration of Helsinki principles. Sponsor-funded medical writers assisted with drafting the manuscript.

### Trial Participants

Participants were adults aged ≥18 years, healthy or with stable and well-controlled comorbidities, and without prior receipt of a Covid-19 vaccine (**Supplementary Methods**). After emergency use authorization (EUA) for some vaccines, participants could be unblinded to enable vaccination of placebo recipients outside of the study; once Ad26.COV2.S received EUA, placebo recipients without Covid-19 vaccination outside the study were offered open-label Ad26.COV2.S vaccination.

### Procedures

Participants randomized 1:1 via computer-generated randomly permuted blocks received 2 doses, referred to as a primary dose plus a booster dose of Ad26.COV2.S (each 5×10^10^ viral particles) or saline placebo as an intramuscular injection (0.5 mL) 56 days apart. Participants and sites were blinded to assignment until the unblinding visit (**Supplementary Methods)**.

Efficacy assessments were performed using centrally molecularly-confirmed Covid-19 cases identified by real-time reverse transcriptase polymerase chain reaction-based or other molecular diagnostic tests. Disease severity was assessed independently by a Clinical Severity Adjudication Committee (**Supplementary Methods**). Participants reported Covid-19 symptoms using the electronic Symptoms of Infection with Coronavirus-19 questionnaire.

### Efficacy Assessments

Unless stated otherwise, efficacy assessments are presented for the first occurrence of molecularly-confirmed Covid-19 with onset ≥14 days after booster vaccination in the per-protocol (PP) population. The primary endpoint was vaccine efficacy against moderate to severe–critical Covid-19. The primary analysis was triggered when ≥90% of participants were unblinded. Immunogenicity was evaluated by spike protein-specific binding antibodies using sera collected at various time points (immunogenicity subset, n=400).

### Safety Assessments

After each vaccination, a safety subset comprising approximately 6000 participants recorded solicited local and systemic adverse events (AEs) in an electronic diary for 7 days and unsolicited AEs for 28 days or until unblinding. In all participants, medically attended AEs were followed for 6 months after each vaccination; serious AEs (SAEs), AEs leading to study or vaccine discontinuation, and thrombosis with thrombocytopenia syndrome (TTS; an AE of special interest [as of protocol amendment 5]), were recorded throughout the study. AEs of clinical interest (not prespecified) were selected based on lists proposed by expert groups and regulatory authorities (**Supplementary Methods**).

### Statistical Analysis

The full analysis set (FAS) comprised all randomized participants who received ≥1 dose of trial vaccine or placebo. The per-protocol (PP) set included participants who received 2 doses of vaccine/placebo in the double-blind phase, were SARS-CoV-2 seronegative at day 1 and day 71 and had no major protocol violations possibly impacting efficacy. The risk set for vaccine efficacy evaluations excluded participants who had a Covid-19 case or discontinued before day 70. The per-protocol first dose (PPFD) set received ≥1 dose of vaccine/placebo in the double-blind phase, were seronegative at baseline, and had no major protocol deviations. Immunogenicity analyses were conducted in the immunogenicity subset, and safety analyses were conducted in the FAS and safety subset (**Supplementary Methods**).

Sample size was based on an assumption of vaccine efficacy of 65% against molecularly-confirmed moderate to severe–critical SARS-CoV-2 infection, a type 1 error rate of 0.025 to evaluate vaccine efficacy, and a 1:1 randomization ratio. In total, 104 events would provide approximately 90% power to reject the primary endpoint null hypothesis.

The primary endpoint null hypothesis was vaccine efficacy of Ad26.COV2.S of 30% or lower (tested at a 0.025 one-sided significance level). If the null hypothesis was rejected, confirmatory secondary endpoints were tested against a null hypothesis using a lower limit of efficacy <0%, employing a prespecified multiple testing strategy preserving the 0.025 family-wise error rate. All other endpoints or subgroup analyses were summarized descriptively with 95% confidence intervals (CI). Efficacy and associated CI calculations were performed using exact Poisson regression.^4^ Estimated cumulative incidence rates to evaluate time to first occurrence of Covid-19 and vaccine efficacy over time were assessed by Kaplan-Meier methods.

The frequency of serious AEs, AEs of special interest, and medically attended AEs were summarized descriptively for the FAS. The frequency and severity of solicited and unsolicited AEs were summarized descriptively in the safety subset.

## RESULTS

### Participants

Enrollment began November 16, 2020, and the primary analysis data cutoff was June 25, 2021 (before Delta became globally dominant and before the emergence of Omicron). Overall, 31,300 participants were randomized and vaccinated in the double-blind phase, of which 16,751 (53.5%) received both doses (Ad26.COV2.S, n=8655; placebo, n=8096) with 14,492 included in the PP population (**Figure 1**). Demographic and baseline characteristics were balanced between groups in the FAS and PP sets (**Table 1**,). In the PP set, median follow-up was 36.0 days post-booster; 4245 (29.3%) participants had ≥2 months follow-up post-booster. Most (92.1%) participants in the FAS were still in the study up to the data cutoff date. Overall, 2459 (7.9%) participants withdrew from the study: 5868 (18.7%) discontinued vaccination, including those who initiated prohibited medication (Ad26.COV2.S group, 284/2124 [13.4%]; placebo group, 1423/3744 [38.0%]). Once unblinded, more placebo recipients (3653/4680 [78.1%]) discontinued vaccination and received another vaccine outside the study than Ad26.COV2.S recipients (417/4267 [9.8%]). In the vaccine arm, 5 (<0.1%) participants withdrew from the study and 29 (1.4%) discontinued vaccination due to an AE. Unsolicited AEs leading to study or vaccine discontinuation in the safety subset are summarized in **Table S3**. Despite discontinuations, follow-up time in the double-blind phase between arms in the PP set and FAS was similar (**Figure S1**).

**Table 1.** Baseline Characteristics of the Trial Participants (Full Analysis Set*)

|  | **Ad26.COV2.S** | **Placebo** | **Total** |
| --- | --- | --- | --- |
| **Characteristic** | **(N=15,708)** | **(N=15,592)** | **(N=31,300)** |
| Age – no. (%) | 15,707 | 15,592 | 31,299 |
| 18-59 yr | 10,089 (64.2) | 9,978 (64.0) | 20,067 (64.1) |
| ≥60 yr | 5618 (35.8) | 5614 (36.0) | 11,232 (35.9) |
| Age – median<br>(range), yr | 53.0 (18-99) | 53 (18-95) | 53 (18-99) |
| Sex – no. (%) | 15,707 | 15,592 | 31,299 |
| Female | 7391 (47.1) | 7429 (47.6) | 14,820 (47.3) |
| Male | 8314 (52.9) | 8160 (52.3) | 16,474 (52.6) |
| Undifferentiated | 2 (<0.1) | 3 (<0.1) | 5 (<0.1) |
| Race<sup>†</sup> – no. (%) | 15,708 | 15,592 | 31,300 |
| American Indian or<br>Alaskan Native<sup>‡</sup> | 393 (2.5) | 396 (2.5) | 789 (2.5) |
| Asian | 1379 (8.8) | 1353 (8.7) | 2732 (8.7) |
| Black or African<br>American | 1309 (8.3) | 1245 (8.0) | 2554 (8.2) |
| Native Hawaiian or<br>other Pacific Islander | 33 (0.2) | 43 (0.3) | 76 (0.2) |
| White | 11,974 (76.2) | 11,933 (76.5) | 23,907 (76.4) |
| Multiracial | 225 (1.4) | 219 (1.4) | 444 (1.4) |
| Not reported,<br>unknown or missing | 395 (2.5) | 403 (2.6) | 798 (2.5) |
| Ethnicity<sup>†</sup> – no. (%) | 15,708 | 15,592 | 31,300 |
| Hispanic or Latino | 2827 (18.0) | 2806 (18.0) | 5633 (18.0) |
| Not Hispanic or<br>Latino | 12,430 (79.1) | 12,344 (79.2) | 24,774 (79.2) |
| Not reported,<br>unknown or missing | 451 (2.9) | 442 (2.8) | 893 (2.9) |
| Country/region – no. (%) | 15,707 | 15,592 | 31,299 |
| Europe | 6416 (40.8) | 6416 (41.1) | 12,832 (41.0) |
| Belgium | 1489 (9.5) | 1492 (9.6) | 2981 (9.5) |
| France | 356 (2.3) | 358 (2.3) | 714 (2.3) |
| Germany | 51 (0.3) | 49 (0.3) | 100 (0.3) |
| Spain | 1563 (10.0) | 1569 (10.1) | 3132 (10.0) |
| United Kingdom | 2957 (18.8) | 2948 (18.9) | 5905 (18.9) |
| Latin America | 1325 (8.4) | 1324 (8.5) | 2649 (8.5) |
| Brazil | 251 (1.6) | 249 (1.6) | 500 (1.6) |
| Colombia | 1074 (6.8) | 1075 (6.9) | 2149 (6.9) |
| Philippines | 784 (5.0) | 788 (5.1) | 1572 (5.0) |
| South Africa | 1037 (6.6) | 1035 (6.6) | 2072 (6.6) |
| United States | 6145 (39.1) | 6029 (38.7) | 12,174 (38.9) |
| SARS-CoV-2 serostatus –<br>no. (%) | 15,708 | 15,592 | 31,300 |
| Positive | 1757 (11.2) | 1721 (11.0) | 3478 (11.1) |
| Negative | 13,803 (87.9) | 13,759 (88.2) | 27,562 (88.1) |
| Missing | 148 (0.9) | 112 (0.7) | 260 (0.8) |
| Body-mass index<sup>§</sup> | 15,691 | 15,583 | 31,274 |
| Median | 26.5 | 26.6 | 26.6 |
| ≥30 – no. (%) | 4142 (26.4) | 4068 (26.1) | 8210 (26.3) |
| One or more comorbidity<br>at baseline – no. (%) | 6519 (41.5) | 6434 (41.3) | 12,953 (41.4) |

\*The full analysis set included all participants who were randomized and received at least one documented dose of Ad26.COV2.S vaccine or placebo, regardless of protocol deviations and serostatus at enrollment.
†Race and ethnicity were self-reported by participants.
‡Participants responding “Yes” to American Indian or Alaska Native in the Ad26.COV2.S group were from Colombia (n=326), the United States (n=38), Spain (n=24), Brazil (n=2), the United Kingdom, and the Philippines (n=1 each); in the placebo group, participants were from Colombia (n=321), the United States (n=39), Spain (n=28), the United Kingdom (n=4), Belgium, Brazil, and France (n=1 each).
§Body-mass index (BMI) is the weight in kilograms divided by the square of the height in meters. A BMI equal to or greater than 30 indicates obesity.
SARS-CoV-2, severe acute respiratory coronavirus 2.

**Figure 1.**
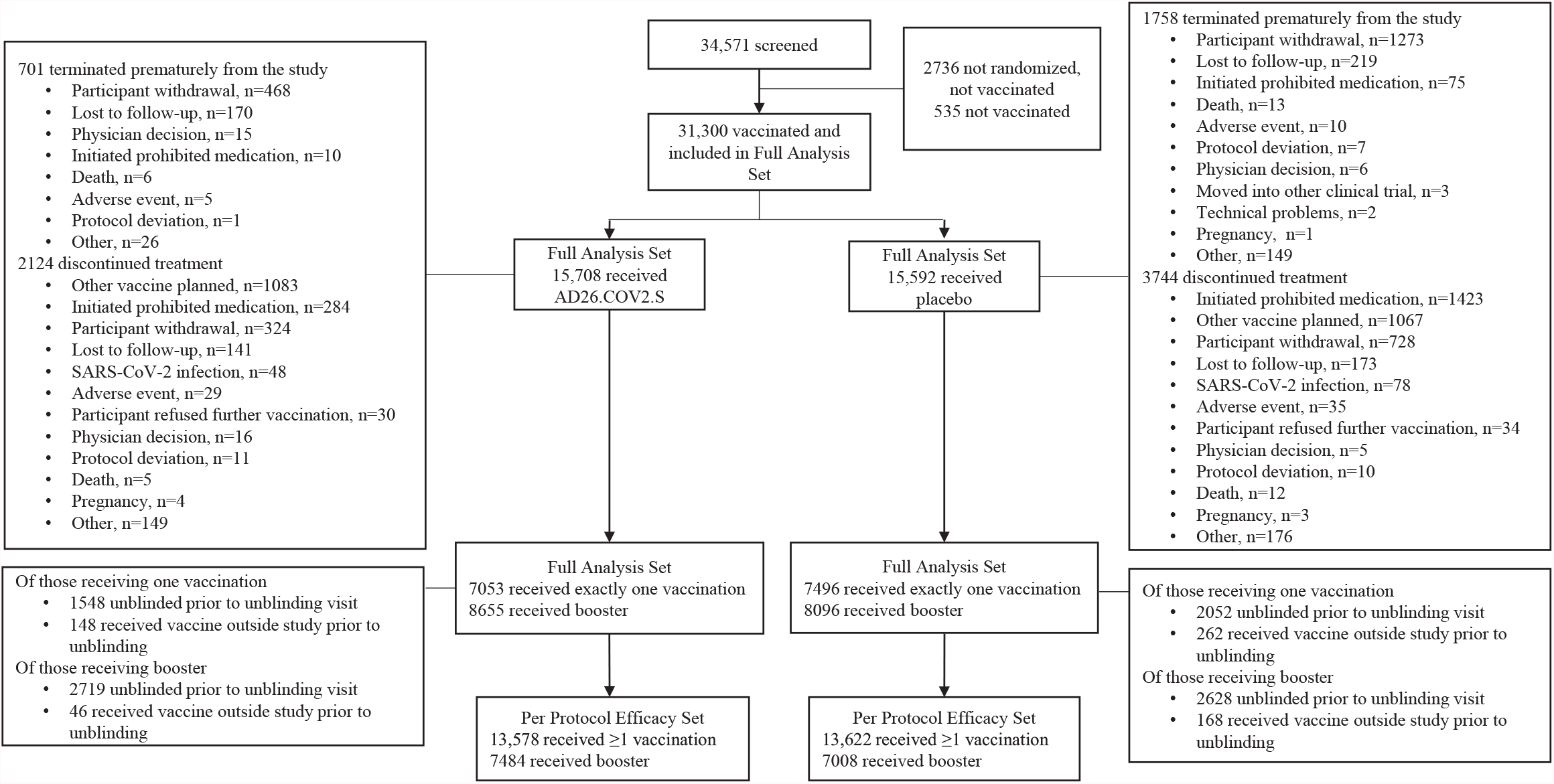
Participant Disposition. Data cutoff for analysis was June 25, 2021. The full analysis set (FAS) included all participants who were randomized and received ≥1 dose of trial vaccine or placebo, regardless of protocol deviations or serostatus at enrollment. The per-protocol set included participants in the FAS who received primary and booster doses of study vaccine and who were seronegative at the time of first vaccination and at day 71, and who had no other major protocol deviations. The safety subset included participants in the FAS who were monitored for solicited and unsolicited adverse events.

### Efficacy

At the primary analysis of the double-blind phase, 14 molecularly-confirmed moderate to severe–critical Covid-19 cases with onset ≥14 days after booster vaccination were reported in the Ad26.COV2.S group and 52 in the placebo group, indicating a vaccine efficacy of 75.2% (adjusted 95% confidence interval [CI], 54.6-87.3) (**Table 2**). Efficacy was similar against non-centrally-confirmed cases (**Table S4**). The cumulative incidence curves of molecularly-confirmed moderate to severe–critical Covid-19 cases with onset ≥1 day after primary vaccination separated after 14 days (**Figure 2A**); separation occurred shortly post-booster (**Figure S2**).

**Table 2.**
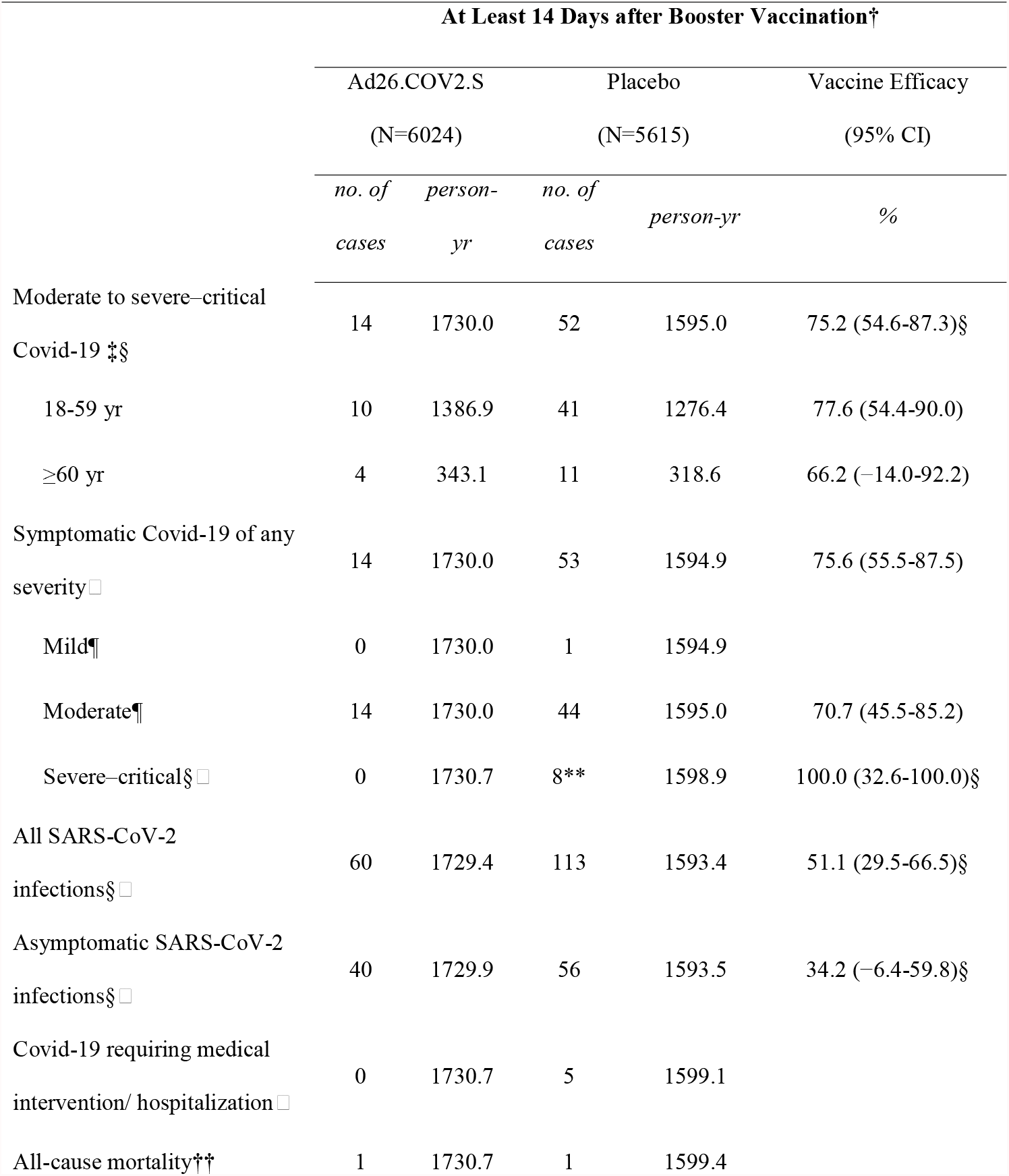
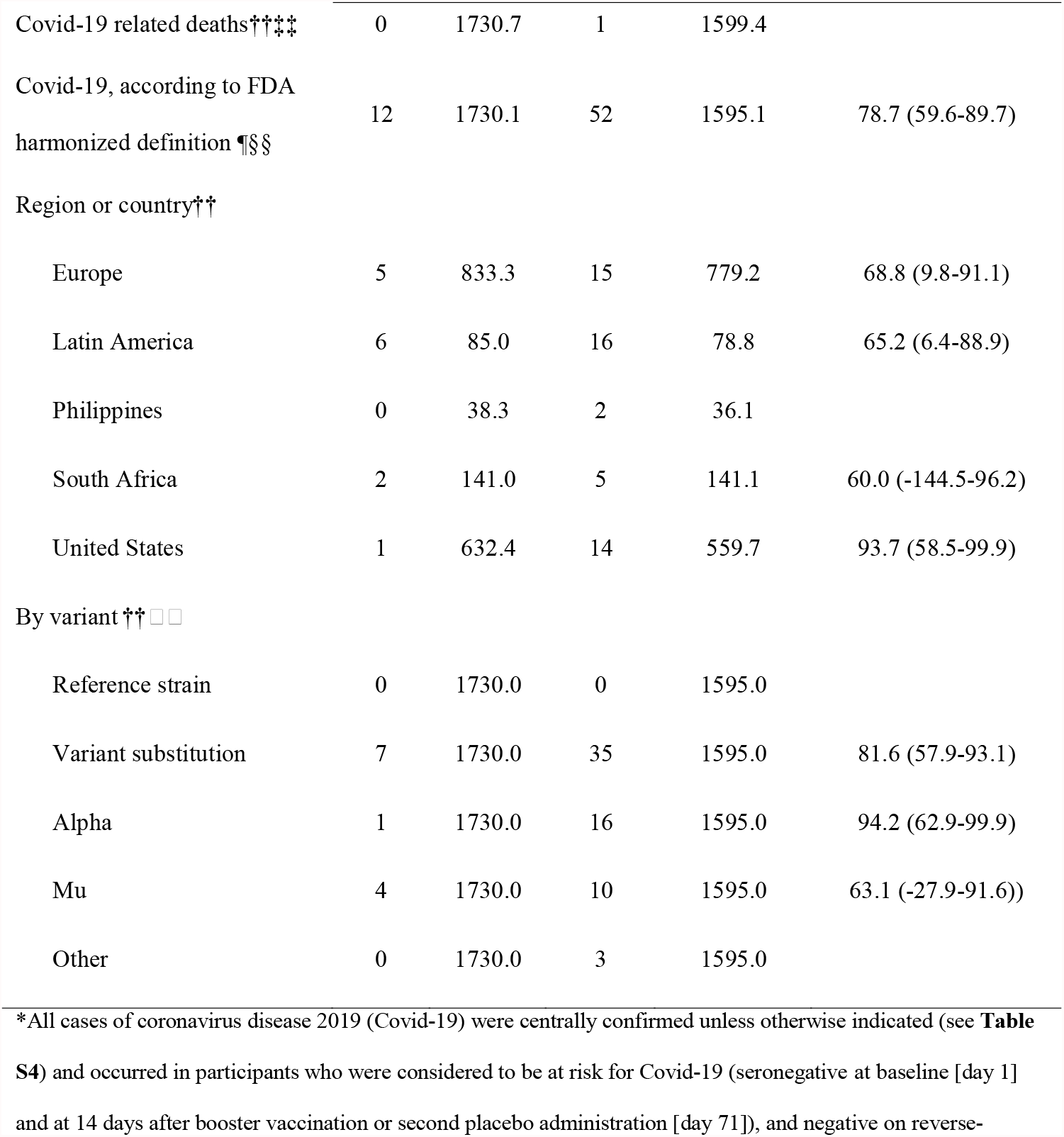
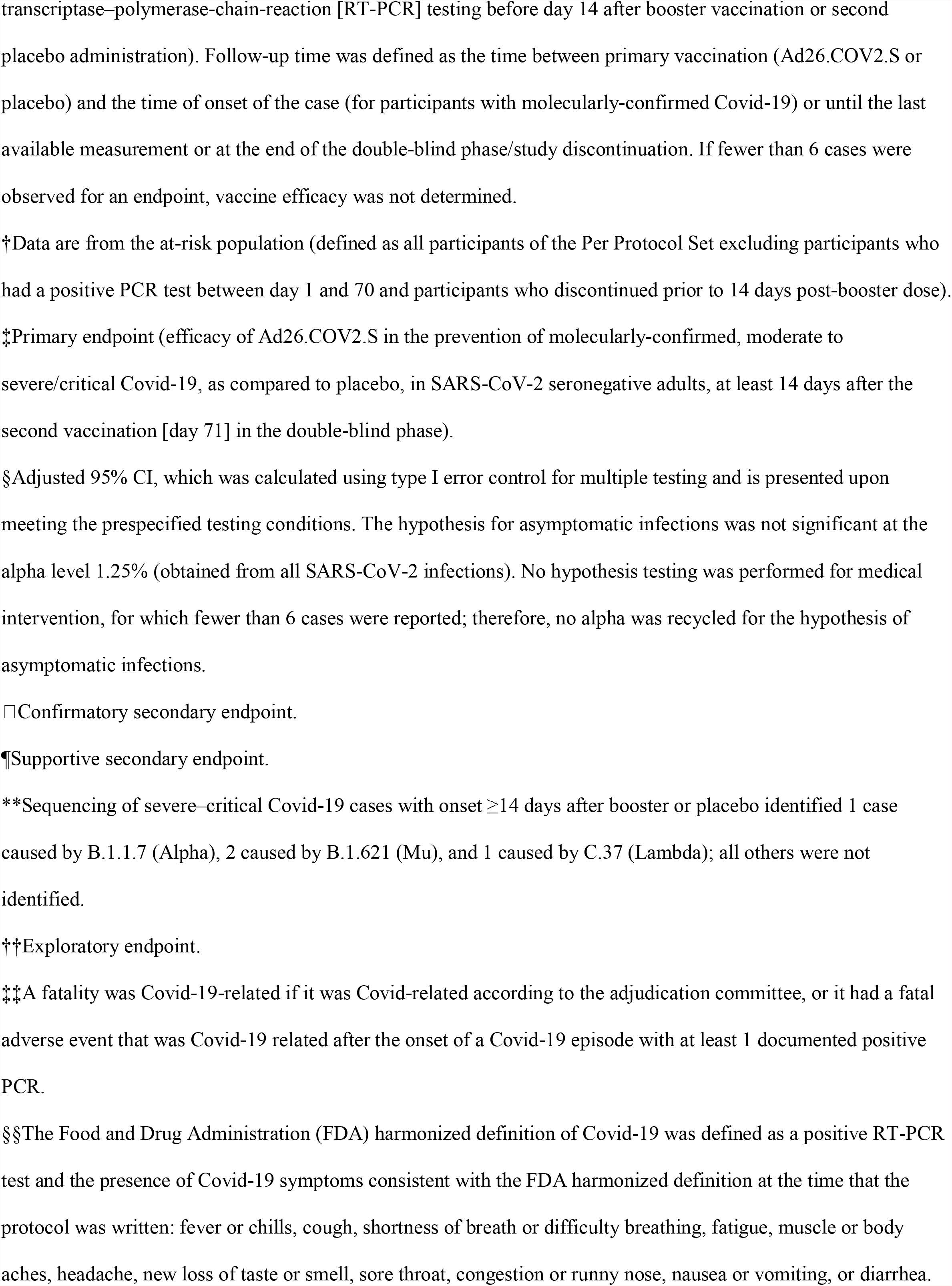
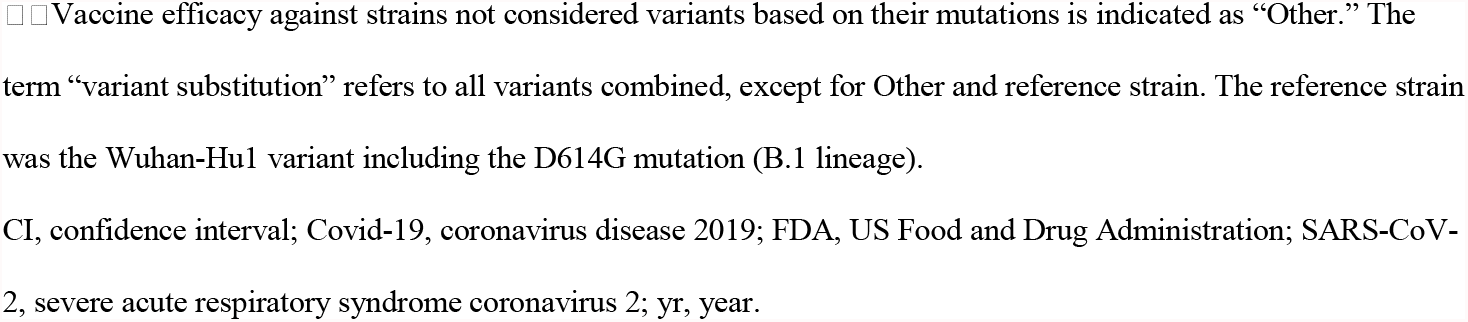
**Vaccine Efficacy against Molecularly-Confirmed Covid-19 with Onset at Least 14 Days after the Administration of a Booster Vaccine or Placebo (Per-Protocol Set)*\****

**Figure 2.**
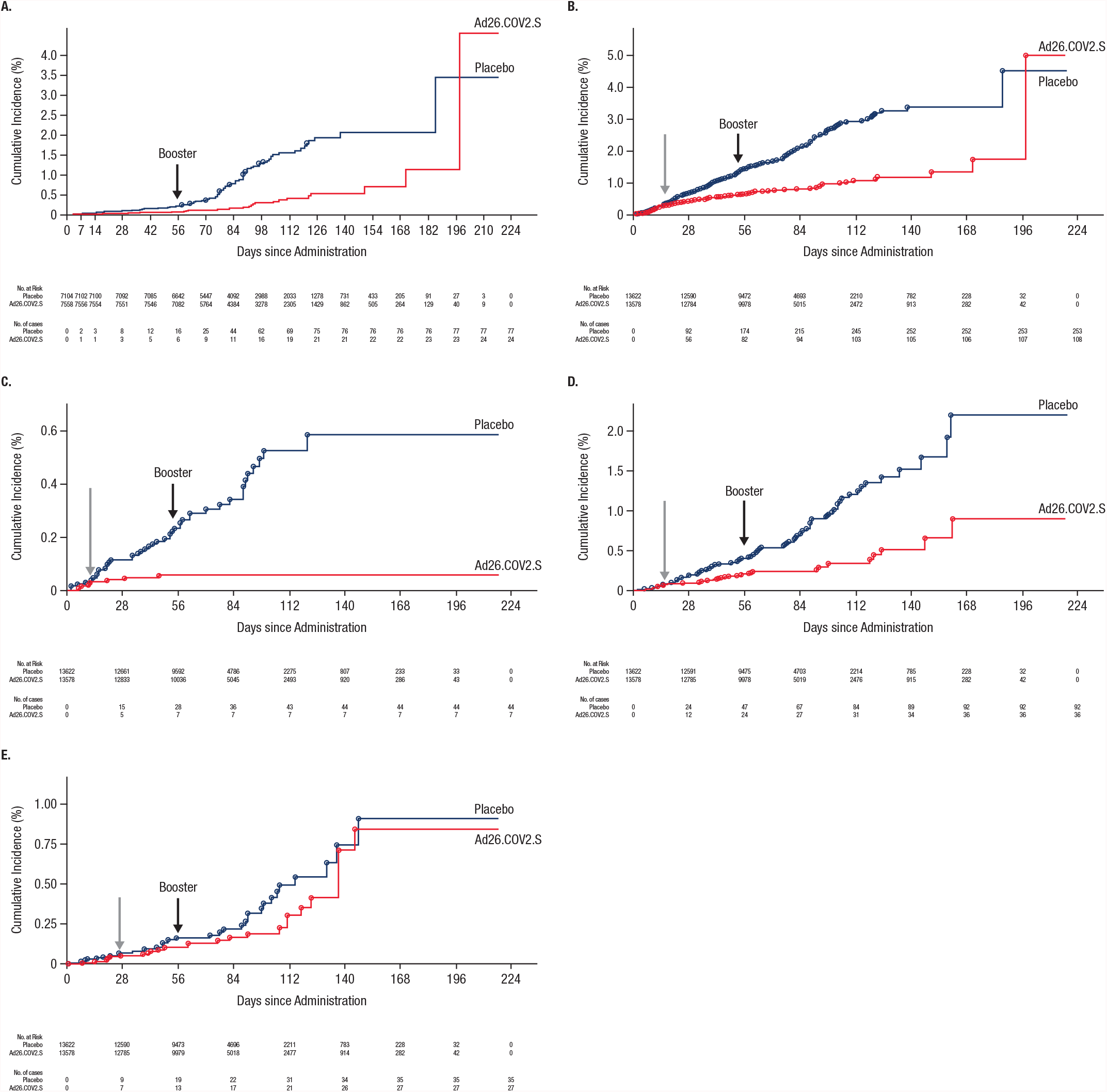
Cumulative Incidence of First Occurrence of Molecularly-Confirmed Moderate to Severe–Critical or Severe–Critical Covid-19 Cases with Onset at Least 1 Day After Vaccination (Per-Protocol and Per-Protocol First Dose Efficacy Set). The cumulative incidence of molecularly-confirmed moderate to severe–critical cases of Covid-19 with onset at least 1 day after the first vaccination in the per-protocol (PP) set.is shown in Panel A (circles indicate severe–critical Covid-19 cases in Panel A), and in the per-protocol first dose (PPFD) set in Panel B. Panel C shows the cumulative incidence of molecularly-confirmed severe–critical cases of Covid-19 with onset at least 1 day after the first vaccination in the PPFD set. Panels D and E show the cumulative incidence of molecularly-confirmed moderate to severe–critical cases of Covid-19 with onset at least 1 day after the first vaccination due, respectively, to Alpha (B.1.1.7) and Mu (B.1.621) in the PPFD set. The gray arrows in panels B–E indicate time points at which the Ad26.COV2.S and placebo curves begin to separate, and the black arrows in all panels indicate administration of the booster vaccine or placebo dose. Cases included for analysis were centrally confirmed cases in the PP and PPFD set among participants who were seronegative at baseline. The PPFD set includes participants who received at least one dose, regardless of their second dose, were baseline-SARS-CoV-2 seronegative and had no major protocol deviations impacting efficacy. Ad26 5e10 vp, Ad26.COV2.S given at a dose level of 5×10^10^ viral particles; CI, confidence interval; Covid-19, coronavirus disease 2019; vp, viral particles.

At the time of the analysis, sequencing data were available for 319/469 (68.0%) molecularly-confirmed infections in the double-blind phase (**Figure S3**). The reference sequence (Wuhan-Hu1 plus D614G) was present in 6.0% of sequenced strains. No moderate to severe–critical cases involving the reference strain were reported after booster vaccination. Overall efficacy against moderate to severe–critical Covid-19 for pooled variants differing from the reference strain (“variant substitutions” were variants of concern/interest, excluding “Other” and “reference”) was 81.6% (95% CI, 57.9-93.1) (**Figure S4**) with 94.2% [62.9-99.9] reported for Alpha (B.1.1.7) and 63.1% [-27.9-91.6]) for Mu (B.1.621) variants. Insufficient cases (<6) were available to analyze other variants, including Delta.

Eight severe–critical Covid-19 cases were reported, all in the placebo group, for vaccine efficacy of 100% (adjusted 95% CI, 32.6-100.0%) (**Table 2**). No cases of Covid-19 requiring medical intervention occurred in the Ad26.COV2.S group versus 5 cases in the placebo group. No Covid-19–related death was reported for any Ad26.COV2.S recipients and for 1 placebo recipient.

Consistent efficacy was observed for subgroups with sufficient numbers of cases (ages 18–59 and ≥60 years; participants with and without comorbidities) (**Figure S5**). Low numbers of participants in some subgroups resulted in wide CIs, possibly confounded by differential distribution of variants across regions. In the US, with the largest representation in the PP efficacy set (36.5%), efficacy against moderate to severe–critical Covid-19 was 93.7% (58.5-99.9).

Vaccine efficacy against all infections, including asymptomatic, was 51.1% (adjusted 95% CI, 29.5-66.5); overall efficacy against asymptomatic infection was 34.2% (adjusted 95% CI, -6.4-59.8), and efficacy against symptomatic infection reached 75.6% (95% CI, 55.5-87.5) (**Table 2**). Ad26.COV2.S recipients with breakthrough infections had fewer symptoms, lower symptom severity, and fewer cases lasting >28 days versus placebo recipients (**Figures S6-S7**).

Efficacy in the PPFD set (n=15,708) against moderate to severe–critical Covid-19 with onset from day 15-56 (representing those who received only 1 dose) was 67.0% (95% CI, 53.6-76.9) and efficacy against severe–critical Covid-19 was 86.6% (55.3-97.4; **Table S5, Figure 2B-C**). Efficacy against moderate to severe–critical Covid-19 caused by variants with onset from day 15-56 was 71.6% (43.2-86.9) for Alpha; and 43.9 (−43.4-79.6) for Mu (**Figure 2D-E**).

### Immunogenicity

In the immunogenicity subset, geometric mean increases in spike-specific binding antibody concentrations were 4.2- and 40.5-fold from baseline to day 29 and day 71, respectively (**Figure S8**). Following a single vaccination, response rates were 91.9% by day 29; after boosting, response rates reached 100% by day 71.

### Safety

The Ad26.COV2.S booster had an acceptable safety and reactogenicity profile. More AEs were reported in the vaccine group than placebo. The overall frequencies of local and systemic solicited AEs were similar following first and booster vaccinations (**Figure 3, Table S6**), with no increase in reactogenicity, and lower frequency in older adults versus younger adults (**Figure S9**).

**Figure 3.**
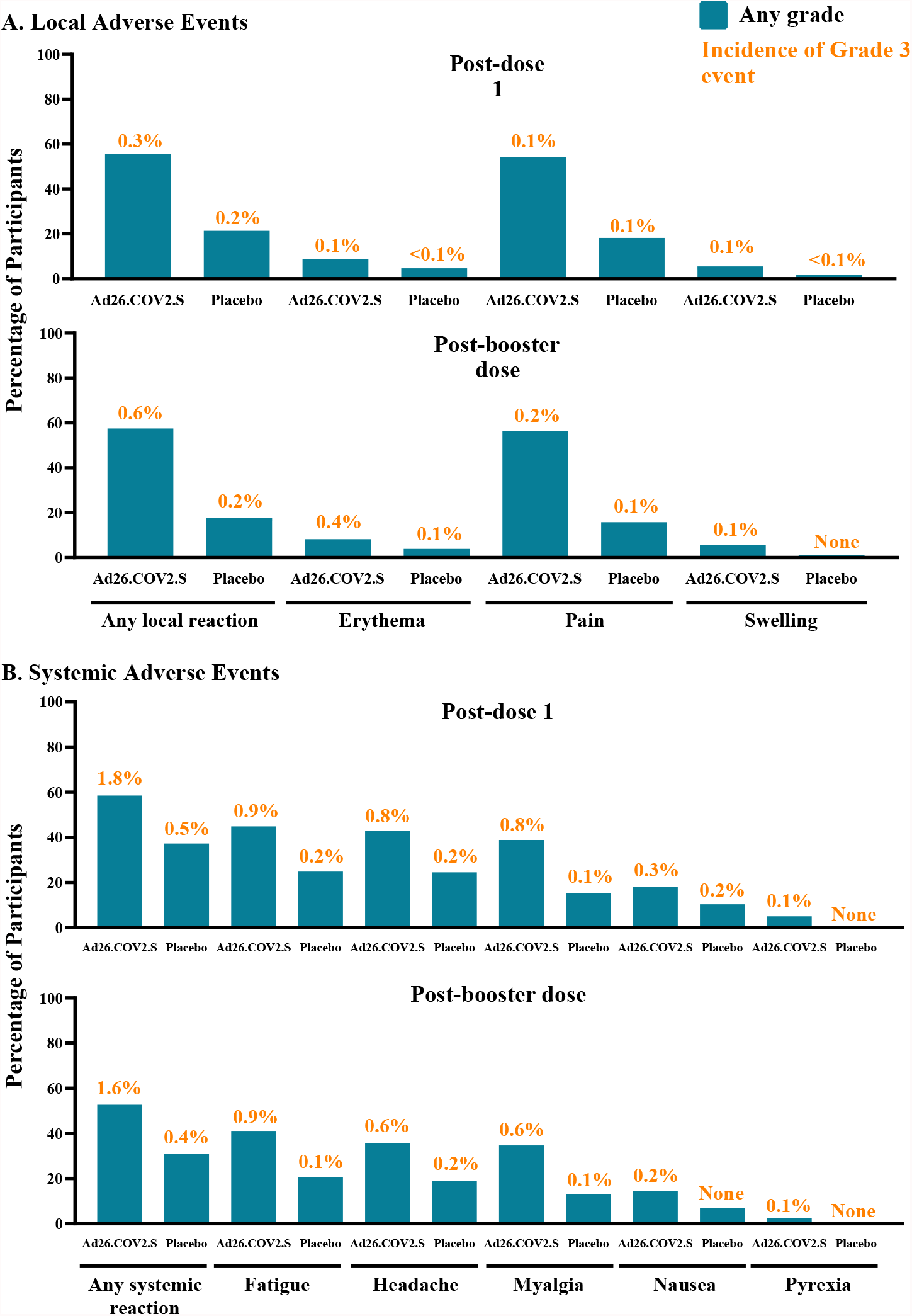
Solicited Local (A) and Systemic (B) Adverse Events Following Prime-Boost Vaccination Regimen in Adults (Safety Set). Pain was categorized as grade 1 (mild; does not interfere with activity), grade 2 (moderate; requires modification of activity or involves discomfort with movement), grade 3 (severe; incapacitating, causing inability to perform usual activities; use of narcotic pain relief), or grade 4 (potentially life-threatening; hospitalization or causing inability to perform basic self-care). Erythema and swelling were categorized as grade 1 (mild; 25 to 50 mm), grade 2 (moderate; 51 to 100 mm), grade 3 (severe; >100 mm), or grade 4 (potentially life-threatening; necrosis or leading to hospitalization). Systemic events were categorized as grade 1 (mild; minimal symptoms), grade 2 (moderate; notable symptoms not resulting in loss of work, school, or cancellation of social activities), grade 3 (severe; incapacitating symptoms resulting in loss of work, school, or cancellation of social activities), or grade 4 (life-threatening; hospitalization or inability to perform basic self-care). Fever was defined as grade 1 (mild; ≥38.0 to 38.4°C), grade 2 (moderate; ≥38.5 to 38.9°C), grade 3 (severe; ≥39.0 to 40.0°C), or grade 4 (potentially life-threatening; >40°C).

The most frequently reported solicited local AE after both vaccinations in the Ad26.COV2.S and placebo groups was vaccination site pain (dose 1, 54.2% and 18.2%, respectively; booster, 56.3% and 15.8%). Most solicited AEs were Grade 1-2 in severity. Grade 3 solicited local AEs were reported in 9 (0.3%) Ad26.COV2.S recipients after dose 1 and 10 (0.6%) recipients after boosting. No Grade 4 local AEs were reported. Local reactogenicity was transient, with median duration 1-3 days after any vaccination.

The most frequently reported solicited systemic AEs were fatigue, headache, and myalgia (**Figure 3, Table S6**). Fatigue was the most common systemic AE in the Ad26.COV2.S and placebo groups after both vaccinations (dose 1, 44.9% and 24.9%, respectively; booster, 41.1% and 20.6%). Grade 3 solicited systemic AEs were reported in 55 (1.8%) Ad26.COV2.S recipients following dose 1 and 25 (1.6%) post-booster. No Grade 4 systemic AEs were reported. Systemic reactogenicity was transient, with median duration 1-2 days post-vaccination.

Most unsolicited AEs were grade 1 or 2 in severity; unsolicited AEs of grade ≥3 severity and unsolicited events considered related to vaccination are in **Tables S7 and S8**.

Eleven participants experienced 13 SAEs considered related to the study vaccine (8 [0.1%] participants in the Ad26.COV2.S and 3 [<0.1%] participants in the placebo group (**Table S9**).

AEs of clinical interest are summarized in **Table S10**. No participant in the vaccine group reported an event that met the pre-established criteria for TTS^5^ during the double-blind phase. One placebo recipient had deep vein thrombosis on day 27 followed by pulmonary embolism in combination with thrombocytopenia on day 29. No cases of Guillain-Barré syndrome, immune thrombocytopenia, or encephalitis were reported during the double-blind phase.

Numerical imbalances were observed during the double-blind phase for arthritis (38 [0.2%] participants in the Ad26.COV2.S group vs 22 [0.1%] in the placebo group) and tinnitus (10 [0.1%] vs 5 [<0.1%]) (see **Table S10** for AEs within 28 days after each vaccination). Imbalances were seen for hemorrhagic disorders within 28 days after each vaccination (24 [0.2%] vs 14 [<0.1%] after dose 1, and 17 [0.2%] vs 7 [<0.1%] post-booster), mostly due to local injection site AEs. Of these, 8 were considered SAEs (**Table S10**). No numerical imbalances were observed for convulsions/seizures, Bell’s Palsy, deep vein thrombosis, pulmonary embolism, myocarditis, or pericarditis.

As of June 25, 2021, 5 participants in the FAS vaccine group discontinued the study due to an AE (cerebral hemorrhage, bipolar disorder/suicidal ideation, urticaria [non-serious and the only AE considered vaccine-related], benign prostatic hyperplasia, cerebral vertebral fracture). As of the data cutoff date, 17 deaths were reported in the double-blind phase (4 in the vaccine group [2 post-dose 1, and 2 post-booster] and 13 in the placebo-group). More deaths were Covid-19-related in the placebo group (7/13) than vaccine group (0). None of these deaths were considered related to study vaccine (**Table S9**).

## DISCUSSION

In this analysis of ENSEMBLE2 (COV3009), a booster dose of Ad26.COV2.S administered at a two-month interval elicited efficacy of 75.2% against moderate to severe–critical Covid-19 and 100% against severe–critical Covid-19 by ≥14 days after boosting. No cases requiring medical intervention and no Covid-19-related deaths were observed in the active arm of the study. Additionally, vaccination reduced the duration, number, and severity of symptoms in breakthrough cases, suggesting a shift from more severe to milder Covid-19 infection. Anamnestic response was demonstrated, as antibody titers increased from baseline approximately 40-fold by two weeks after the Ad26.COV2.S booster, as compared to 7.2-fold four weeks post-primary vaccination, coinciding with increased efficacy and suggesting that increased immunogenicity corresponds to increased protection. In the United States, efficacy after the Ad26.COV2.S booster (93.7%) was comparable to that seen with primary 2-dose regimens of mRNA vaccines (93%–95%).^6-8^

The Ad26.COV2.S booster appeared to improve efficacy against SARS-CoV-2 variants in ENSEMBLE2. Efficacy estimates against moderate to severe–critical Covid-19 caused by Alpha and Mu variants after primary single-dose Ad26.COV2.S vaccination (day 15-56) were 71.6% and 43.9%, respectively. This was consistent with the phase 3 ENSEMBLE (COV3001) trial (70.1% and 35.8%^9^), which assessed efficacy outcomes after a single dose of Ad26.COV2.S and was the basis for licensure/conditional approval in many countries.^10^ After the booster dose in ENSEMBLE2, efficacy estimates against Alpha and Mu were higher (94.2% and 63.1%), suggesting the benefit of boosting. In the United States, where Alpha became dominant during both studies,^11^ efficacy in the boosted population was 93.7% compared to 72.9% in ENSEMBLE. When the Delta variant surged in the United States from May to August 2021, Ad26.COV2.S single-dose effectiveness against Covid-19 declined, but effectiveness against hospitalization remained ≥80%.^12^ During emergence of Omicron, an Ad26.COV2.S booster dose 6 to 9 months after primary vaccination in South Africans elicited 85% vaccine effectiveness against hospitalization.^13^ These data support overall improved efficacy against variants after the Ad26.COV2.S booster, although conclusions for specific variants are limited. Attenuated protection in some countries or regions may be attributable to reduced vaccine efficacy against specific SARS-CoV-2 variants and low case numbers.^14-16^ The vaccine was also efficacious in participants with comorbidities in the current study.

Vaccine efficacy against moderate to severe–critical Covid-19 with onset ≥14 days after primary vaccination in ENSEMBLE2 (67%) was consistent with efficacy at the same time point in the final analysis of the double-blind phase of ENSEMBLE (56%).^9^ Between-study differences in efficacy may be attributed to differences in time, location, and epidemiologic pressure. Importantly, efficacy against severe–critical disease was high and consistent between the studies. Real-world data suggest these efficacies translate into clinical practice.^13,17-20^ Furthermore, Ad26.COV2.S elicited sustained CD8^+^ and CD4^+^ T-cell immune responses with cross-reactivity against Omicron,^21,22^ supporting the protection against this variant observed in a real-world study.^13^

The Ad26.COV2.S booster demonstrated an acceptable safety profile in adults ≥18 years old. Local and systemic reactogenicity was similar to that seen after the first dose, with no increase in adverse reactions post-booster. In the primary analysis of ENSEMBLE, more venous thromboembolic and convulsions/seizure events were seen after Ad26.COV2.S versus placebo.^10^ Conversely, in ENSEMBLE2, more of these events occurred after placebo dose 1. Although more noninfectious arthritis events occurred after Ad26.COV2.S in ENSEMBLE2, the converse was seen in ENSEMBLE (more after placebo), and no signal has been identified in post-marketing data. The hemorrhagic disorders imbalance in this study was mostly driven by events related to vaccine administration. These inconsistencies in AE occurrence between studies suggest imbalances may be attributable to chance.

There are limitations to this study. As ENSEMBLE2 was conducted at the peak of the Covid-19 wave of early 2021, when Covid-19 vaccines were first made available by EUA, it was no longer ethical to maintain the placebo control, leading to early unblinding. All participants could request unblinding to determine whether they qualified for Covid-19 vaccination outside the study and placebo recipients could receive the open-label crossover vaccination (timing varied by country). Unblinding/crossover reduced participant numbers receiving both doses and planned follow-up time in the double-blind phase and led to limited numbers of Covid-19 cases being available for evaluation of the booster dose; data within subgroups, including by variant, should be interpreted with caution. More participants in the placebo group than the Ad26.COV2.S group terminated prematurely, partly because after unblinding, placebo recipients terminated participation to receive another Covid-19 vaccine outside the study, and possibly due to non-study antibody testing. Most participants nevertheless completed the double-blind phase, and the study power remained strong. The person-years of follow-up in the PP and FAS sets were generally similar, indicating that blinding was properly maintained and bias minimized. Moreover, vaccine efficacy estimation methods accounted for imbalances of follow-up. Given the short follow-up and low numbers for some groups, the exact breadth and incremental protection of the booster dose merits further investigation.

A single dose of Ad26.COV2.S is efficacious against symptomatic Covid-19, and a booster administered 2 months later substantially increased vaccine efficacy, including against symptomatic and severe–critical Covid-19. Ad26.COV2.S received FDA and EMA authorization for use as a booster dose in October and December 2021, respectively.

## Supporting information

CONSORT Checklist

## Data Availability

The data sharing policy of Janssen Pharmaceutical Companies of Johnson & Johnson is available at https://www.janssen.com/clinical-trials/transparency. As noted on this site, requests for access to the study data can be submitted through Yale Open Data Access (YODA) Project site at http://yoda.yale.edu.

## ACKNOWLEDGMENTS

This work was supported by Janssen Vaccines & Prevention B.V. in collaboration with the Biomedical Advanced Research and Development Authority, the Department of Defense, the National Institutes of Health, and the COVID-19 Prevention Network. This project has been funded in whole or in part with federal funds from the Biomedical Advanced Research and Development Authority, part of the Office of the Assistant Secretary for Preparedness and Response at the U.S. Department of Health and Human Services (HHS), under Other Transaction Agreement HHSO100201700018C and from the National Institute of Allergy and Infectious Diseases (NIAID), NIH. We thank the individuals who volunteered to participate in this trial, the staff members at the trial locations, the Data Safety Monitoring Board, all investigators at the clinical sites, members of the Clinical Severity Adjudication Committee (Brian T. Garibaldi, MD, MEHP; Timothy E. Albertson, MD, MPH, PhD; Christian Sandrock, MD, MPH; Janet S. Lee, M.D.; Mark R. Looney, MD; Victor F Tapson, MD; Charles Shey Wiysonge, MD, MPhil, PhD), and the COV3009 (ENSEMBLE2) study team (Luis Humberto Anaya Velarde, Daniel Backenroth, Jisha Bhushanan, Börries Brandenburg, Vicky Cárdenas, Bohang Chen, Fei Chen. Polan Chetty, Pei-Ling Chu, Kimberly Cooper, Jerome Custers, Hilde Delanghe, Anna Duca, Tracy Henrick, Jarek Juraszek, Catherine Nalpas, Monika Peeters, Jose Pinheiro, Sanne Roels, Martin F. Ryser, Jose Salas, Samantha Santoro Matias, Ilse Scheys, Pallavi Shetty, Georgi Shukarev, Jeffrey Stoddard, Willem Talloen, NamPhuong Tran, Nathalie Vaissiere, Elisabeth van Son-Palmen, Jiajun Xu, Erin A. Goecker; Alexander L. Greninger, Keith R. Jerome, Pavitra Roychoudhury, and Simbarashe G. Takuva). We also thank Kurt Kunz, M.D., M.P.H., and Jill E. Kolesar, Ph.D., of Cello Health Communications/MedErgy for writing and editorial assistance funded by Janssen Global Services, LLC

