## Supplementary Appendix for "Efficacy and Safety of a Booster Regimen of Ad26.COV2.S Vaccine against Covid-19"

This appendix has been provided by the authors to give readers additional information about their work.

**ENSEMBLE2 Adjudication Committee**

| Committee Member | Affiliation | Location |
|---|---|---|
| Brian T. Garibaldi, MD, MEHP | Johns Hopkins University School of Medicine | Baltimore, MD, USA |
| Timothy E. Albertson, MD, MPH, PhD | University of California, Davis School of Medicine | Sacramento, CA, USA |
| Christian Sandrock, MD, MPH | University of California, Davis School of Medicine | Sacramento CA, USA |
| Janet S. Lee, M.D. | University of Pittsburgh | Pittsburgh, PA, USA |
| Mark R. Looney, MD | University of California, San Francisco | San Francisco, CA, USA |
| Victor F Tapson, MD | Cedars-Sinai Medical Center | Los Angeles, CA, USA |
| Charles Shey Wiysonge, MD, MPhil, PhD | South African Medical Research Council | Cape Town, South Africa |

**ENSEMBLE2 Study Group**

| Study Group Member | Affiliation | Location |
|---|---|---|
| Luis Humberto Anaya Velarde, MSc, MD | Janssen Research & Development | Beerse, Belgium |
| Daniel Backenroth, PhD | Janssen Pharmaceuticals | Raritan, NJ, USA |
| Jisha Bhushanan, B.Pharm | ICON Plc | Livingston, UK |
| Börries Brandenburg, PhD | Janssen Vaccines & Prevention B.V. | Leiden, The Netherlands |
| Vicky Cárdenas, PhD | Janssen Research & Development | Spring House, PA, USA |
| Bohang Chen, MD | Janssen Research & Development | Titusville, NJ, USA |
| Fei Chen, PhD | Janssen Research & Development | Raritan, NJ, USA |
| Polan Chetty, MSc | Janssen-Cilag UK | High Wycombe, UK |
| Pei-Ling Chu, PhD | Janssen Research & Development | Raritan, NJ, USA |
| Kimberly Cooper, MS | Janssen Research & Development | Spring House, PA, USA |
| Jerome Custers, PhD | Janssen Vaccines & Prevention, B.V. | Leiden, The Netherlands |
| Hilde Delanghe, MS | Janssen Research & Development | Beerse, Belgium |
| Anna Duca, BSN | Janssen Research & Development | Titusville, NJ, USA |
| Tracy Henrick, MS | Janssen Research & Development | Spring House, PA, USA |
| Jarek Juraszek, PhD | Janssen Vaccines & Prevention B.V. | Leiden, The Netherlands |
| Catherine Nalpas, PhD | Janssen-Cilag France | Issy-les-Moulineaux, France |
| Monika Peeters, MSc | Janssen Pharmaceutica NV | Beerse, Belgium |
| Jose Pinheiro, PhD | Janssen Research & Development | Raritan, NJ, USA |

| | | |
|---|---|---|
| Sanne Roels, PhD | Janssen Research & Development | Beerse, Belgium |
| Martin F. Ryser, MD | Janssen Research & Development | Beerse, Belgium |
| Jose Salas, PhD | Janssen Research & Development | Beerse, Belgium |
| Samantha Santoro Matias, BS | Janssen Research & Development | Pennsylvania, USA |
| Ilse Scheys, MS | Janssen Research & Development | Beerse, Belgium |
| Pallavi Shetty, MSc | Janssen Research & Development | Beerse, Belgium |
| Georgi Shukarev, MD | Janssen Vaccines & Prevention B.V. | Leiden, The Netherlands |
| Jeffrey Stoddard, MD | Janssen Research & Development | Raritan, NJ, USA |
| Willem Talloen, PhD | Janssen Pharmaceutica NV | Beerse, Belgium |
| NamPhuong Tran, MD | Janssen Research & Development | Los Angeles, CA, USA |
| Nathalie Vaissiere, MS | Janssen Research & Development | Beerse, Belgium |
| Elisabeth van Son-Palmen, HBO-B | Janssen Research & Development | Leiden, The Netherlands |
| Jiajun Xu, PhD | Janssen Research & Development | Shanghai, China |
| Erin A. Goecker, MS | University of Washington | Seattle, WA, USA |
| Alexander L. Greninger, MD, PhD, MS, MPhil | University of Washington; Fred Hutchinson Cancer Research Center | Seattle, WA, USA |
| Keith R. Jerome, MD, PhD | University of Washington; Fred Hutchinson Cancer Research Center | Seattle, WA, USA |
| Pavitra Roychoudhury, PhD | University of Washington; Fred Hutchinson Cancer Research Center | Seattle, WA, USA |
| Simbarashe G. Takuva, MBChB, MSc | University of the Witwatersrand; University of Pretoria; Fred Hutchinson Cancer Research Center | Seattle, WA, USA; Johannesburg, South Africa; Pretoria, South Africa |

**Participating Investigators**

The following Principal Investigators participated in the ENSEMBLE2 study

| Name | Institute | Location |
|---|---|---|
| Accini Mendoza, Jose Luis | IPS Centro Cientifico Asistencial S.A.S | Barranquilla, Colombia |
| Ahsan, Habibul | The University of Chicago Medicine | Illinois, USA |
| Arribas, Jose Ramon | Hospital Universitario La Paz | Madrid, Spain |
| Bahrami, Ghazaleh | Synexus Clinical Research US, Inc. | Kentucky, USA |
| Bajwa, Ali | Centex Studies, Inc. | Texas, USA |
| Baron, Mira | Palm Beach Research Center | Florida, USA |
| Bertoch, Todd | JBR Clinical Research | Utah, USA |
| Bethune, Claire | Derriford Hospital | Devon, United Kingdom |
| Bollen, Hilde | Huisartsen Alkerstraat | Alken, Belgium |
| Botelho-Nevers, Elisabeth | CHU Saint Etienne - Hôpital Nord | Loire, France |
| Burgess, Lesley | Tread Research | Western Cape, South Africa |
| Bush, Larry | JEM Research Institute | Florida, USA |
| Capeding, Maria Rosario | Tropical Disease Foundation | Manila, Philippines |
| Carr, Quito Osuna | MedPharmics – Albuquerque | New Mexico, USA |
| Cathie, Katrina | Southampton General Hospital | Hampshire, United Kingdom |
| Cereto Castro, Fernando | Hospital Quironsalud Barcelona | Barcelona, Spain |
| Chavoustie, Steven | Healthcare Clinical Data Inc. | Florida, USA |
| Chronos, Nicolas | ARS-Lake Oconee | Georgia, USA |
| Cicconi, Paola | Windrush Medical Practice | Oxfordshire, United Kingdom |
| Dargan, Paul | St Thomas' Hospital | Greater London, United Kingdom |
| Darton, Thomas | Royal Hallamshire Hospital | South Yorkshire, United Kingdom |
| Deane, Peter | Finger Lakes Clinical Research | New York, USA |
| Del Pozo, Jose Luis | Clinica Universidad de Navarra | Navarra, Spain |
| Derdelinckx, Inge | UZ Gasthuisberg / UZ Leuven | Leuven, Belgium |
| Dever, Michael | Clinical Neuroscience Solutions, Inc. | Florida, USA |
| DiBuono, Mark | Richmond Behavioral Associates ERG Clinical Research - New York PLLC | New York, USA |
| Doust, Matthew | Hope Research Institute | Arizona, USA |
| Echave-Sustaeta, Jose Maria | Hospital Universitario Quironsalud Madrid | Madrid, Spain |

| | | |
|---|---|---|
| Eder, Frank | Meridian Clinical Research LLC | New York, USA |
| Ellis, Kimberly | AMR Norfolk, Formerly Tidewater Associates, an AMR Company | Virginia, USA |
| Elzi, Stanton | Paradigm Clinical Research Centers, Inc. | Colorado, USA |
| Engelbrecht, Johannes | Dr JM Engelbrecht Trial Site | Western Cape, South Africa |
| Felton, Timothy | Royal Manchester Children's Hospital | Greater Manchester, United Kingdom |
| Flume, Patrick | Medical University of South Carolina (MUSC) | South Carolina, United States |
| Fragoso, Veronica | Texas Center for Drug Development, PA | Texas, USA |
| Freedman, Andrew | University Hospital of Wales | Cardiff, United Kingdom |
| Frentiu, Emilia | Hôpital de Brabois Adultes | Meurthe et Moselle, France |
| Galloway, Christopher | Headlands Research LLC | Florida, USA |
| Galtier, Florence | Hopital Saint Eloi | Herault, France |
| Garcia Alcaide | Clinica Universidad de Navarra (MAD) | Madrid, Spain |
| Garcia Diaz, Julia | Ochsner Medical Center | Louisiana, USA |
| Geller, Steven | Centennial Medical Group, PC | Maryland, USA |
| Girodet, Pierre-Olivier | Groupe Hospitalier Sud - Hôpital Haut-Lévêque | Gironde, France |
| Gler, Maria Tarcela | Makati Medical Center | Manila, Philippines |
| Glover, Richard | AMR Newton, Formerly Heartland Associates Newton, an AMR company | Kansas, USA |
| Green, Christopher | Queen Elizabeth Hospital | West Midlands, United Kingdom |
| Greenberg, Richard | University of Kentucky | Kentucky, USA |
| Griffin, Carl | Lynn Health Science Institute | Oklahoma, USA |
| Grobbelaar, Coert | The Aurum Institute Clinical Research Site - Pretoria | Gauteng, South Africa |
| Guancia, Adonis | Riverside Medical Center | Bacolod, Philippines |
| Harris, James | The South Bend Clinic Center for Research | Indiana, USA |
| Hassman, Michael | Hassman Research Institute | New Jersey, USA |
| Hellstrom-Louw, Elizabeth | Be Part Yoluntu Centre | Western Cape, South Africa |
| Hussen, Nazreen | Worthwhile Clinical Trials | Gauteng, South Africa |
| Isidro, Marie Grace Dawn | West Visayas State University Medical Center | Iloilo, Philippines |

| | | |
|---|---|---|
| Jain, Manish | Great Lakes Clinical Trials | Illinois, USA |
| João Filho, Esaú Custódio | HFSE - Hospital Federal dos Servidores do Estado do Rio de Janeiro | Rio de Janeiro, Brazil |
| Johnson, Daniel | Artemis Institute for Clinical Research, LLC | California, USA |
| Joseph, Natasha | Peermed CTC ta MERC Kempton Park | Gauteng, South Africa |
| Junquera, Patricia | Prestige Clinical Research Center, Inc. | Florida, USA |
| Kennedy, Patrick | Barts and The London School of Medicine & Dentistry | Greater London, United Kingdom |
| Kim, Kenneth | Ark Clinical Research - PARENT | California, USA |
| Kimmel, Murray | Optimal Research, LLC | Florida, USA |
| Konis, George | Woodland International Research Group, LLC | Arkansas, USA |
| Kutner, Mark | Suncoast Research Group, LLC | Florida, USA |
| Lacombe, Karine | Hôpital Saint-Antoine | Paris, France |
| Launay, Odile | Hôpital Cochin | Paris, France |
| Lazarus, Rajeka | Bristol Royal Infirmary | Bristol, United Kingdom |
| Lederman, Samuel | Altus Research, Inc. | Florida, USA |
| Lefebvre, Gigi | ARS-St. Petersburg | Florida, USA |
| Leroux-Roels, Isabel | Universitair Ziekenhuis Gent | Gent, Belgium |
| Lins, Muriel | AZ Sint-Maarten | Mechelen, Belgium |
| Liu, Edward | Jersey Shore University Medical Center | New Jersey, USA |
| Llewelyn, Martin | Royal Sussex County Hospital | East Sussex, United Kingdom |
| Mahomed, Akbar | Dr AA Mahomed Medical Centre | Mpumalanga, South Africa |
| Martinot, Jean Benoit | Pneumocare SPRL (Centre de recherche Clinique) | Namur, Belgium |
| McCaughan, Frank | Addenbrooke's Hospital | Cambridgeshire, United Kingdom |
| McGettigan, John | Quality of Life Medical & Research Center, LLC | Arizona, USA |
| Miller, Vicki | DM Clinical Research | Texas, USA |
| Mills, Anthony | Anthony Mills, MD, Inc. | California, United States |
| Molto Marhuenda, Jose | Hospital Universitari Germans Trias i Pujol | Barcelona, Spain |
| Myers, Paul | HCA Healthcare Research Institute | Tennessee, USA |
| Newman Lobato Souza, Tamara | Instituto de Infectologia Emílio Ribas | Sao Paulo, Brazil |
| Ochoa Mazarro, Maria Dolores | Hospital Universitario de La Princesa | Madrid, Spain |

| | | |
|---|---|---|
| Onate Gutierrez, Jose Millan | Centro Medico Imbanaco de Cali S.A | Cali, Colombia |
| Oshita, Masaru | Benchmark Research Sacramento | California, USA |
| Otero Romero, Susana | Hospital Universitari Vall d'Hebron | Barcelona, Spain |
| Overcash, Jeffrey Scott | Velocity Clinical Research, San Diego | California, USA |
| Paiva, Leonardo | Instituto Nacional de Infectologia Evandro Chagas (INI) - FIOCRUZ | Rio do Janeiro, Brazil |
| Palfreeman, Adrian | Leicester General Hospital | Leicestershire, United Kingdom |
| Patel, Rahul | Meridian Clinical Research | Maryland, USA |
| Patel, Suchet | Meridian Clinical Research – Baton Rouge | New York, USA |
| Pelkey, Leslie | Cherry Health | Michigan, USA |
| Pounds, Kevin | Synexus Clinical Research US, Inc. | Arizona, USA |
| Pouzar, Joe | Centex Studies, Inc. | Texas, USA |
| Pragalos, Antoinette | CTI Clinical Research Center | Ohio, USA |
| Presti, Rachel | Washington University | Missouri, USA |
| Price, David | Royal Victoria Infirmary | Tyne & Wear, United Kingdom |
| Ramalho Madruga, José Valdez | Centro de Referência e Treinamento em DST/AIDS - SP | Sao Paulo, Brazil |
| Rankin, Bruce | Accel Research Sites - DeLand Clinical Research Unit | Florida, USA |
| Riegel Santos, Breno | Hospital Nossa Senhora da Conceicao | Rio Grande do Sul, Brazil |
| Riesenberg, Robert | Atlanta Center for Medical Research | Georgia, USA |
| Riffer, Ernie | Synexus Clinical Research US, Inc. | Arizona, United States |
| Safirstein, Beth | Velocity Clinical Research, Hallandale Beach | Florida, USA |
| Salazar, Hernan | Endeavor Clinical Trials, P.A. | Texas, USA |
| Sanchez Vallejo, Gregorio | CEQUIN - Fundación Cardiomet Eje Cafetero | Armenia, Colombia |
| Seep, Michael | Centex Studies Inc. | Louisiana, United States |
| Short, Philip | Ninewells Hospital | Tayside Region, United Kingdom |
| Soentjens, Patrick | Institute of Tropical Medicine Antwerp | Antwerpen, Belgium |
| Solis, Joel | Centex Studies, Inc. | Texas, USA |
| Soriano Viladomiu, Alejandro | Hospital Clinic de Barcelona | Barcelona, Spain |
| Spangenthal, Selwyn | American Health Research, Inc. | North Carolina, USA |
| Spinner, Christoph | Klinikum rechts der Isar der TU Muenchen | Bayern, Germany |

| | | |
|---|---|---|
| Strout, Cynthia | Coastal Carolina Research Center, Inc | South Carolina, USA |
| Surowitz, Ronald | Health Awareness, Inc. | Florida, USA |
| Thalamas, Claire | Hopital Purpan and Hopital Rangueil | Haute Garonne, France |
| Trout, Guillermo | T Y C Inversiones S A S Grupsalud | Santa Marta, Colombia |
| Villalobos, Ralph Elvi | Philippine General Hospital | Manila, Philippines |
| Webster, Brian | Wilmington Health | North Carolina, USA |
| White, Alexander | Progressive Medical Research | Florida, USA |
| Williams, Hayes | Accel Research Site – Birmingham Clinical Research Unit | Alabama, USA |
| Winston, Alan | NIHR Imperial CRF | Greater London, United Kingdom |
| Zervos, Marcus | Henry Ford Hospital | Michigan, USA |

### Supplementary Methods

#### *Changes in Conduct and Protocol Amendments*

Six amendments were made to the protocol before the primary analysis.

With the first protocol amendment (25 September 2020), the dose level of Ad26.COV2.S was adjusted from $1\times10^{11}$ virus particle (vp) to $5\times10^{10}$ vp based on new data from the first-in-human (FIH) study VAC31518COV1001, including safety and immunogenicity data from Cohort 1a, safety data from Cohort 3, and immunogenicity data from the sentinel group of Cohort 3. Changes were also made throughout the protocol in response to the feedback received from health authorities, partners, and the community, including: the trigger for the evaluation of the primary endpoint was modified, adding one condition related to the number of Covid-19 cases (6) meeting the primary endpoint definition of moderate to severe/critical Covid-19 in the elderly population, that needed to be met; guidance was added regarding the use of licensed Covid-19 vaccines, when one might become available, during the study; the enrollment of participants aged ≥18 to <40 years was limited to approximately 20% of the total study population; the minimum target of recruited participants ≥60 years of age was adjusted from 25% to 30%; a secondary endpoint looking at the first occurrence of molecularly-confirmed, moderate to severe–critical Covid-19 with onset 14 days after the first vaccination was added; and an exploratory objective to assess the impact of the vaccine on other respiratory diseases was added. Minor errors and inconsistencies were corrected throughout the protocol.

With the second protocol amendment (27 November 2020), it was clarified that all participants with a reverse-transcriptase polymerase chain reaction (RT-PCR) positive finding for SARS CoV 2 from any source, even if asymptomatic, were to be followed until two consecutive negative PCRs were obtained. Text regarding biomarker evaluation of RNAseq analyses (PAXgene tube) was deleted and the total blood drawn was adjusted; based on Health Authority request, text regarding United Kingdom specific self-swabbing test was deleted. Text was also added introducing the utilization of tokenization and matching procedures to obtain the medical data of participants 5 years prior to enrollment of the participant until 5 years after study completion from consenting participants in the United States (US). Clarifications and minor editorial changes were made throughout the protocol.

With the third protocol amendment (18 December 2020), procedures were introduced to be followed in the event that an investigator received a request to unblind study participants who might become eligible to receive an authorized/licensed Covid-19 vaccine, if/when those were to become available. This change was made to ensure (1) that the participants were informed that no data was available regarding the safety of receiving two different Covid-19 vaccines, (2) that in the event the participant became unblinded, no further study vaccination would be permitted, (3) that unblinded participants would continue to be followed in this study in line with the Schedules of Activities, and that safety, efficacy, and immunogenicity evaluations would continue to be performed, although the data would be analyzed separately from the point of unblinding. Finally, clarifications and minor editorial changes were made throughout the protocol.

With the fourth protocol amendment (12 March 2021), procedures were introduced to be followed after Emergency Use Authorization (EUA) of Ad26.COV2.S in the US and approval of Protocol Amendment 4 by the local Health Authority and the Independent Ethics Committee/Institutional Review Board (IEC/IRB). At the unblinding visit, a single dose of Ad26.COV2.S vaccine would be offered to those enrolled participants who initially received placebo, leading to de facto unblinding of all participants and investigators and entry into the open-label phase of the study. In the new Schedule of Activities, the unblinding visit was scheduled as soon as reasonably practicable and preferably no later than 2 months following EUA in the United States and approval of Protocol Amendment 4 by the local Health Authority and the IEC/IRB. The study design was also updated, with the 2-dose placebo arm replaced by a 1 dose active vaccination arm, thus allowing assessment of the level of efficacy and duration of protection with a booster schedule of Ad26.COV2.S compared to a single dose schedule, as well as a direct comparison of the immunogenicity of the 2 Ad26.COV2.S schedules, whereby the single dose is introduced at different time points. With this change, newly enrolled participants were randomized 1:1 to the 1- or 2-dose regimen. All participants were encouraged to remain in the study and continue to be followed for efficacy/effectiveness, safety, and immunogenicity. Other changes include two new exploratory endpoints to gather more information regarding the efficacy against variants; new objectives and endpoints for the open-label phase of the study; additional minor editorial changes and clarifications throughout the protocol.

With the fifth protocol amendment (6 May 2021), additional safety measures were introduced following reports of adverse events suggesting an increased risk of thrombosis combined with thrombocytopenia following the use of the Ad26.COV2.S vaccine under EUA in the US. Thus, thrombosis with thrombocytopenia syndrome (TTS), a very rare event, was to be followed as an adverse event of special interest (AESI) that mandated reporting to the sponsor within 24 hours of awareness. In addition, blood samples would also be collected for baseline assessments of platelet count and storage for future coagulation-related testing. Finally, the protocol was adjusted to align with the latest vaccine risk language. Clarifications and minor editorial changes were made throughout the protocol.

With the sixth protocol amendment (15 October 2021), a booster vaccination with Ad26.COV2.S ($5x10^{10}$ vp dose level) was offered to all ongoing participants who received only a single vaccination with Ad26.COV2.S in the study (see **Supplementary Methods [Booster Vaccine Procedures]**). Other changes include: objectives for the open-label phase were updated and new statistical analyses comparing the 2-dose cohort with the 1-dose cohort and the booster cohort were added; capillary leak syndrome (CLS) was added to the list of reasons for the discontinuation of the Ad26.COV2.S vaccine; vaccine allocation per age group was allowed to vary regionally based on national recommendations; clarifications and minor editorial changes.

#### *Study Pause*

Safety was monitored in a blinded manner by the sponsor and a committee comprised of the principal investigator and representatives of the sponsor. Monitoring included the study vaccination pausing rules (applicable to Stage 1 only). If a study vaccination was considered to raise significant safety concerns (and/or a specific set of pausing criteria had been met), further vaccination of participants was paused. On 13 April 2021, following EUA in the US, a safety signal was identified for thrombosis with thrombocytopenia syndrome (TTS) in post-marketing data in the US, which was reported to the Vaccine Adverse Event Reporting System [VAERS]. As a precautionary measure, the CDC and FDA recommended a pause in the use of Ad26.COV2.S vaccine. In accordance with CDC and FDA guidance, the sponsor decided to voluntarily pause vaccinations in all studies in the program (including ENSEMBLE2 [Study COV3009]) on 13 April 2021. After assessment of the TTS safety signal by experts and the health authorities, vaccinations in the study were resumed upon implementation of protocol amendment 5: first in Colombia (22 May 2021), with the other countries following thereafter.

#### *Inclusion and Exclusion Criteria*

Every potential participant must have satisfied all of the following criteria to be enrolled in the study: participants must have provided consent indicating that he or she understood the purpose, procedures, and potential risks and benefits of the study, and was willing to participate in the study; participants must have been willing and able to adhere to the prohibitions and restrictions specified in this protocol. Participants must have been ≥18 to <60 years or ≥60 years of age on the day of signing the ICF; however, vaccine administration/recruitment in each age group may have changed per country based on national recommendation.

Participants must have been either in good or stable health according to the investigator's clinical judgment, including a BMI <30 kg/m$^2$. For Stage 1, participants may have had underlying illnesses not associated with increased risk of progression to severe Covid-19, as long as their symptoms and signs were stable and well-controlled. If participants were on medication for a condition not part of the comorbidities listed in Exclusion Criterion 14 of the protocol, the medication dose cannot have been increased within 12 weeks preceding the first vaccination and must be expected to remain stable for the duration of the study. For Stage 2, participants may have had a stable and well-controlled comorbidity, including comorbidities associated with an increased risk of progression to severe Covid-19. If participants were on medication for a comorbidity (including comorbidities associated with an increased risk of progression to severe Covid-19), the medication dose cannot have been increased within 12 weeks preceding the first vaccination and must be expected to remain stable for the duration of the study.

For patients with HIV infection, the infection must have been stable/well-controlled, including: CD4 cell count ≥300 cells/µL within 6 months prior to screening; HIV viral load <50 copies/mL within 6 months prior to screening; participant must have been on a stable antiretroviral treatment (ART) for 6 months (unless the change was due to tolerability, in which case the regimen could be for only the previous 3 months; changes in formulation were allowed, as were nationwide guidelines that require transition from one ART regimen to another), and the participant must have been willing to continue his/her ART throughout the study as directed by his/her local physician. Participants with ongoing and progressive comorbidities associated with HIV infection were excluded but comorbidities associated with HIV infection that had been clinically stable for the past 6 months were not an exclusion criterion. HIV diagnosis was laboratory-confirmed using any evidence (historic or current) from medical records, such as ELISA with confirmation with Western Blot or RT-PCR, or of a detectable viral load (country-specific regulatory approved tests).

Prior to randomization, participants must have been either not of childbearing potential or of childbearing potential and practicing an acceptable effective method of contraception and agreeing to remain on such a method of contraception from providing consent until 3 months after the last dose of study vaccine. Participants using contraceptives (birth control) should have done so consistent with local regulations regarding the acceptable methods of contraception for those participating in clinical studies. The use of hormonal contraception should have started at least 28 days before the first administration of study vaccine. Acceptable effective methods for this study included: hormonal contraception, combined (estrogen and progestogen containing) hormonal contraception associated with inhibition of ovulation (oral, intravaginal, or transdermal), progestogen-only hormonal contraception associated with inhibition of ovulation (oral, injectable, or implantable), intrauterine device; intrauterine hormone-releasing system; bilateral tubal occlusion/ligation procedure; vasectomized partner (the vasectomized partner should be the sole partner for that participant); sexual abstinence (defined as refraining from heterosexual intercourse from providing consent until 3 months after the last dose of study vaccine). All participants of childbearing potential must: have had a negative highly sensitive urine pregnancy test at screening; have had a negative highly sensitive urine pregnancy test on the day of and prior to each study vaccine administration.

Participants must have agreed to not donate bone marrow, blood, and blood products from the first study vaccine administration until 3 months after the last dose of the study vaccine; must have been willing to provide verifiable identification, had means to be contacted and to contact the investigator during the study; must have been able to read, understand, and complete questionnaires in the electronic clinical outcome assessment (eCOA; i.e., the COVID 19 signs and symptoms surveillance question, the e-Diary, and the electronic patient-reported outcomes [ePROs]).

Any potential participant who met any of the following criteria would be excluded from participating in the study: participant had a clinically significant acute illness (not including minor illnesses such as diarrhea or mild upper respiratory tract infection) or temperature ≥38.0ºC (100.4°F) within 24 hours prior to the planned first dose of study vaccine; randomization at a later date was permitted at the discretion of the investigator and after consultation with the sponsor.

Participants were excluded if they had a known or suspected allergy or history of anaphylaxis or other serious adverse reactions to vaccines or their excipients (including specifically the excipients of the study vaccine).

Participants were excluded if they had abnormal function of the immune system expected to have an impact on the immune response of the study vaccine and resulting from clinical conditions (e.g., autoimmune disease or potential immune-mediated disease or known or suspected immunodeficiency, or patient on hemodialysis) expected to impact immune response to study vaccine. Participants with clinical conditions stable under non-immunomodulator treatment (e.g., autoimmune thyroiditis, autoimmune inflammatory rheumatic disease such as rheumatoid arthritis) may have been enrolled at the discretion of the investigator. Non-immunomodulator treatment was allowed as well as steroids at a non-immunosuppressive dose or route of administration. Participants were excluded if they had abnormal immune function due to chronic or recurrent use of systemic corticosteroids at an immunosuppressive dose (≥2 weeks of daily receipt of 20 mg of prednisone or equivalent) within 6 months before administration of the first dose of study vaccine and during the study and administration of antineoplastic and immunomodulating agents or radiotherapy within 6 months before administration of the first dose of study vaccine and during the study.

Participants were excluded if they received treatment with Ig in the 3 months or exogenous blood products (autologous blood transfusions were not exclusionary) in the 4 months before the planned administration of the first dose of study vaccine or had any plans to receive such treatment during the study.

Participants were excluded if they received or planned to receive: licensed live attenuated vaccines within 28 days before or after planned administration of the first or subsequent study vaccinations; other licensed (not live) vaccines – within 14 days before or after planned administration of the 1st or subsequent study vaccinations.

Participants were excluded for the use of any coronavirus vaccine (licensed or investigational) other than Ad26.COV2.S at any time prior to vaccination and during the study.

Participants were excluded if they received: an investigational drug within 30 days (including investigational drugs for prophylaxis of Covid-19); used an invasive investigational medical device within 30 days; received investigational Ig or investigational monoclonal antibodies within 3 months; or received convalescent serum for Covid-19 treatment within 4 months or received an investigational vaccine (including investigational adenoviral-vectored vaccines) within 6 months before the planned administration of the first dose of study vaccine or were enrolled or planning to participate in another investigational study during the course of this study.

Participants were excluded if they had a history of an underlying clinically significant acute or chronic medical condition or physical examination findings for which, in the opinion of the investigator, participation would not be in the best interest of the participant (eg, compromise the well-being) or that could prevent, limit, or confound the protocol-specified assessments.

Participants were excluded if they had a contraindication to IM injections and blood draws, eg, bleeding disorders.

Participants were excluded if they had: major psychiatric illness, which in the investigator's opinion would compromise the participant's safety or compliance with the study procedures; could not communicate reliably with the investigator; were unlikely to adhere to the requirements of the study; or were unlikely to complete the full course of vaccination and observation.

In Stage 1, participants with comorbidities that were or might have been associated with an increased risk of progression to severe Covid-19 were excluded, including: participants with moderate-to-severe asthma; chronic lung diseases such as chronic obstructive pulmonary disease (COPD) (including emphysema and chronic bronchitis), idiopathic pulmonary fibrosis and cystic fibrosis; diabetes (including type 1 or type 2); serious heart conditions, including heart failure, coronary artery disease, congenital heart disease, cardiomyopathies, and pulmonary hypertension; moderate to severe high blood pressure; obesity (body mass index [BMI] ≥30 $kg/m^2$); chronic liver disease, including cirrhosis; sickle cell disease; thalassemia; cerebrovascular disease; neurologic conditions (eg, dementia); end stage renal disease; organ transplantation; cancer; HIV infection and other immunodeficiencies; hepatitis B infection; sleep apnea; participants who lived in nursing homes or long term care facilities; a history of or current

Parkinson's disease; seizures; ischemic strokes; intracranial hemorrhage; encephalopathy and meningoencephalitis; a history of malignancy within 1 year before screening (exceptions were squamous and basal cell carcinomas of the skin and carcinoma in situ of the cervix, or other malignancies with minimal risk of recurrence); a history of acute polyneuropathy (eg, Guillain-Barré syndrome); chronic active hepatitis B or hepatitis C infection

Participants were excluded who had surgery requiring hospitalization (defined as inpatient stay for longer than 24 hours or overnight stay), within 12 weeks before first vaccination, or would not have fully recovered from surgery requiring hospitalization, or had surgery requiring hospitalization planned during the time the participant was expected to participate in the study or within 6 months after the last study vaccine administration.

In areas where seroprevalence was predicted to be high, a screening serologic test for past or current infection with SARS-CoV-2 could have been performed (in a local laboratory) at the discretion of the sponsor to restrict the proportion of seropositive participants in the study.

***Booster Vaccine Procedures***

With Protocol Amendment 6 (15 October 2021), all ongoing study participants who had received only a single vaccination with Ad26.COV2.S were offered a booster vaccination with Ad26.COV2.S ($5x10^{10}$ vp dose level). Those participants who had already received 2 vaccinations with Ad26.COV2.S or any Covid-19 vaccinations outside of the study (including Ad26.COV2.S) at the time of local approval of Protocol Amendment 6 were not eligible to receive Ad26.COV2.S booster vaccination. According to Protocol Amendment 6, this booster dose was then administered in the open-label phase of the study, preferably within 6 to 12 months after the participant's first Ad26.COV2.S vaccination in the study and coinciding with the next scheduled visit; otherwise, the booster vaccination was not to occur earlier than 3 months after the participant's first Ad26.COV2.S vaccination. After the booster dose, a minimum follow-up of 6 months was implemented. All participants (whether consenting to the Ad26.COV2.S booster vaccination) were encouraged to remain in the study were monitored for safety, immunogenicity, and efficacy according to their original schedule.

In addition, participants could be offered a single Ad26.COV2.S booster dose under the following special conditions: (1) had met vaccination discontinuation criteria under previous amendments and had not withdrawn consent  from the study, received any Covid-19 experimental medication (including vaccines other than the study vaccine), had not experienced thrombosis with thrombocytopenia syndrome (TTS), heparin-induced thrombocytopenia (HIT), or capillary leak syndrome (CLS), and were not planning to receive another Covid-19 vaccine within 3 months of vaccination with the Ad26.COV2.S booster; (2) participants who were pregnant could receive the Ad26.COV2.S booster, if permitted by local regulations and if the investigator considered the potential benefits to outweigh the potential risks to the mother and fetus; (3) participants receiving systemic corticosteroids (chronic or recurrent use) or antineoplastic and immunomodulating agents or radiotherapy could receive the Ad26.COV2.S booster if permitted by local regulations and after being made aware that the safety and efficacy data in patients receiving these therapies in combination with Ad26.COV2.S is limited; (4) participants who had become infected with SARS-CoV-2 during the study could receive booster vaccination with Ad26.COV2.S vaccine (even if they had received steroid treatment, convalescent plasma, or monoclonal antibody treatment) after they had recovered from the acute illness and at least 1 month had passed and after being made aware that the safety and efficacy data on vaccinating a previously infected individual is limited. If participants had any other illness, booster vaccination with Ad26.COV2.S was deferred until after recovery from the acute illness.

Investigators were encouraged to follow health authority guidelines and to consider current local public health guidance regarding prioritization of immunization, when feasible. Investigators were permitted to prioritize those participants who had received their first dose at an earlier date relative to the booster vaccination.

#### *Efficacy Assessments*

Identification and molecular confirmation of SARS-CoV-2 infection and symptomatic Covid-19 were performed throughout the study (see **Supplementary Methods [Study Procedures for Reporting Symptoms of Covid-19]**). Molecularly-confirmed Covid-19 was used for the analysis of the Covid-19 case definition and was defined as a positive RT-PCR or another molecular test result from any available respiratory tract sample (eg, nasal swab sample, sputum sample, throat swab sample, saliva sample) that was confirmed by the central laboratory.

Covid-19 cases were assessed independently by a Clinical Severity Adjudication Committee (CSAC). Classification was based on the highest degree of severity during the observation period and on the CSAC's clinical judgment. The occurrence of Covid-19-related hospitalization and Covid-19-related complications (including hyperinflammatory syndrome, pneumonia, neurological or vascular complications, severe pneumonia, severe neurological or vascular events, acute respiratory distress syndrome, renal complications, sepsis, septic shock, and death) were monitored throughout the study.

As secondary objectives, vaccine efficacy in the prevention of asymptomatic SARS-CoV-2 infection and mild Covid-19 were analyzed. To identify asymptomatic infection, an immunologic test for SARS-CoV-2 seroconversion (ELISA and/or SARS-CoV-2 immunoglobulin assay) based on SARS-CoV-2 N protein was performed on: samples obtained at day 1 (before the first vaccination) and 14 days, 6 months, and 1 year after the booster vaccination; and on blood samples obtained at the unblinding visit (before open-label vaccination) and the booster vaccination visit (before booster vaccination).

#### *Covid-19 Case Definitions*

For the primary endpoint, all moderate and severe/critical Covid-19 cases were considered. Molecular confirmation of SARS-CoV-2 infection by a central laboratory was used for the analysis of the case definition.

Moderate Covid-19 cases were defined by (1) a SARS-CoV-2 positive RT-PCR or molecular test and (2) ≥2 of the following symptoms (new or worsening): fever (≥38.0°C or ≥100.4°F), cough, heart rate ≥90 beats/minute, shaking chills or rigors, muscle or body pain, headache, new loss of taste or smell, sore throat, red or bruised-looking feet or toes, gastrointestinal symptoms (nausea, vomiting, or diarrhea), or malaise (loss of appetite, generally unwell, fatigue, physical weakness); or ≥1 of the following signs or symptoms (new or worsening): shortness of breath, respiratory rate >20 breaths/minute, abnormal oxygen saturation (but above 93%), clinical or radiologic evidence of pneumonia, or radiological evidence of deep vein thrombosis.

Severe–critical Covid-19 cases were defined by a positive RT-PCR test for SARS-CoV-2 with ≥1 of the following features: respiratory failure; evidence of shock (systolic blood pressure <90 mm Hg, diastolic blood pressure <60 mm Hg, or requiring vasopressors); respiratory rate >30 breaths/minute; heart rate ≥125 beats/minute; oxygen saturation of 93% or less (ambient air at sea level), or a ratio of the partial pressure of oxygen to the fraction of inspired oxygen <300 mm Hg; intensive care unit admission; significant acute renal, hepatic, or neurologic dysfunction, or death.

Mild Covid-19 cases were defined by a positive RT-PCR test for SARS-CoV-2 and at any time during observation, ≥1 of the following symptoms, but not meeting the definition for moderate or severe–critical Covid-19: fever (≥38.0°C or ≥100.4°F), sore throat, malaise (loss of appetite, generally unwell, fatigue, physical weakness), headache, muscle pain (myalgia), gastrointestinal symptoms (nausea, vomiting, or diarrhea), cough, chest congestion, runny nose, wheezing, skin rash, eye irritation or discharge, chills, new or changing olfactory or taste disorders, red or bruised-looking feet or toes, or shaking chills or rigors.

***Study Procedures for Reporting Symptoms of Covid-19***

All participants were given access to an electronic clinical outcome assessment (eCOA) digital tool, which was used to collect the following: Covid-19 signs and symptoms surveillance info for all participants; electronic patient-reported outcome (ePRO) using the Symptoms of infection with Coronavirus-19 (SIC), provided in the local language in accordance with local guidelines; body temperature, and pulse oximetry results) for all participants at baseline and in the event of Covid-19-like signs and symptoms; and e-Diary data (Safety Subset only) for 7-day reactogenicity (solicited signs and symptoms, including body temperature). All eCOA assessments were to be conducted prior to any tests, procedures, or other consultations, thus limiting bias in participant responses. The baseline SIC was completed before the first vaccine administration. All participants were also provided a thermometer to measure body temperature, a kit to self-collect nasal swabs samples and saliva samples, and a pulse oximeter at baseline to collect physical exam findings during a Covid-19 episode.

Participants with symptoms completed the SIC daily and took pulse oximeter readings three times daily from the day symptoms began for the duration of illness. Based on the information in the SIC, site investigators contacted participants on the day or day after symptoms began to assess whether the symptoms qualified as suspected Covid-19. All participants with Covid-19-like signs or symptoms meeting the prespecified criteria for suspected Covid-19 (see **Supplementary Methods [Covid-19 Case Definitions]**) and all participants with at least one positive RT-PCR test for SARS-CoV-2 were considered to have a Covid-19 episode and followed the following procedures. Participants with symptoms took nasal swabs at home on the day or day after symptoms began (as soon as possible after prespecified criteria for suspected Covid-19 were met) every other day until there were two consecutive negative swabs; in addition, qualified site staff were available to collect nasal swab samples as needed. Swab samples were transferred to the study site as soon as possible after collection, where they were refrigerated in sterile saline and stored in a freezer before shipment to the central laboratory on dry ice.

For the duration of follow-up of Covid-19 episodes, all participants with Covid-19 signs and symptoms measured body temperature daily (oral route preferred, or according to the local standard of care); the highest temperature each day was recorded via the ePRO (SIC) in the eCOA. During each suspected Covid-19 episode, vital signs were measured by qualified study site staff, preferably before collection of nasal swabs and blood draws, including: supine systolic and diastolic blood pressure, heart rate, respiratory rate, oxygen saturation, and body temperature.

Participants with at least 1 SARS-CoV-2 positive nasal swab collected on the day or day after symptoms began or on visits 2-4 days after symptoms began, were asked to return to the site approximately two weeks later (28 ± 7 days), at which time a blood sample was drawn for seroconfirmation of SARS-COV-2 infection (antibody).

Participants with a Covid-19 episode were followed until 14 days after symptom onset or until there were two consecutive negative swabs, whichever came last. If two consecutive nasal swabs negative for SARS-CoV-2 were not available, participants could cease collection of nasal swabs and saliva samples at the two-week visit, if they experienced at least two consecutive days with no Covid-19-related signs and symptoms.

#### *Safety Definitions*

An adverse event (AE) is any untoward medical occurrence in a clinical study participant administered a pharmaceutical (investigational or non-investigational) product. An AE does not necessarily have a causal relationship with the vaccine, and can therefore be any unfavorable and unintended sign (including an abnormal finding), symptom, or disease temporally associated with the use of a medicinal (investigational or non-investigational) product, whether or not related to that medicinal (investigational or non-investigational) product.

Serious adverse events (SAEs) were any AE that resulted in death, was life-threatening, required hospitalization (or prolonging of existing hospitalization), resulted in persistent or significant disability, was a congenital anomaly, was a suspected transmission of any infectious agent via a medicinal product, or was considered medically important based on investigator judgment.

Medically-attended adverse events (MAAEs) were any AEs with medically-attended visits (excluding routine study visits), including hospital, emergency room, urgent care clinic, or other visits to or from medical personnel for any reason. New onset of chronic diseases was collected as part of MAAEs.

Suspected adverse events of special interest (AESI) included thrombotic events and thrombocytopenia (defined as a platelet count below 150,000/μL) and were reported from the moment of vaccination until the end of the study/early withdrawal. From the time of local approval of Protocol Amendment 5 onwards, TTS was considered an AESI.

Other adverse events of interest were not prespecified per protocol, but AEs were selected for analysis based on the lists of Adverse Events of Interest proposed by expert groups and regulatory authorities for safety data collection for Covid-19 vaccines, including: Brighton

Collaboration (Law and Sturkenboom 2020)[1]; ACCESS protocol (Willame 2021)[2]; US CDC (preliminary list of AESI for VAERS surveillance) (Shimabukuro 2021)[3]; Medicines and Healthcare products Regulatory Agency (unpublished guideline).

Adverse events of clinical interest included the following conditions:

- Encephalitis, including acute disseminated encephalomyelitis (ADEM) and meningoencephalitis
- Guillain-Barré Syndrome
- Transverse myelitis
- Bell's palsy
- Multiple sclerosis, including optic neuritis
- Sensorineural Hearing loss
- Generalized convulsion
- Autoimmune thyroiditis
- Immune thrombocytopenia
- Other types of thrombocytopenia
- Acute aseptic arthritis
- Cardiac inflammatory disorders, including myocarditis and pericarditis
- Microangiopathy
- Heart failure
- Stress cardiomyopathy
- Coronary artery disease, incl. acute myocardial infarction
- Arrhythmia
- Deep vein thrombosis
- Non-hemorrhagic stroke
- Pulmonary embolism
- Hemorrhagic stroke
- Disseminated intravascular coagulation
- Acute kidney failure
- Acute hepatic failure
- Anaphylaxis
- Type 1 diabetes mellitus
- Narcolepsy

***Clinical Severity Adjudication Committee***

A Clinical Severity Adjudication Committee (CSAC) was used for adjudication of the severity of Covid-19 cases. The CSAC reviewed all cases in the study, taking into account all available relevant information at the time of adjudication. In addition, the CSAC reviewed cases requiring medical intervention (such as a composite endpoint of hospitalization, intensive care unit (ICU) admission, mechanical ventilation, and ECMO, linked to objective measures such as decreased oxygenation, X-ray, or CT findings) and the onset of cases. The CSAC's decisions were considered the definitive classification of the case. The role of the Committee and adjudication process was provided in the committee's charter.

***Laboratory Methods***

Next-generation sequencing was performed using the Swift Biosciences SNAP Version 2.0 kit with 1% and baseline polymorphisms defined with a cut-off of 15%, performed at the Virology Laboratory of the University of Washington, Department of Laboratory Medicine and Pathology (UW Virology). The SNAP assay utilizes multiple overlapping amplicons in a single tube to prepare ready-to-sequence libraries. The primer pairs used in SNAP are designed for generating libraries from first- or second-strand cDNA produced from viral isolates or clinical specimens. This design enables successful SARS-CoV-2 library preparation from samples with low viral titers and provides powerful solutions for the confident detection of nucleotide variants. The Swift Biosciences SARS-CoV2 Version 2.0 kit (Catalog # CovGl V2-96) is optimized for Illumina sequencing platforms and for additional genome coverage. A full clinical validation with determination of analytical sensitivity and specificity, limit of detection, accuracy, and assay precision (reproducibility and repeatability) has been performed.

Amino acid substitution profiles were created to identify the different SARS-CoV-2 lineages and variants (including variants of concern and variants of interest). The SARS-CoV-2 Wuhan-Hu1 variant with the addition of amino acid variation D614G (B.1 lineage) was taken as the reference sequence. Amino acid variations were defined as changes from the reference sequence. Amino acid substitutions and deletions were used to identify the variants; the most conserved S mutations of each lineage were selected to exclude as few sequences as possible but also not to misclassify the lineages.

Because full genomes were not available, the lineages were approximated based on the S-protein amino-acid sequence. The assignment of approximate lineages based on the predefined set of S amino acid substitutions agreed with the known circulating lineages in the considered countries based on the GISAID database (GISAID 2021).[4]

To classify SARS-CoV-2 S sequences into their most probable lineages, the following substitution profiles were used:

- B.1.1.7 (Alpha): H69del, V70del, Y144del, N501Y, A570D, D614G, P681H, S982A, T716I, D1118H
- B.1.351 (Beta): K417N, E484K, N501Y, D614G, A701V
- P.1 (Gamma): K417T, E484K, N501Y, D614G, H655Y
- B.1.617.2/AY.1/AY.2 (Delta): L452R, T478K, D614G, P681R
- B.1.427/429 (Epsilon): W152C, L452R, D614G
- B.1.525 (Eta): A67V, H69del, V70del, Y144del, E484K, D614G, Q677H, F888L
- B.1.526 (Iota): L5F, T95I, D253G, D614G, E484K, A701V
- B.1.617.1 (Kappa): G142D, E154K, L452R, E484Q, D614G, P681R
- C.37 (Lambda): R246del, S247del, Y248del, L249del, T250del, P251del, G252del, D253N, L452Q, F490S, D614G, T859N
- P.3 (Theta): L141del, G142del, V143del, A243del, L244del, E484K, N501Y, D614G, P681H, E1092K, H1101Y, V1176F
- P.2 (Zeta): E484K, D614G, V1176F (not part of P.1 or P.3)
- B.1.621 (Mu): T95I, Y144T, Y145S, ins145N, R346K, E484K, N501Y, D614G, P681H, D950N
- C.36.3: W152R, R346S, L452R, D614G, Q677H, A899S

- R.1: W152L, E484K, D614G, G769V
- B.1.1.519: T478K, D614G, P681H, T732A

Samples for viral sequencing were taken throughout the study schedule; however, sequencing was triggered at the discretion of the virologist considering the SARS-CoV-2 viral load levels and the limitations of the sequencing assays.

#### *Statistical Analysis Methods*

<u>Population Analysis Sets</u>

The full analysis set (FAS) comprised all randomized participants who received at least one documented administration of study vaccine or placebo in the double-blind phase, regardless of the occurrence of protocol deviations and serostatus at enrollment. Safety analyses were performed on the FAS. Vaccine efficacy analyses could be repeated in the FAS.

The safety subset included approximately 6000 participants from the FAS (~3000 from the vaccine group and ~3000 from the placebo group; and including at least 2000 from the older age group [≥60 years of age]). The safety subset was used for the analysis of solicited AEs for 7 days after each vaccination and unsolicited AEs for 28 days after each vaccination.

The per-protocol efficacy set (PP; primary efficacy analysis set) included participants in the FAS who received 2 doses of vaccine or placebo in the double-blind phase, who were seronegative or missing by serology at the time of first vaccination and at 14 days after the second vaccination (day 71), and who had no major protocol deviations before unblinding that were judged to possibly impact the efficacy of the vaccine. Participants positive by PCR at baseline were excluded. Participants who were unblinded/became aware of their study vaccine allocation were thereafter excluded from the PP population. The PP set was the main analysis population for efficacy endpoints. Some violations did not lead to exclusion from the PP set, but led to the exclusion of data from the time point of the major protocol deviation onwards, including the use of prohibited concomitant medications, or existence of medical conditions found likely to impact the efficacy of the vaccine based on a medical review documented prior to database lock; administration of another SARS-CoV-2 vaccine before or during the double-blind phase; or administration of the open-label vaccination with Ad26.COV2.S (after receiving placebo) prior to being unblinded. Missing PCR or serology results at baseline or day 71 were considered as negative.

The immunogenicity subset included ~400 participants in the FAS, located in the US or UK, from whom additional blood samples were collected for analysis of humoral immune responses. To maintain ~400 participants, additional participants might be added to the original subset during the open-label phase of the study, thereby creating new immunogenicity subsets.

The per-protocol immunogenicity (PPI) set included all randomized participants who received 2 doses of study vaccine in the double-blind phase, including participants in the Immunogenicity Subset for whom immunogenicity data were available, and excluding participants with major protocol deviations expected to impact the immunogenicity outcomes. Additionally, for those participants who experienced a molecularly-confirmed SARS-CoV-2 event, samples taken after the event and samples taken outside protocol windows were not taken into account in the assessment of the immunogenicity. The PPI population was used for the primary immunogenicity analysis. For key tables, sensitivity immunogenicity analyses were also performed on the FAS, including participants who were part of the Immunogenicity Subset for whom immunogenicity measures were available. Excluded samples could also be taken into account in the sensitivity analysis. Analyses of vaccine immunogenicity were based on the PPI population.

The per-protocol first dose efficacy set (PPFD) included participants in the FAS who received at least the first dose of study vaccine in the double-blind phase and were seronegative or missing serology at baseline, and who had no major protocol deviations before unblinding that were judged to possibly impact the efficacy of the vaccine. Those who were PCR positive at baseline were excluded.

<u>Sample Size Calculation</u>

For the double-blind phase, the study target number of events (TNE) was determined using the following assumptions: (1) VE for molecularly-confirmed, moderate to severe–critical SARS-CoV-2 infection of 65%; (2) type I error rate $\alpha=2.5\%$ to evaluate VE of the vaccine regimen using the exact binomial distribution; (3) randomization ratio of 1:1 for active versus placebo. If assumptions 1–3 were true, approximately 90% power could reject the null hypothesis of H0: VE≤30%, according to the primary endpoint case definition of moderate to severe/critical Covid-19.

Events were defined as the first occurrence of molecularly-confirmed, moderate to severe–critical Covid-19 according to the case definition in the PP set at least 14 days after the booster vaccination (day 71).

Under the assumptions above, the TNE required to compare the active vaccine versus placebo was 104.

For the open-label phase, a superiority evaluation was performed for the primary objective of the open-label phase for comparison between the 2-dose and the 1-dose schedule using a null hypothesis of relative VE=0%. Assuming a relative VE=50% or more, at a one-sided 2.5% significance level with 90% power, the TNE was 92.

<u>Hypothesis Testing</u>

This study was designed to test the hypothesis for the primary endpoint of vaccine efficacy (VE) against molecularly-confirmed, moderate to severe–critical Covid-19 infection with onset at least 14 days after booster vaccination with Ad26.COV2.S versus placebo, in the per-protocol, including all events with and without comorbidities: the null hypothesis (H0: VE ≤ 30% versus H1: VE>30%) was evaluated at a 2.5% one-sided significance level. Statistical hypothesis testing followed the prespecified scheme for family-wise type I error control.

A successful primary efficacy conclusion required establishing the hypothesis H1: VE>30% for the primary endpoint.

If the primary hypothesis testing was successful, each secondary endpoint was evaluated separately against a null hypothesis employing a lower limit (H0: VE ≤0% versus H1: VE >0%) in the per-protocol set, including all events with onset at least 14 days after the booster vaccination at the time of the primary analysis. The analysis of secondary endpoints was conducted with the family-wise error rate controlled at 2.5% and was adjusted for testing of multiple endpoints using a graphical approach following the method of Bretz et al, 2009.[5]

Only after establishing success for the primary endpoint, the first secondary hypothesis (any symptomatic infection: burden of disease) was evaluated at α=2.5%. If success was established on the first secondary endpoint, the alpha that was passed on to the hypotheses of "Any infection" and "Severe events" was 2.5%/2 (α=1.25%). The available alpha for the hypothesis test on the endpoint of asymptomatic cases and medical intervention depended on the rejection of the hypotheses on "Any infection" and "severe events."

Primary Analysis

The primary efficacy analysis of the double-blind phase could begin once all participants reached the open-label phase or had been unblinded. The analysis could also begin, depending on the operational implementation of unblinding visits, as well as the stage of the pandemic, when a minimum of 90% of the study population had reached the open-label phase/been unblinded.

Evaluation of the primary hypothesis and vaccine efficacy were estimated using an exact Poisson regression, taking into account the follow-up time, in the PP set including seronegative participants. To evaluate the primary endpoint, hypothesis testing was performed as described above (H0: VE ≤30% versus H1: VE >30%).

For participants with molecularly-confirmed Covid-19, the follow-up time was the time between the first vaccination and the time of onset of the case. For participants without molecularly-confirmed Covid-19, the follow-up time was the time since first vaccination until the last available measurement or at the end of the double-blind phase/study discontinuation, whichever came first.

The time to first occurrence of molecularly-confirmed moderate or severe–critical Covid-19 was the time between the first vaccination until the time of onset of the case. Participants without moderate and severe–critical Covid-19 were censored at the last available measurement or at end of double-blind phase/study discontinuation, whichever came last.

Subgroup analyses were performed to supplement the primary analysis, including: for age group (18 to <60 years, ≥60 years) and presence of comorbidities (yes/no); these analyses each employed a descriptive summary including (unadjusted) 95% confidence intervals to describe the VE in each subpopulation using the same methods. Depending on the recruited study population, the ≥60 years subgroup could have been further subcategorized (≥70 years, ≥80 years). Safety and efficacy analysis were further summarized descriptively for subgroups (sex, race, ethnicity, age categories, country, region [Europe, Latin-America, The Philippines, South Africa, United States], presence of baseline comorbidity, baseline seropositivity status, frailty index, baseline $PCR^{+}$ test) and variants known to circulate to date of the finalization of the Statistical Analysis Plan. No formal hypothesis testing was performed in these subgroups.

Vaccine efficacy against moderate to severe–critical disease was summarized for the following time intervals using VE and associated 95% confidence intervals: day 2-14, day 15-56, day 57-70, day 57 to the end of the double-blind phase, day 71 to the end of the double-blind phase, and VE against moderate to severe–critical disease with onset at least 14 days after double-blind vaccination, regardless of the second dose (PPFD set, from 14 days after the first dose [day 15] through the end of the double-blind phase).

<u>Supportive Analyses</u>

In a supportive analysis of the primary efficacy endpoint, vaccine efficacy was estimated using a Cox proportional hazards regression model with an associated two-sided 95% confidence interval (Wald test) and stratified for age (>=60 yrs., <60 yrs.) and comorbidities (with or without comorbidities) using the actual strata.

The primary efficacy analysis and supportive analysis were repeated in FAS participants who were seronegative with onset of molecularly-confirmed, moderate and severe–critical Covid-19 one day after vaccination in the double-blind phase. In addition, the primary efficacy analysis and supportive analyses were repeated regardless of serostatus and for seropositive participants if >6 moderate and severe/critically ill Covid-19 observed in seropositive participants.

<u>Missing Data</u>

Missing data were not imputed, and participants who did not report an event/concomitant medication were considered as participants without an event/concomitant medication. An AE with a missing severity or relationship was considered as an AE reported but was considered as not reported for the analysis of severity or relationship.

<u>Unblinding and Censoring</u>

Data for participants who received a Covid-19 vaccine outside the study (regardless of whether the vaccine is Ad26.COV2.S or a different vaccine) were censored for all analyses on the date of unblinding or the date of non-study vaccination, whichever came first.

<u>Interim Analyses</u>

Interim analyses in the form of continuous monitoring on potential harm set according to the following boundaries:

1. Potential Harm of Symptomatic Cases (FAS) monitored after every event starting from the 12th event
2. Potential Harm of Severe Cases (FAS) monitored after every event starting from the 5th event

All boundaries for potential harm were monitored by the Statistical Support Group (SSG). If a boundary was crossed, the SSG would then immediately inform the Chair of the Independent Data Monitoring Committee (IDMC) through secure communication procedures, leading to the convening of the IDMC to evaluate the totality of the data and provide a recommendation to the Sponsor.

### Supplementary Figures

#### Figure S1. Kaplan-Meier Curves of Follow-up Time in the Double-blind Phase (Full Analysis Set)

vp, viral particles.

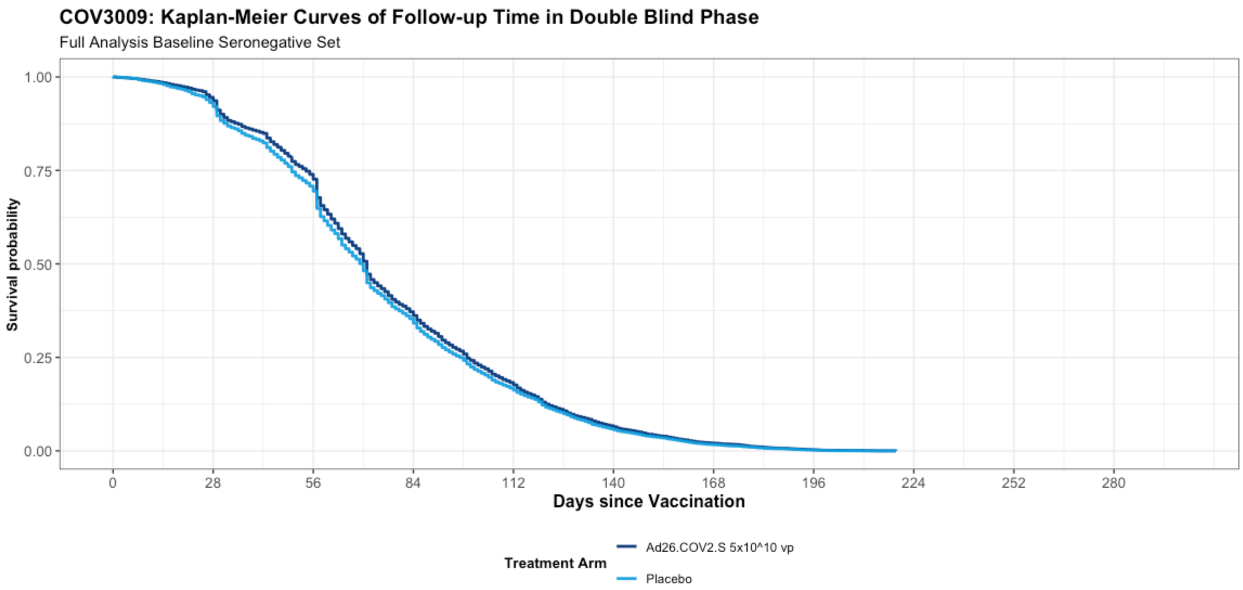

**Figure S2. Cumulative Incidence of First Occurrence of Molecularly-Confirmed Moderate to Severe–Critical or Severe–Critical Covid-19 Cases with Onset at Least 1 Day After Booster Vaccination (Per-Protocol Set).**

The cumulative incidence of molecularly-confirmed moderate to severe–critical cases of Covid-19 with onset at least 1 day after booster vaccination in the per-protocol (PP) set.is shown in Panel A. The cumulative incidence of molecularly-confirmed moderate to severe–critical cases of Covid-19 with onset at least 1 day after booster vaccination and due to Alpha (B.1.1.7) and Mu (B.1.621) variants in the PP set are shown in panels B and C, respectively. Cases included for analysis were centrally confirmed cases in the PP set among participants who were seronegative at baseline. The PP set included participants who received 2 doses of vaccine or placebo in the double-blind phase, who were seronegative (or missing) by serology at the time of the first vaccination (day 1) and at 14 days after the second vaccination (day 71), who were negative by PCR at baseline, and who had no major protocol deviations before unblinding that were judged to possibly impact the efficacy of the vaccine. Ad26 5e10 vp, Ad26.COV2.S given at a dose level of $5\times10^{10}$ viral particles; CI, confidence interval; Covid-19, coronavirus disease 2019; vp, viral particles.

A)

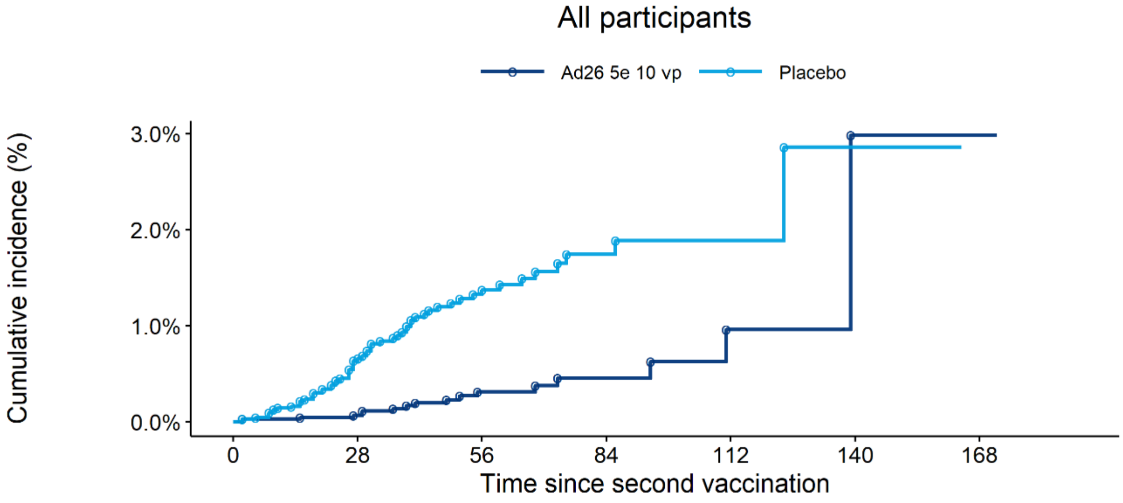

Participants at risk

| | | | | | | | |
|---|---|---|---|---|---|---|---|
| Ad26 5e 10 vp | 7475 | 4493 | 2288 | 873 | 282 | 44 | 1 |
| Placebo | 6980 | 4168 | 2026 | 758 | 228 | 30 | 0 |

Number of cases

| | | | | | | | |
|---|---|---|---|---|---|---|---|
| Ad26 5e 10 vp | 0 | 4 | 12 | 14 | 16 | 17 | 17 |
| Placebo | 0 | 33 | 55 | 60 | 61 | 62 | 62 |

B)

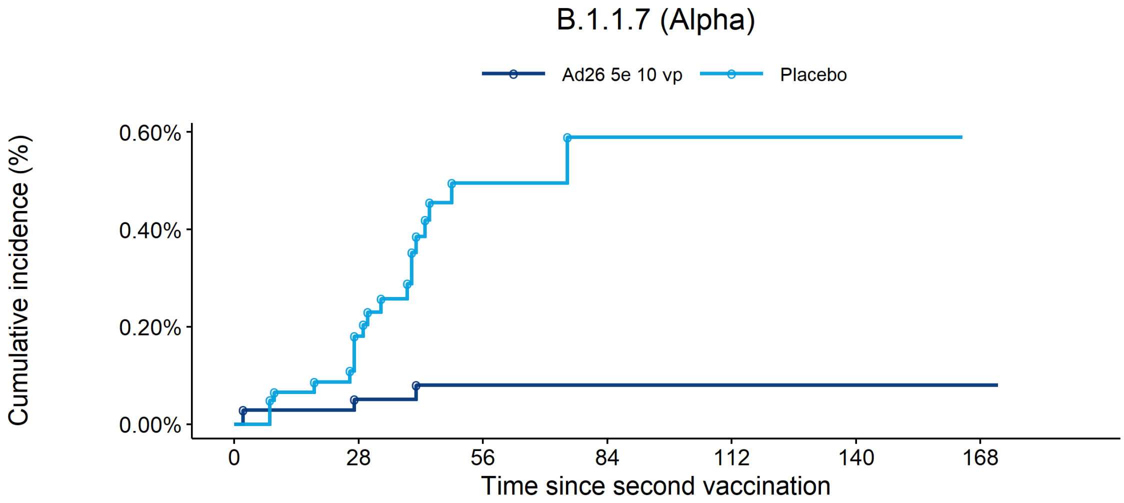

Participants at risk

| | | | | | | | |
|---|---|---|---|---|---|---|---|
| Ad26 5e 10 vp | 7475 | 4493 | 2288 | 873 | 282 | 44 | 1 |
| Placebo | 6980 | 4168 | 2026 | 758 | 228 | 30 | 0 |

Number of cases

| | | | | | | | |
|---|---|---|---|---|---|---|---|
| Ad26 5e 10 vp | 0 | 3 | 4 | 4 | 4 | 4 | 4 |
| Placebo | 0 | 9 | 19 | 20 | 20 | 20 | 20 |

C)

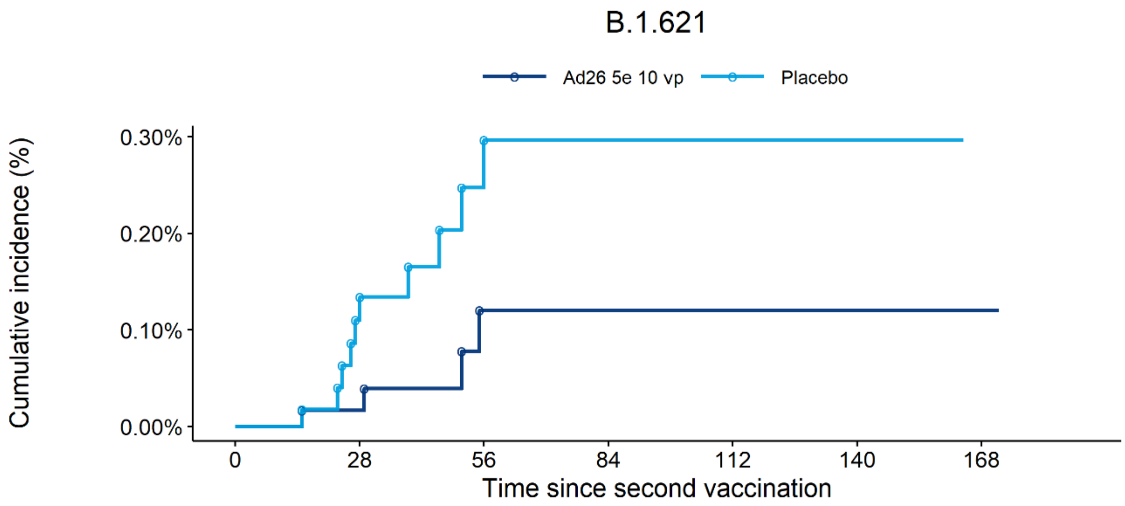

Participants at risk

| | | | | | | | |
|---|---|---|---|---|---|---|---|
| Ad26 5e 10 vp | 7475 | 4493 | 2288 | 873 | 282 | 44 | 1 |
| Placebo | 6980 | 4168 | 2026 | 758 | 228 | 30 | 0 |

Number of cases

| | | | | | | | |
|---|---|---|---|---|---|---|---|
| Ad26 5e 10 vp | 0 | 1 | 4 | 4 | 4 | 4 | 4 |
| Placebo | 0 | 6 | 10 | 10 | 10 | 10 | 10 |

**Figure S3. Case Accrual by SARS-CoV-2, Including Cases Not Sequenced (Full Analysis Set)**

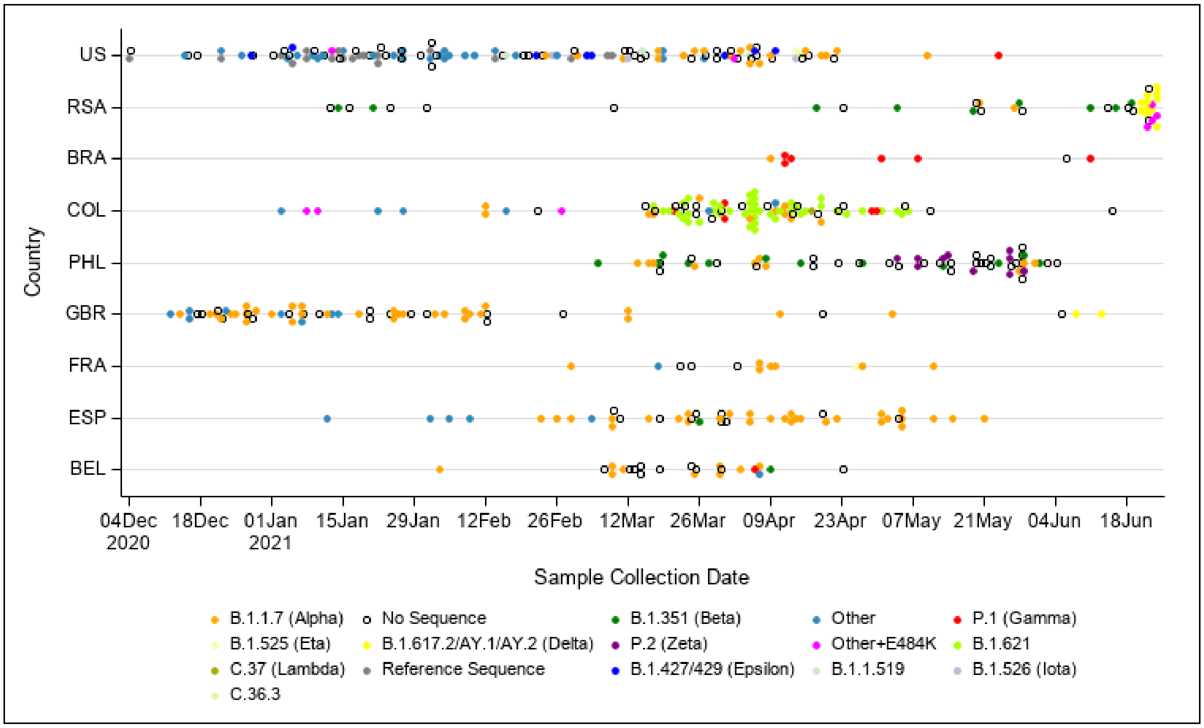

The distribution of SARS-CoV-2 variants is shown by individual country over time for the double-blind phase of the study. Case accrual by variants was performed where sequence data were available. The reference sequence is defined as the SARS-CoV-2 Wuhan-Hu1 sequence but with the addition of the amino acid variation D614G. Next-Generation Sequencing was performed using the Swift Biosciences SNAP Assay (v2) using 1% and baseline polymorphisms defined with a cut-off of 15%. Amino acid variants are defined as changes from the reference sequence. The last unblinding visit was June 21-25, 2021, except Germany (May 25, 2021). SARS-CoV-2, severe acute respiratory syndrome coronavirus-2; US, United States; RSA, Republic of South Africa; BRA, Brazil; COL, Colombia; PHL, Philippines; GBR, United Kingdom; FRA, France; ESP, Spain; BEL, Belgium.

**Figure S4. Vaccine Efficacy against Moderate to Severe–Critical Covid-19 with Onset at Least 14 Days after Booster Vaccination by Virus Variant (Per-protocol Set)**

"Other" refers to all strains not considered variants based on their mutations. "Variant substitution" refers to all variants combined, except for Other and the reference strain. Participants were seronegative at baseline and through day 70, as determined by PCR and serological tests. 95% CIs for vaccine efficacy against moderate to severe–critical Covid-19 (primary endpoint) are adjusted, implementing type I error control for multiple testing and presented upon meeting the prespecified testing conditions. Except as indicated by **, all others were descriptive 95% CI, with no adjustments made for multiple testing. If fewer than 6 cases were observed for an endpoint, then the VE will not be shown. [a]The risk set excluded participants who had a Covid-19 case or discontinued before day 70. [b]Moderate to severe–critical Covid-19 cases. CI, confidence interval; PP, per-protocol; PY, person-years; VE, vaccine efficacy; vp, viral particles.

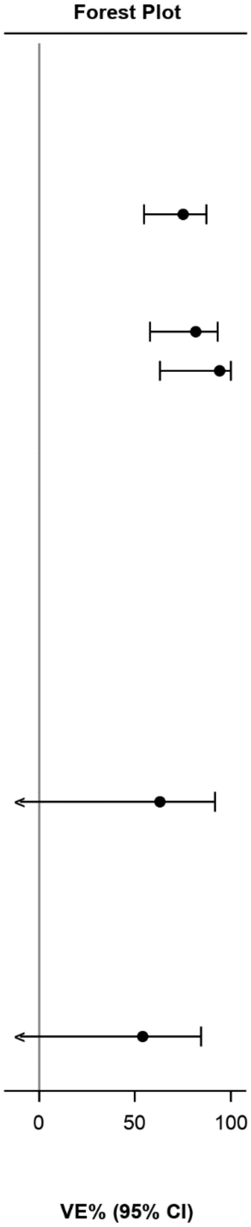

| | Ad26 5e10 vp | | Placebo | | |
|---|---|---|---|---|---|
| | #Cases(N) | PY | #Cases(N) | PY | VE% (95% CI) |
| Analysis set: PP | (7484) | | (7008) | | |
| Risk set[a] | (6024) | | (5615) | | |
| Primary endpoint[b] | | | | | |
| All participants** | 14 (6024) | 1729.99 | 52 (5615) | 1594.98 | 75.2% (54.55; 87.30) |
| Reference strain | 0 (6024) | 1729.99 | 0 (5615) | 1594.98 | |
| Variant substitution | 7 (6024) | 1729.99 | 35 (5615) | 1594.98 | 81.6% (57.87; 93.09) |
| B.1.1.7 (Alpha) | 1 (6024) | 1729.99 | 16 (5615) | 1594.98 | 94.2% (62.91; 99.86) |
| B.1.351 (Beta) | 0 (6024) | 1729.99 | 3 (5615) | 1594.98 | |
| B.1.617.2/AY.1/AY.2 (Delta) | 2 (6024) | 1729.99 | 1 (5615) | 1594.98 | |
| B.1.427/429 (Epsilon) | 0 (6024) | 1729.99 | 0 (5615) | 1594.98 | |
| B.1.525 (Eta) | 0 (6024) | 1729.99 | 0 (5615) | 1594.98 | |
| P.1 (Gamma) | 0 (6024) | 1729.99 | 1 (5615) | 1594.98 | |
| B.1.526 (Iota) | 0 (6024) | 1729.99 | 2 (5615) | 1594.98 | |
| B.1.617.1 (Kappa) | 0 (6024) | 1729.99 | 0 (5615) | 1594.98 | |
| C.37 (Lambda) | 0 (6024) | 1729.99 | 1 (5615) | 1594.98 | |
| P.3 (Theta) | 0 (6024) | 1729.99 | 0 (5615) | 1594.98 | |
| P.2 (Zeta) | 0 (6024) | 1729.99 | 0 (5615) | 1594.98 | |
| B.1.621 | 4 (6024) | 1729.99 | 10 (5615) | 1594.98 | 63.1% (-27.86; 91.56) |
| C.36.3 | 0 (6024) | 1729.99 | 0 (5615) | 1594.98 | |
| R.1 | 0 (6024) | 1729.99 | 0 (5615) | 1594.98 | |
| B.1.1.519 | 0 (6024) | 1729.99 | 0 (5615) | 1594.98 | |
| Other+E484K | 0 (6024) | 1729.99 | 1 (5615) | 1594.98 | |
| Other | 0 (6024) | 1729.99 | 3 (5615) | 1594.98 | |
| No sequencing data available | 7 (6024) | 1729.99 | 14 (5615) | 1594.98 | 53.9% (-22.05; 84.25) |

**Figure S5. Vaccine Efficacy against Moderate to Severe–Critical Covid-19 with Onset at Least 14 Days after Booster Vaccination by Subgroups (Per-protocol Set)**

95% CIs for vaccine efficacy against moderate to severe–critical Covid-19 (primary endpoint) are adjusted, implementing type I error control for multiple testing and presented upon meeting the prespecified testing conditions. Except as indicated by **, all others were descriptive 95% CI, with no adjustments made for multiple testing. If fewer than 6 cases were observed for an endpoint, then the VE will not be shown. [a]The risk set excluded participants who had a Covid-19 case as determined by PCR and serological tests or discontinued before day 70. [b]Moderate to severe–critical Covid-19 cases. CI, confidence interval; PP, per-protocol; PY, person-years; VE, vaccine efficacy; vp, viral particles.

Forest Plot

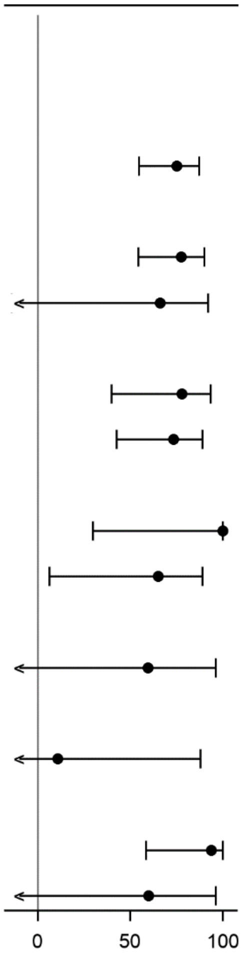

| | Ad26 5e10 vp | | Placebo | | |
|---|---|---|---|---|---|
| | #Cases(N) | PY | #Cases(N) | PY | VE% (95% CI) |
| Analysis set: PP | (7484) | | (7008) | | |
| Risk set[a] | (6024) | | (5615) | | |
| Primary endpoint[b] | | | | | |
| All participants** | 14 (6024) | 1729.99 | 52 (5615) | 1594.98 | 75.2% (54.55; 87.30) |
| By Age Group I | | | | | |
| 18-59 years | 10 (4692) | 1386.93 | 41 (4359) | 1276.36 | 77.6% (54.44; 89.97) |
| >=60 years | 4 (1332) | 343.06 | 11 (1256) | 318.61 | 66.2% (-13.97; 92.16) |
| By Comorbidities | | | | | |
| Yes | 5 (2116) | 592.77 | 21 (2008) | 550.31 | 77.9% (39.77; 93.49) |
| No | 9 (3908) | 1137.23 | 31 (3607) | 1044.67 | 73.3% (42.61; 88.83) |
| By Country | | | | | |
| Belgium | 0 (705) | 179.50 | 7 (721) | 181.61 | 100.0% (29.80; 100.00) |
| Colombia | 6 (280) | 84.97 | 16 (269) | 78.83 | 65.2% (6.40; 88.85) |
| Germany | 0 (44) | 10.89 | 0 (15) | 3.40 | |
| Spain | 2 (603) | 154.01 | 5 (614) | 156.28 | 59.4% (-147.93; 96.13) |
| France | 0 (79) | 20.26 | 0 (76) | 19.05 | |
| Great Britain | 3 (1535) | 468.68 | 3 (1387) | 418.90 | 10.6% (-567.31; 88.03) |
| Philippines | 0 (139) | 38.30 | 2 (134) | 36.07 | |
| United States | 1 (2232) | 632.40 | 14 (1999) | 559.74 | 93.7% (58.45; 99.85) |
| South Africa | 2 (407) | 140.98 | 5 (400) | 141.10 | 60.0% (-144.53; 96.19) |

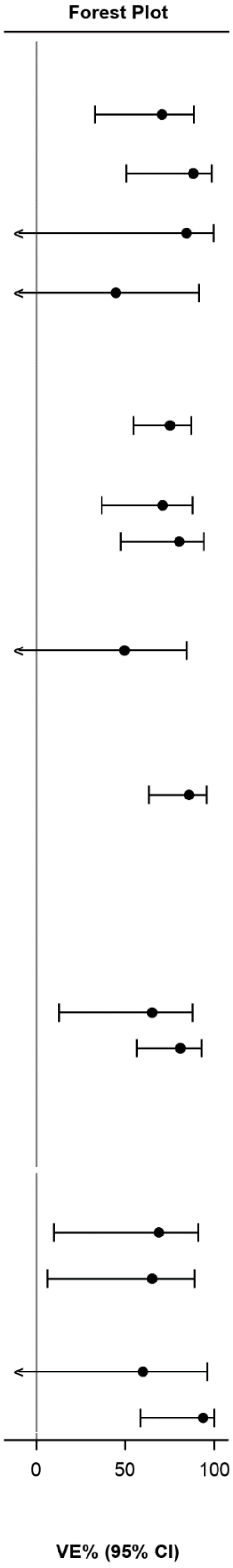

| | Ad26 5e10 vp | | Placebo | | |
|---|---|---|---|---|---|
| | #Cases(N) | PY | #Cases(N) | PY | VE% (95% CI) |
| By Age and comorbidities 1 | | | | | |
| 18-59, no | 8 (3274) | 977.16 | 25 (3015) | 894.45 | 70.7% (33.01; 88.58) |
| 18-59, yes | 2 (1418) | 409.78 | 16 (1344) | 381.91 | 88.4% (50.45; 98.70) |
| >=60, no | 1 (634) | 160.07 | 6 (592) | 150.21 | 84.4% (-28.91; 99.66) |
| >=60, yes | 3 (698) | 182.99 | 5 (664) | 168.40 | 44.8% (-183.80; 91.43) |
| By HIV Infection | | | | | |
| Yes | 0 (72) | 19.45 | 0 (66) | 17.73 | |
| No | 14 (5952) | 1710.54 | 52 (5549) | 1577.25 | 75.2% (54.54; 87.29) |
| By Sex | | | | | |
| Male | 9 (3320) | 958.29 | 29 (3139) | 900.45 | 70.8% (36.72; 87.86) |
| Female | 5 (2702) | 771.24 | 23 (2474) | 693.91 | 80.4% (47.40; 94.19) |
| Undifferentiated | 0 (2) | 0.47 | 0 (2) | 0.61 | |
| By Race | | | | | |
| Amer. Indian/Alaskan | 6 (151) | 48.47 | 12 (158) | 48.91 | 49.6% (-45.24; 84.46) |
| Asian | 0 (327) | 96.42 | 5 (301) | 88.82 | |
| Black or African Amer. | 1 (352) | 122.40 | 2 (383) | 133.46 | |
| Native Hawaiian/other | 0 (7) | 2.46 | 0 (14) | 3.92 | |
| White | 5 (5014) | 1407.78 | 32 (4608) | 1277.23 | 85.8% (63.35; 95.69) |
| Multiple | 0 (66) | 20.15 | 1 (51) | 14.80 | |
| Unknown | 2 (35) | 10.41 | 0 (28) | 7.52 | |
| Not Reported | 0 (57) | 18.06 | 0 (55) | 16.39 | |
| Missing | 0 (15) | 3.84 | 0 (17) | 3.90 | |
| By Ethnicity | | | | | |
| Hispanic/Latino | 7 (729) | 211.20 | 18 (677) | 189.00 | 65.2% (12.68; 87.71) |
| Not Hispanic/Latino | 7 (5132) | 1466.74 | 34 (4776) | 1356.41 | 81.0% (56.37; 92.88) |
| Not Reported | 0 (122) | 39.66 | 0 (112) | 35.59 | |
| Unknown | 0 (28) | 9.19 | 0 (33) | 10.20 | |
| Missing | 0 (13) | 3.21 | 0 (17) | 3.78 | |
| By Region | | | | | |
| Europe | 5 (2966) | 833.34 | 15 (2813) | 779.24 | 68.8% (9.78; 91.14) |
| Latin America | 6 (280) | 84.97 | 16 (269) | 78.83 | 65.2% (6.40; 88.85) |
| Philippines | 0 (139) | 38.30 | 2 (134) | 36.07 | |
| South Africa | 2 (407) | 140.98 | 5 (400) | 141.10 | 60.0% (-144.53; 96.19) |
| United States | 1 (2232) | 632.40 | 14 (1999) | 559.74 | 93.7% (58.45; 99.85) |

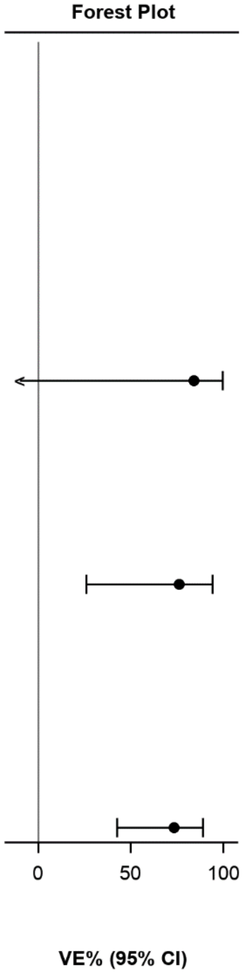

| | Ad26 5e10 vp | | Placebo | | |
|---|---|---|---|---|---|
| | #Cases(N) | PY | #Cases(N) | PY | VE% (95% CI) |
| By Comorbidities categories 2 | | | | | |
| Asthma | 0 (94) | 25.45 | 1 (84) | 21.80 | |
| Cancer | 0 (19) | 4.80 | 0 (19) | 4.74 | |
| Cerebrovascular Disease | 0 (15) | 4.37 | 0 (19) | 5.34 | |
| Cystic Fibrosis | 0 (0) | - | 0 (0) | - | |
| Chronic Kidney Disease | 0 (15) | 4.55 | 0 (13) | 3.36 | |
| COPD | 0 (53) | 14.69 | 1 (58) | 14.57 | |
| Serious heart conditions | 0 (202) | 52.43 | 2 (169) | 43.41 | |
| Hypertension | 1 (515) | 136.12 | 6 (501) | 129.57 | 84.1% (-30.76; 99.66) |
| ICP from blood transpl. | 0 (5) | 1.56 | 0 (6) | 1.67 | |
| ICP from organ transpl. | 0 (0) | - | 0 (0) | - | |
| Liver Disease | 0 (21) | 5.80 | 0 (16) | 4.32 | |
| Neurologic conditions | 0 (25) | 6.70 | 0 (22) | 5.67 | |
| Obesity | 4 (1434) | 410.47 | 16 (1394) | 391.80 | 76.1% (26.03; 94.19) |
| Pulmonary Fibrosis | 0 (2) | 0.48 | 0 (2) | 0.48 | |
| Sickle Cell Disease | 0 (1) | 0.47 | 0 (1) | 0.26 | |
| Type 1 Diabetes Mellitus | 0 (23) | 6.18 | 0 (17) | 4.20 | |
| Type 2 Diabetes Mellitus | 0 (301) | 84.45 | 1 (252) | 70.08 | |
| Thalassemia | 1 (6) | 1.54 | 0 (6) | 1.65 | |
| None | 9 (3908) | 1137.23 | 31 (3607) | 1044.67 | 73.3% (42.61; 88.83) |

**Figure S6. Number of Symptomatic Covid-19 Cases with Onset at Least 14 Days after Booster Vaccination by Number of Signs and Symptoms (A) and by Severity (B and C; Per-protocol Set)**

Covid-19, coronavirus disease 2019; vp, viral particles.

A)

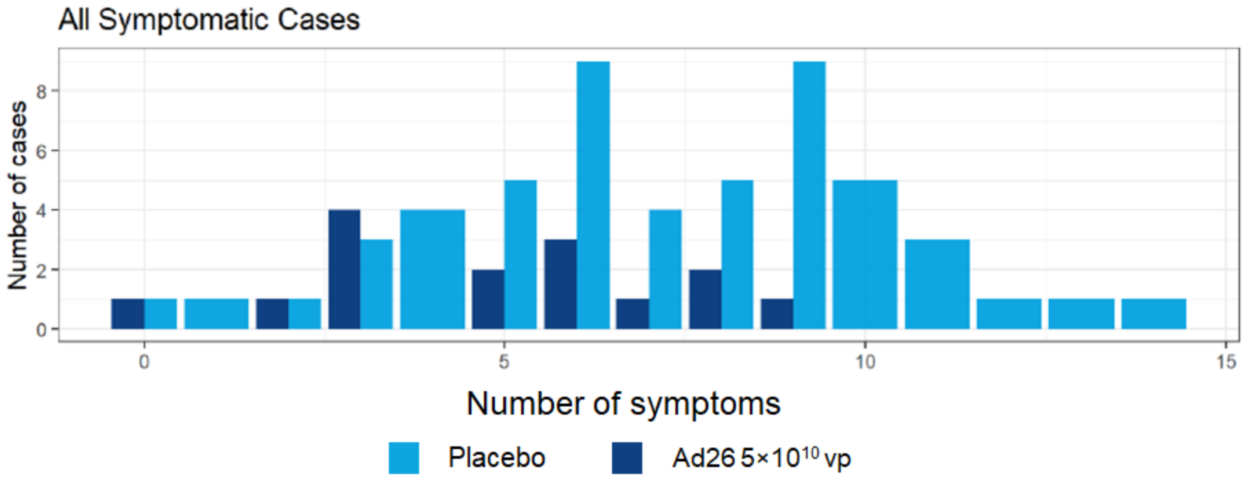

B)

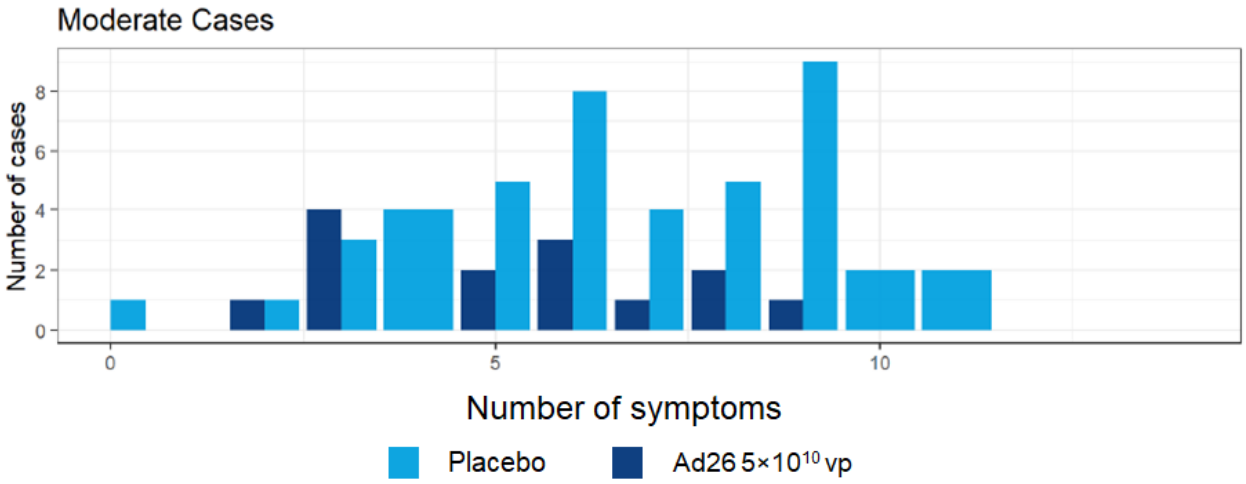

C)

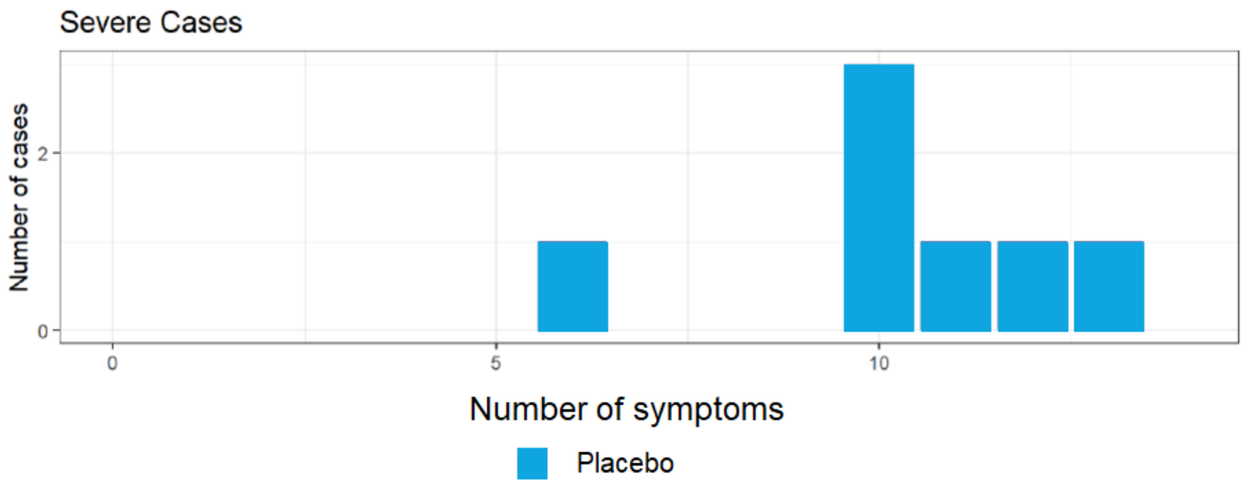
Severe Cases
Number of cases
2
0
0
5
10
Number of symptoms
Placebo

**Figure S7. Cases of Moderate to Severe–Critical Covid-19 with Onset at Least 28 Days after Booster Vaccination by Covid-19 Duration.**

Fewer overall cases of Covid-19 lasting >28 days were reported for the Ad26.COV2.S group than for the placebo group (post-hoc analysis 5/13 [38.5%] vs 28/50 [56.0%]. Ad26 5e10 vp=Ad26.COV2.S at a dose level of $5 \times 10^{10}$ viral particles.

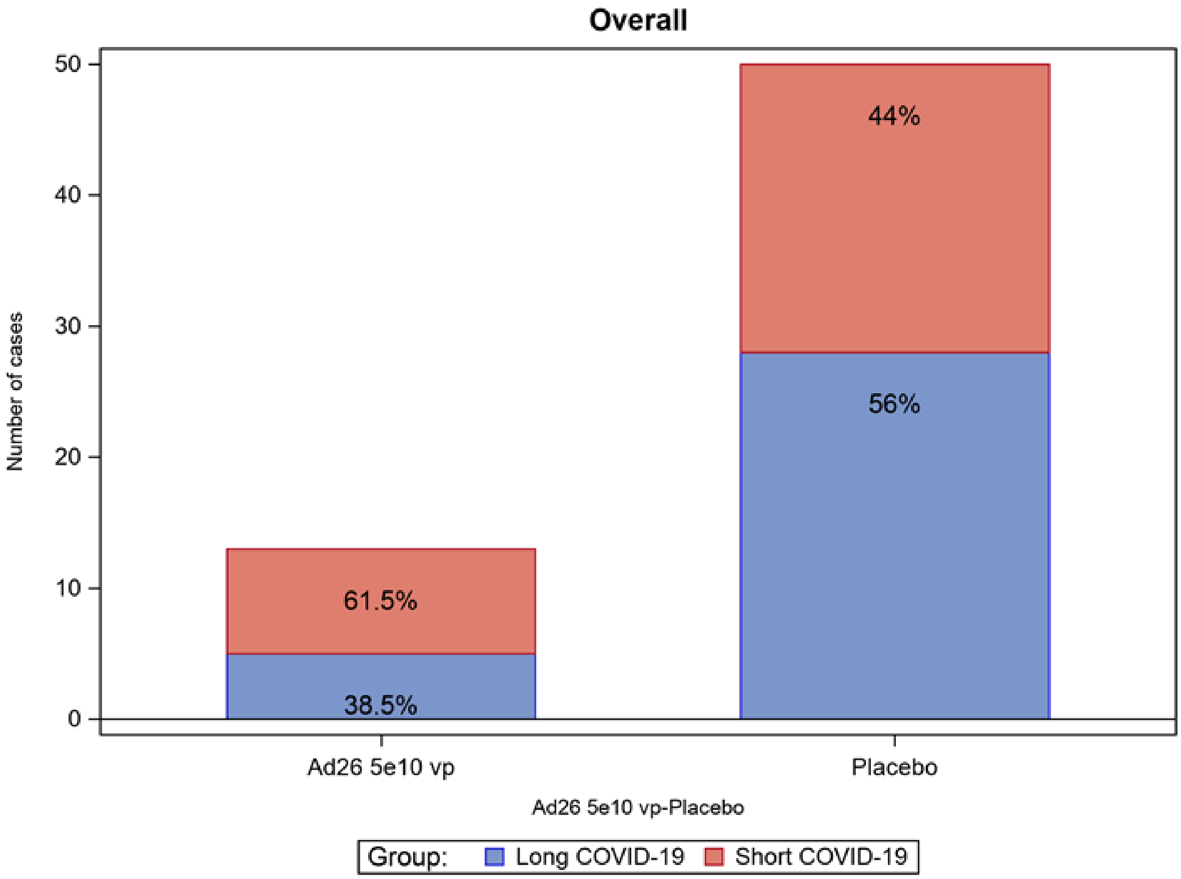

**Figure S8. Concentration of SARS-CoV-2 S Protein Binding Antibodies Over Time (Per-protocol Immunogenicity Set)**

In the immunogenicity subset, the spike-specific binding antibody geometric mean concentration (GMC) was 367 (95% CI, 295-456) 28 days after the first dose Ad26.COV2.S (day 29), increasing to 518 (95% CI, 422-635) before boosting, and to 2220 (95% CI, 1794-2748) 14 days post-booster (day 71). The geometric mean increases in spike-specific binding antibody concentrations were 7.2- and 40.5-fold from baseline to day 29 and day 71, respectively, and 4.7-fold from day 57 (pre-booster) to day 71. Following a single vaccination, responder rates were 91.9% by day 29; after boosting, response rates reached 100% by day 71. ELISA, enzyme-linked immunosorbent assay; GMC, geometric mean concentration; LLOQ, lower limit of quantification; SARS-CoV-2, severe acute respiratory syndrome coronavirus 2; ULOQ, upper limit of quantification.

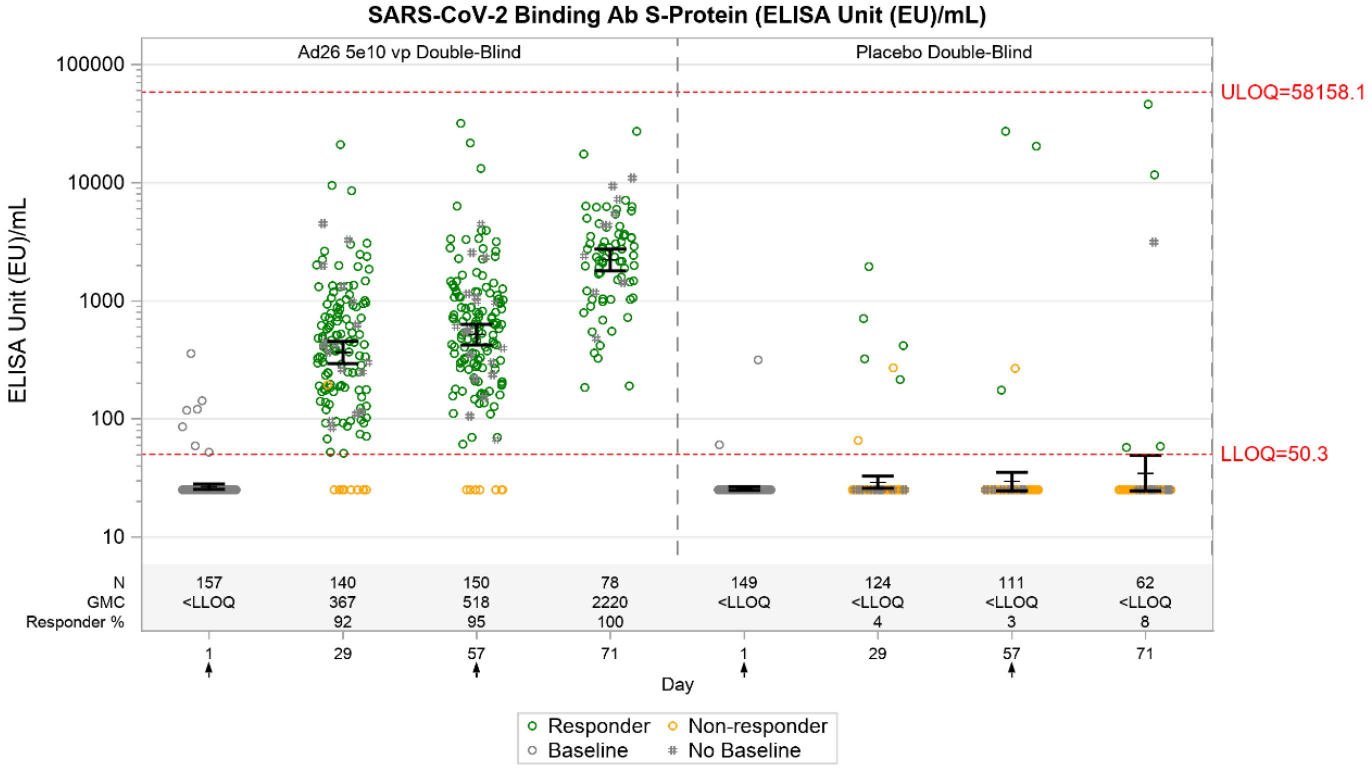

**Figure S9. Solicited Local (A and B) and Systemic (C and D) Adverse Events Following Prime-Boost Vaccination Regimen in Adults Aged 18 to 59 Years and ≥60 Years (Safety Set)**

AE, adverse event; vp, viral particles.

A) Post-primary dose solicited local AEs among those aged 18–59 years and ≥60 years

**18–59 y**

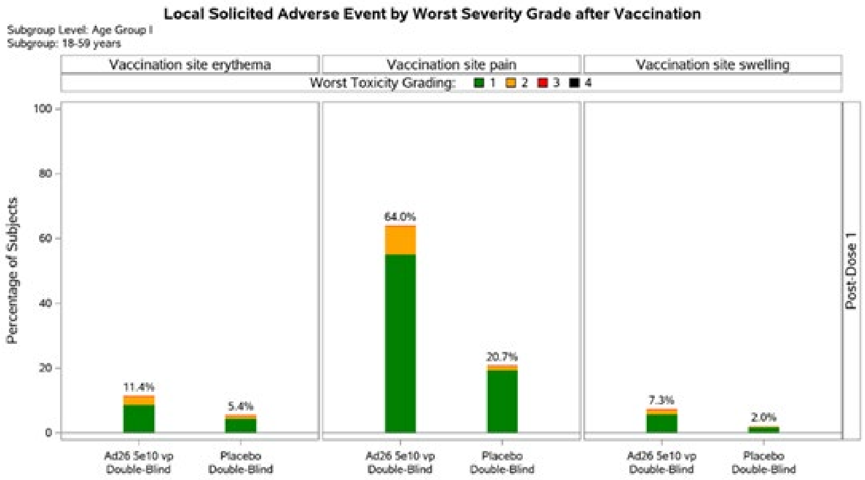

**≥60 y**

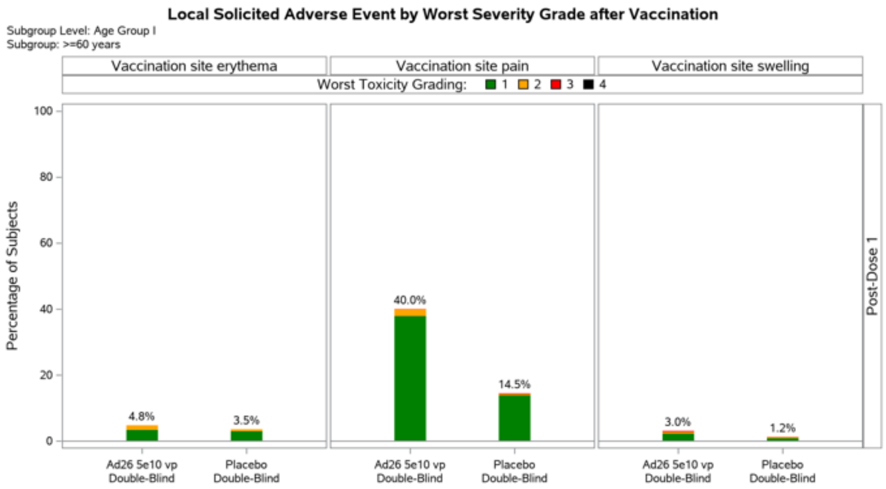

B) Post-booster dose solicited local AEs among those aged 18–59 years and ≥60 years

**18–59 y**

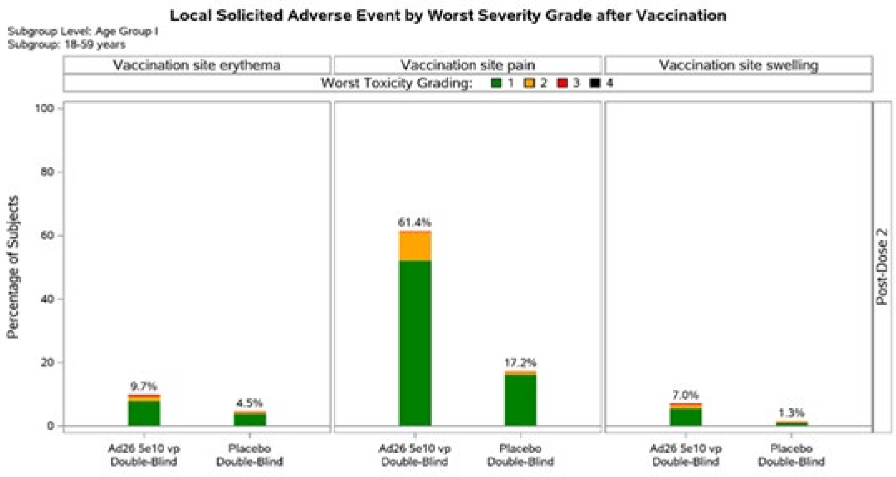

**≥60 y**

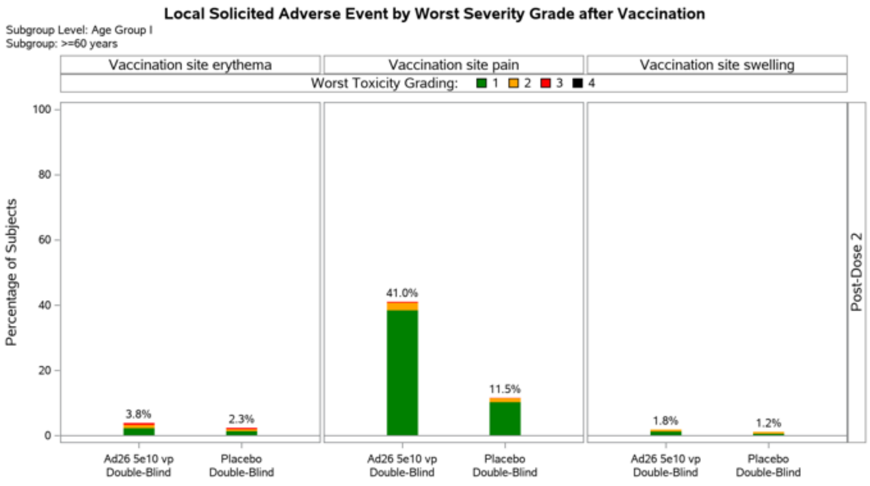

C) Post-primary dose solicited systemic AEs among those aged 18–59 years and ≥60 years

**18–59 y**

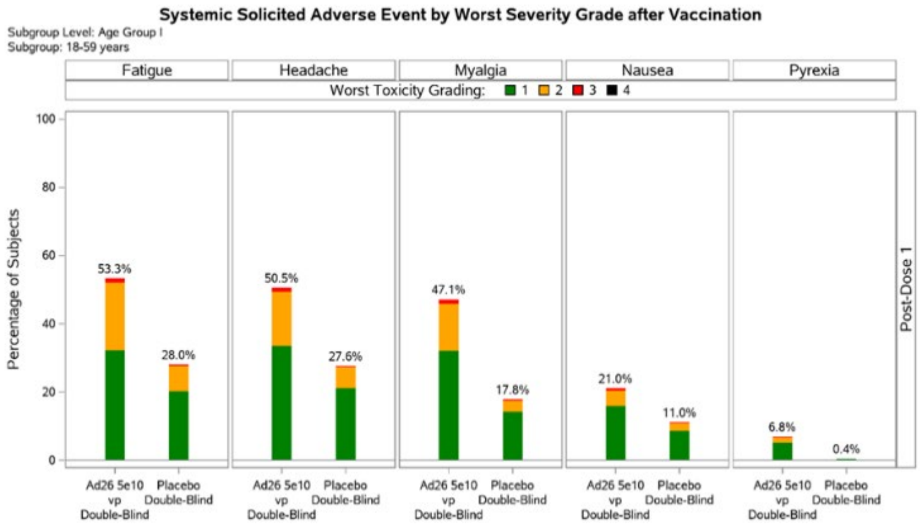

**≥60 y**

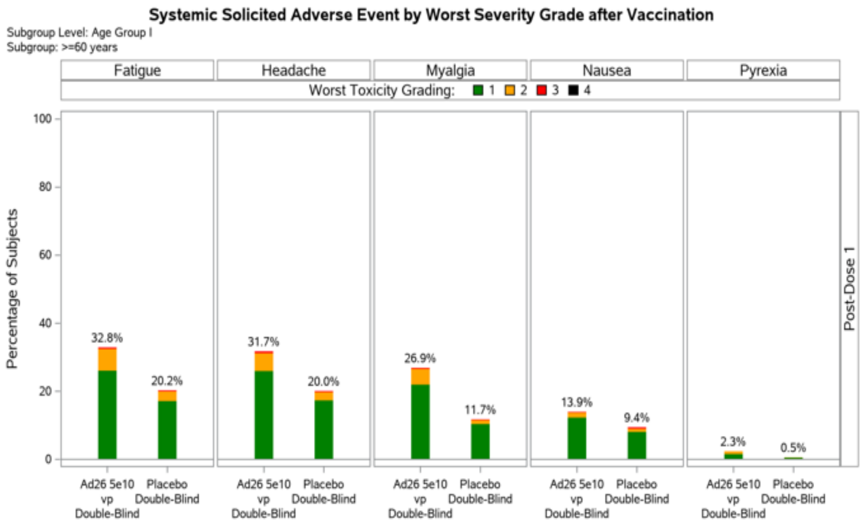

D) Post-booster dose solicited systemic AEs among those aged 18–59 years and ≥60 years

**18–59 y**

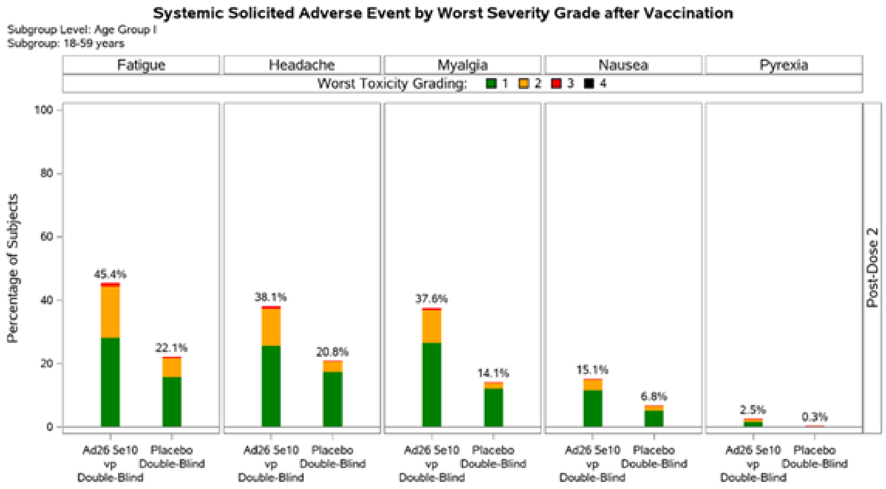

**≥60 y**

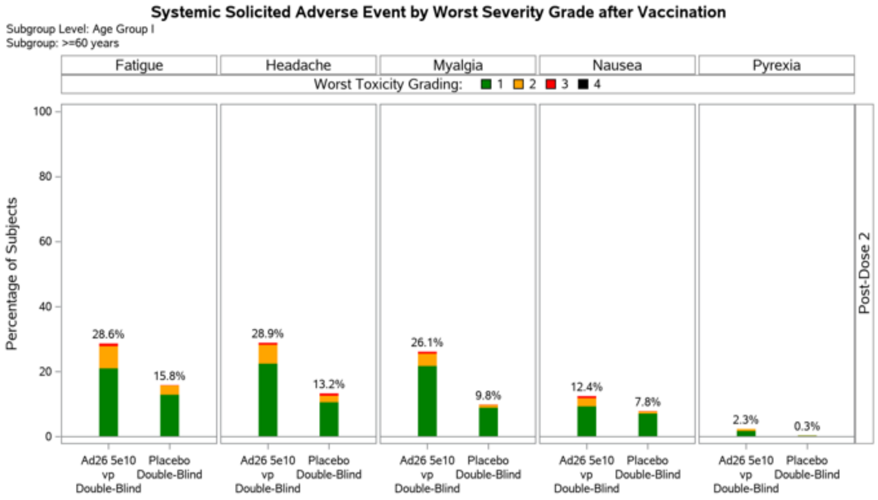

### Supplementary Tables

#### Table S1. Objectives and Endpoints of the Double-Blind Phase of the Trial

| Objectives | Endpoints |
|---|---|
| **Primary** | |
| To demonstrate the efficacy of Ad26.COV2.S in the prevention of molecularly-confirmed[a], moderate to severe/critical Covid-19[b], as compared to placebo, in SARS-CoV-2 seronegative adults | First occurrence of molecularly-confirmed[a], moderate to severe/critical Covid-19[b], with onset at least 14 days after the second vaccination (Day 71). |
| **Secondary**[f]<br>*(The method used to perform hypothesis testing preserving the family-wise error rate [FWER] will be specified in the Statistical Analysis Plan [SAP])* | |
| *Efficacy* | |
| To demonstrate the efficacy of Ad26.COV2.S in the prevention of molecularly-confirmed[a], moderate to severe/critical Covid-19[b], as compared to placebo, in adults regardless of their serostatus | • First occurrence of molecularly-confirmed[a], moderate to severe/critical Covid-19[b], with onset 1 day after the 1st vaccination<br>• First occurrence of molecularly-confirmed[a], moderate to severe/critical Covid-19[b], with onset 14 days after the second vaccination (Day 71) |
| To evaluate the efficacy of Ad26.COV2.S in the prevention of molecularly-confirmed[a], moderate to severe/critical Covid-19[b] as compared to placebo | • First occurrence of molecularly-confirmed[a], moderate to severe/critical Covid-19[b] with onset 1 day after the 1st vaccination<br>• First occurrence of molecularly-confirmed[a], moderate to severe/critical Covid-19[b] with onset 14 days after the 1st vaccination (Day 15)<br>• First occurrence of molecularly-confirmed[a], moderate to severe/critical Covid-19[b] with onset 28 days after the 1st vaccination (Day 29). |
| To assess the effect of Ad26.COV2.S on Covid-19 requiring medical intervention (based on objective criteria) compared to placebo | First occurrence of Covid-19 requiring medical intervention (such as a composite endpoint of hospitalization, ICU admission, mechanical ventilation, and ECMO, linked to objective measures such as decreased oxygenation, X-ray or CT findings) and linked to any molecularly-confirmed[a], |

| Objectives | Endpoints |
| --- | --- |
| | Covid-19[b,c] at least 14 days after the second vaccination (Day 71) |
| To assess the effect of Ad26.COV2.S on SARS-CoV-2 viral RNA load compared to placebo for moderate to severe/critical Covid-19[b] | Assessment of the SARS-CoV-2 viral load by quantitative RT-PCR, in participants with molecularly-confirmed[a], moderate to severe/critical Covid-19[b] by serial viral load measurements during the course of a Covid-19 episode, at least 14 days after the second vaccination (Day 71) |
| To assess the effect of Ad26.COV2.S on molecularly-confirmed[a], mild Covid-19[c] | First occurrence of molecularly-confirmed[a], mild Covid-19[c], at least 14 days after the second vaccination (Day 71) |
| To assess the effect of Ad26.COV2.S on Covid-19 as defined by the US FDA harmonized case definition[d] | First occurrence of molecularly-confirmed[a] Covid-19[d] at least 14 days after the second vaccination (Day 71) |
| To assess the effect of Ad26.COV2.S on all molecularly-confirmed[a] symptomatic Covid-19[b,c], as compared to placebo | Burden of disease (BOD) endpoint (see Section 9.5.2) derived from the first occurrence of molecularly-confirmed[a] symptomatic Covid-19[b,c] (meeting the mild, moderate or severe/critical Covid-19 case definition) with onset at least 14 days after the second vaccination (Day 71) |
| To assess the effect of Ad26.COV2.S on occurrence of confirmed asymptomatic or undetected infections with SARS-CoV-2, as compared to placebo[e] | • Serologic conversion between baseline and other blood samples before unblinding visit using an enzyme-linked immunosorbent assay (ELISA) and/or SARS-CoV-2 immunoglobulin assay that is dependent on the SARS-CoV-2 nucleocapsid (N) protein<br>• Asymptomatic infection detected by RT-PCR |
| To assess the efficacy of Ad26.COV2.S in the prevention of SARS-CoV-2 infection (both symptomatic and asymptomatic infections combined, that are serologically and/or molecularly-confirmed[a]), as compared to placebo | First occurrence of SARS-CoV-2 infection (serologically and/or molecularly-confirmed[a]) with onset at least 14 days after the second vaccination (Day 71) |
| *Safety* | |
| To evaluate safety in terms of SAEs and AESIs (during the entire study), MAAEs (until 6 months after the last double-blind vaccination), and MAAEs leading to study discontinuation (during the entire study) for all participants | Occurrence and relationship of SAEs and AESIs (during the entire study), MAAEs (until 6 months after the last double-blind vaccination), and MAAEs leading to study discontinuation (during the entire study) for all participants |

| Objectives | Endpoints |
|---|---|
| In a subset of participants, to evaluate the safety and reactogenicity in terms of solicited local and systemic AEs during 7 days after each vaccination, and in terms of unsolicited AEs during 28 days after each vaccination | Occurrence, intensity, duration and relationship of solicited local and systemic AEs during 7 days following each vaccination and of unsolicited AEs during 28 days following each vaccination |
| *Immunogenicity* | |
| In a subset of participants, to evaluate the immunogenicity of Ad26.COV2.S, as compared to placebo | Analysis of antibodies binding to the SARS-CoV-2 S protein by ELISA. |
| **Exploratory** | |
| To assess the effect of Ad26.COV2.S on SARS-CoV-2 viral RNA load compared to placebo for mild Covid-19[c] | Assessment of the SARS-CoV-2 viral load by quantitative RT-PCR, in participants with molecularly-confirmed[a], mild Covid-19[c] by serial viral load measurements during the course of a Covid-19 episode |
| To assess the effect of Ad26.COV2.S on health care utilization (such as hospitalization, ICU admission, ventilator use) linked to any molecularly-confirmed[a] Covid-19, as compared to placebo | Health care utilization (such as hospitalization, ICU admission, ventilator use) linked to any molecularly-confirmed[a] Covid-19 at least 14 days after the second vaccination (Day 71) |
| To assess the efficacy of Ad26.COV2.S in the prevention of SARS-CoV-2 infection in participants with comorbidities associated with increased risk of progression to severe Covid-19, as compared to placebo | First occurrence of SARS-COV-2 infection (serologically and/or molecularly-confirmed[a]) in participants with comorbidities associated with increased risk of progression to severe Covid-19 with onset at least 14 days after the second vaccination (Day 71) |
| To explore the effect of Ad26.COV2.S on other potential complications of Covid-19 (linked to any respiratory disease and linked to any molecularly-confirmed[a] Covid-19) not previously described, as compared to placebo | First occurrence of potential complications of Covid-19 linked to any respiratory disease and linked to any molecularly-confirmed[a] Covid-19, with onset at least 14 days after the second vaccination (Day 71) |
| To explore the effect of Ad26.COV2.S on all-cause mortality, as compared to placebo | Deaths occurring at least 14 days after the second vaccination (Day 71) |
| To evaluate the immune response in participants with Covid-19 in relation to risk of development of Covid-19, protection induced by Ad26.COV2.S, and risk of accelerated disease | Assessment of the correlation of humoral immune responses with emphasis on neutralizing, binding and functional antibodies, with the risk of Covid-19 and protection induced by the study vaccine |
| In a subset of participants to further assess the humoral immune response to Ad26.COV2.S, as compared to placebo | Humoral immunogenicity endpoints:<br>• Functional and molecular antibody characterization including, but not limited to avidity, Fc-mediated viral clearance, Fc characteristics, Ig subclass, IgG isotype, antibody glycosylation, and assessment of antibody repertoire |

| Objectives | Endpoints |
|---|---|
| | • Original and/or emerging SARS-CoV-2 lineage neutralization as measured by virus neutralization assay (VNA; wild-type virus and/or pseudovirion expressing SARS-CoV-2 S protein)<br>• Adenovirus neutralization as measured by VNA<br>• Analysis of antibodies to the receptor-binding domain (RBD) of the SARS-CoV-2 S protein<br>• Passive transfer: analysis of immune mediators correlating with protection against experimental SARS-CoV-2 challenge in a suitable animal model. |
| To explore changes in the SARS-CoV-2 genome | Development of SARS-CoV-2 variants |
| To examine efficacy for moderate/severe and severe disease as well as medical utilization or death in the vaccine and placebo groups for variant strains that have been identified or if there are cases associated with a combination of variants. | Occurrence of moderate/severe or severe Covid-19, medical utilization, or death for each of the circulating viral variants, identified by S-gene sequencing |
| To evaluate patient-reported outcomes (PROs) in relation to the presence of SARS-CoV-2 infection and the presence, severity and duration of Covid-19 signs and symptoms in participants who received Ad26.COV2.S, as compared to placebo | • Presence, severity and duration of Covid-19 signs and Symptoms;<br>• Confirmation of SARS-CoV-2 infection by molecular testing |
| To assess the difference in severity of cases in participants who received Ad26.COV2.S as compared to placebo | Reduction in severity of Covid-19 signs and Symptoms |
| To assess the impact of pre-existing humoral immunity against coronaviruses other than SARS-CoV-2 at baseline on Ad26.COV2.S vaccine immunogenicity | Analysis of antibodies binding to coronaviruses other than SARS-CoV-2 by ELISA |
| To assess the incidence of co-infection of SARS-CoV-2 and other respiratory pathogens and to assess the effect of the vaccine during such co-infections as well as to estimate the incidence of other respiratory pathogens during the study period. | Analysis of broad respiratory pathogens panel in the nasal swabs collected during a confirmed Covid-19 episode and in a subset of nasal swab samples from participants with a symptomatic infection. |
| In US participants, To increase the information on prior medical history (electronic health records, claims, laboratory data from other care settings) in order to further evaluate its potential effect on the response to immunization and the impact of immunization on efficacy and duration of efficacy as well as AEs that may occur during and after completion of the study. | Utilization of tokenization and matching procedures for exploratory analysis of participant's medical data prior to, during, and following participation in the study (real-world data). Analysis will be performed to relate real-world data to vaccine immune responses, efficacy and duration of protection, and AEs (see Section 4.1 and Section 8.7). |

[a] Molecularly-confirmed Covid-19 is defined as a positive SARS-CoV-2 viral RNA result by a central laboratory using a RT-PCR-based or other molecular diagnostic test .
[b] Per case definition for moderate to severe/critical Covid-19 as determined by the Clinical Severity Adjudication Committee.
[c] Per case definition for mild Covid-19 (see Supplementary Methods) as determined by the Clinical Severity Adjudication Committee.
[d] Per case definition for Covid-19 according to the US FDA harmonized case definition.
[e] Per case definition for asymptomatic or undetected Covid-19 as determined by the Clinical Severity Adjudication Committee.
[f] All secondary efficacy endpoint analyses will occur in the PP analysis set, in seronegative participants unless otherwise indicated in the statistical analysis plan.

**Table S2. Baseline Characteristics of the Trial Participants (Per-protocol Set*)**

| Characteristic | Ad26.COV2.S (N=7,484) | Placebo (N=7.008) | Total (N=14,492) |
|---|---|---|---|
| Age – no. (%) | 7484 | 7008 | 14,492 |
| 18-59 yr | 5590 (74.7) | 5233 (74.7) | 10,823 (74.7) |
| ≥60 yr | 1894 (25.3) | 1775 (25.3) | 3669 (25.3) |
| Age − median (range), yr | 50 (18-99) | 50 (18-92) | 50 (18-99) |
| Sex – no. (%) | 7484 | 7008 | 14,492 |
| Female | 3401 (45.4) | 3126 (44.6) | 6527 (45.0) |
| Male | 4081 (54.5) | 3879 (55.4) | 7960 (54.9) |
| Undifferentiated | 2 (<0.1) | 3 (<0.1) | 5 (<0.1) |
| Race† – no. (%) | 7484 | 7008 | 14492 |
| American Indian or Alaskan Native‡ | 166 (2.2) | 174 (2.5) | 340 (2.3) |
| Asian | 373 (5.0) | 339 (4.8) | 712 (4.9) |
| Black or African American | 382 (5.1) | 402 (5.7) | 784 (5.4) |
| Native Hawaiian or other Pacific Islander | 10 (0.1) | 16 (0.2) | 26 (0.2) |
| White | 6349 (84.8) | 5886 (84.0) | 12,235 (84.4) |
| Multiracial | 72 (1.0) | 63 (0.9) | 135 (0.9) |
| Not reported, unknown or missing | 132 (1.8) | 128 (1.8) | 260 (1.8) |
| Ethnicity† − no. (%) | 7484 | 7008 | 14,492 |
| Hispanic or Latino | 874 (11.7) | 822 (11.7) | 1696 (11.7) |
| Not Hispanic or Latino | 6386 (85.3) | 5969 (85.2) | 12,355 (85.3) |
| Not reported, unknown or missing | 224 (3.0) | 217 (3.1) | 441 (3.0) |
| Country/region – no. (%) | 7484 | 7008 | 14,492 |
| Europe | 3814 (51.0) | 3679 (52.5) | 7493 (51.7) |
| Belgium | 751 (10.0) | 766 (10.9) | 1517 (10.5) |
| France | 96 (1.3) | 89 (1.3) | 185 (1.3) |
| Germany | 47 (0.6) | 46 (0.7) | 93 (0.6) |
| Spain | 692 (9.2) | 710 (10.1) | 1402 (9.7) |
| United Kingdom | 2228 (29.8) | 2068 (29.5) | 4296 (29.6) |
| Latin America | 313 (4.2) | 307 (4.4) | 620 (4.3) |
| Brazil | 0 | 0 | 0 |
| Colombia | 313 (4.2) | 307 (4.4) | 620 (4.3) |
| The Philippines | 142 (1.9) | 137 (2.0) | 279 (1.9) |
| South Africa | 407 (5.4) | 403 (5.8) | 810 (5.6) |

| | | | |
|---|---|---|---|
| United States | 2808 (37.5) | 2482 (35.4) | 5290 (36.5) |
| SARS-CoV-2 serostatus – no. (%) | 7484 | 7008 | 14,492 |
| Positive | 0 | 0 | 0 |
| Negative | 7441 (99.4) | 6971 (99.5) | 14,412 (99.4) |
| Missing | 43 (0.6) | 37 (0.5) | 80 (0.6) |
| Body-mass index§ | 7474 | 7001 | 14,475 |
| Median | 26.30 | 26.50 | 26.40 |
| ≥30 – no. (%) | 1867 (25.0) | 1748 (25.0) | 3615 (25.0) |
| Baseline comorbidity – no. (%) | 7484 | 7008 | 14,492 |
| One or more | 2747 (36.7) | 2537 (36.2) | 5284 (36.5) |

*The per-protocol set included participants in the FAS who received 2 doses of vaccine or placebo in the double-blind phase, were PCR negative or missing a PCR test result at the time of first vaccination (day 1), were seronegative or missing by serology at the time of first vaccination and at 14 days after the second vaccination (day 71), and who had no major protocol deviations before unblinding that were judged to possibly impact the efficacy of the vaccine.

†Race and ethnicity were self-reported by participants.

‡Participants responding "Yes" to American Indian or Alaska Native in the Ad26.COV2.S group were from Colombia (n=148), the United States (n=13), Spain (n=4), and the United Kingdom (n=1); in the placebo group, participants were from Colombia (n=153), the United States (n=13), Spain (n=3), the United Kingdom (n=4), and South Africa (n=1).

§Body-mass index (BMI) is the weight in kilograms divided by the square of the height in meters. A BMI equal to or greater than 30 indicates obesity.

SARS-CoV-2, severe acute respiratory coronavirus 2.

**Table S3. Unsolicited Adverse Events Leading to Study Withdrawal or Study Treatment Discontinuation, Double-blind Phase (Full Analysis Set)**

| | Ad26.COV2.S | Placebo |
|---|---|---|
| **Post-Dose 1** | (N=15,705) | (N=15,588) |
| **AE, n (%)*** | | |
| Participants with ≥1 AEs leading to study withdrawal | 2 (<0.1) | 8 (0.1) |
| Injury, poisoning and procedural complications | 1 (<0.1) | 0 |
| Cervical vertebral fracture | 1 (<0.1) | 0 |
| Skin and subcutaneous tissue disorders | 1 (<0.1) | 0 |
| Urticaria | 1 (<0.1) | 0 |
| Cardiac disorders | 0 | 2 (<0.1) |
| Cardiac arrest | 0 | 1 (<0.1) |
| Ventricular extrasystoles | 0 | 1 (<0.1) |
| Infections and infestations | 0 | 5 (<0.1) |
| Covid-19 | 0 | 4 (<0.1) |
| Covid-19 pneumonia | 0 | 1 (<0.1) |
| Disseminated tuberculosis | 0 | 1 (<0.1) |
| Urinary tract infection | 0 | 1 (<0.1) |
| Metabolism and nutrition disorders | 0 | 1 (<0.1) |
| Diabetic ketoacidosis | 0 | 1 (<0.1) |
| Social circumstances | 0 | 1 (<0.1) |
| Homicide | 0 | 1 (<0.1) |
| Participants with ≥1 AEs leading to study vaccination termination | 18 (0.1) | 22 (0.1) |
| Infections and infestations | 11 (0.1) | 16 (0.1) |
| Covid-19 | 10 (0.1) | 14 (0.1) |
| Covid-19 pneumonia | 1 (<0.1) | 3 (<0.1) |
| Skin and subcutaneous tissue disorders | 4 (<0.1) | 1 (<0.1) |
| Urticaria | 3 (<0.1) | 0 |
| Angioedema | 1 (<0.1) | 0 |
| Rash | 1 (<0.1) | 0 |
| Rash erythematous | 0 | 1 (<0.1) |
| Immune system disorders | 1 (<0.1) | 0 |
| Allergy to vaccine | 1 (<0.1) | 0 |
| Injury, poisoning and procedural complications | 1 (<0.1) | 0 |
| Cervical vertebral fracture | 1 (<0.1) | 0 |
| Investigations | 1 (<0.1) | 0 |
| Myocardial necrosis marker increased | 1 (<0.1) | 0 |
| Cardiac disorders | 0 | 2 (<0.1) |
| Myocardial infarction | 0 | 1 (<0.1) |
| Ventricular extrasystoles | 0 | 1 (<0.1) |
| Gastrointestinal disorders | 0 | 1 (<0.1) |

| | | |
|---|---|---|
| Abdominal pain lower | 0 | 1 (<0.1) |
| Nervous system disorders | 0 | 2 (<0.1) |
| Migraine with aura | 0 | 1 (<0.1) |
| Trigeminal neuralgia | 0 | 1 (<0.1) |
| Social circumstances | 0 | 1 (<0.1) |
| Homicide | 0 | 1 (<0.1) |
| **Post-Booster Dose** | (N=8646) | (N=8646) |
| **AE, n (%)*** | | |
| Participants with ≥1 AEs leading to study withdrawal | 1 (<0.1) | 0 |
| Nervous system disorders | 1 (<0.1) | 0 |
| Cerebral hemorrhage | 1 (<0.1) | 0 |
| Participants with ≥1 AEs leading to study vaccination termination | 1 (<0.1) | 1 (<0.1) |
| Nervous system disorders | 1 (<0.1) | 0 |
| Cerebral hemorrhage | 1 (<0.1) | 0 |
| Musculoskeletal and connective tissue disorders | 0 | 1 (<0.1) |
| Pain in extremity | 0 | 1 (<0.1) |

*Subjects are counted only once within a phase for any given event, regardless of the number of times they actually experienced the event in that phase. AE, adverse event; Covid-19, coronavirus disease 2019.

**Table S4. Vaccine Efficacy against Covid-19 With any Documented Positive PCR with Onset at Least 14 Days after the Administration of a Booster Vaccine or Placebo (Per-Protocol Set)***

| | **At Least 14 Days after Booster Vaccination†** | | | | |
|---|---|---|---|---|---|
| | Ad26.COV2.S (N=6024) | | Placebo (N=5615) | | Vaccine Efficacy (95% CI) |
| | *no. of cases* | *person-yr* | *no. of cases* | *person-yr* | % |
| Moderate to severe–critical Covid-19‡§ | 20 | 1729.5 | 56 | 1594.9 | 67.1 (44.3-81.3)§ |
| 18-59 yr | 15 | 1386.5 | 44 | 1276.2 | 68.6 (42.5-83.8) |
| ≥60 yr | 5 | 343.0 | 12 | 318.6 | 61.3 (−18.0-89.3) |
| Symptomatic Covid-19 of any severity‖ | 20 | 1729.5 | 57 | 1594.8 | 67.6 (45.3-81.6) |
| Mild¶ | 0 | 1729.5 | 1 | 1594.8 | |
| Moderate¶ | 19 | 1729.5 | 48 | 1594.9 | 63.5 (36.7-79.7) |
| Severe–critical§‖ | 1 | 1730.4 | 8 | 1598.9 | 88.4 (−4.15-99.9)§ |
| All SARS-CoV-2 infections§‖ | 20 | 1729.5 | 57 | 1594.8 | 67.7 (44.7-80.6)§ |

*Cases of coronavirus disease 2019 (Covid-19) include all cases with a documented positive PCR, whether centrally or non-centrally confirmed, occurring in participants who were considered to be at risk for Covid-19 (seronegative at baseline [day 1] and at 14 days after booster vaccination or second placebo administration [day 71]), and negative on reverse-transcriptase–polymerase-chain-reaction [RT-PCR] testing before day 14 after booster vaccination or second placebo administration). Follow-up time was defined as the time between primary vaccination (Ad26.COV2.S or placebo) and the time of onset of the case (for participants with molecularly-confirmed Covid-19) or until the last available measurement or at the end of the double-blind phase/study discontinuation. If fewer than 6 cases were observed for an endpoint, vaccine efficacy was not determined.

†Data are from the at-risk population (defined as all participants of the Per-Protocol Set excluding participants who had a positive PCR test between day 1 and 70 and participants who discontinued prior to 14 days post-booster dose).

‡Primary endpoint (efficacy of Ad26.COV2.S in the prevention of molecularly-confirmed, moderate to severe/critical Covid-19, as compared to placebo, in SARS-CoV-2 seronegative adults, at least 14 days after the second vaccination [day 71] in the double-blind phase).

§Adjusted 95% CI, which was calculated using type I error control for multiple testing and was presented upon meeting prespecified testing conditions. The hypothesis for asymptomatic infections was not significant at the alpha level 1.25% (obtained from all SARS-CoV-2 infections). No hypothesis testing was performed for medical intervention, for which fewer than 6 cases were reported; therefore, no alpha was recycled for the hypothesis of asymptomatic infections.

‖Confirmatory secondary endpoint.

¶Supportive secondary endpoint.

CI, confidence interval; Covid-19, coronavirus disease 2019; SARS-CoV-2, severe acute respiratory syndrome coronavirus 2; yr, year.

**Table S5. Vaccine Efficacy against Molecularly-Confirmed Covid-19 between Day 15 and Day 56, by Covid-19 Severity and by Variant (Per-protocol First Dose Efficacy Set)***

| | Ad26.COV2.S (N=13,578) | | Placebo (N=13,622) | | Vaccine Efficacy (95% CI) |
|---|---|---|---|---|---|
| | *no. of cases* | *person-yr* | *no. of cases* | *person-yr* | % |
| First molecularly-confirmed moderate to severe/critical Covid-19 | 46 | 1876.1 | 137 | 1842.9 | 67.0 (53.6-76.9) |
| First molecularly-confirmed severe/critical Covid-19 | 3 | 1878.7 | 22 | 1849.3 | 86.6 (55.3-97.4) |
| First molecularly-confirmed moderate to severe/critical Covid-19 caused by | | | | | |
| Variant substitution** | 25 | 1876.1 | 73 | 1842.9 | 66.4 (46.4-79.5) |
| B.1.1.7 (Alpha) | 11 | 1876.1 | 38 | 1842.9 | 71.6 (43.2-86.9) |
| B.1.621 (Mu) | 8 | 1876.1 | 14 | 1842.9 | 43.9 (-43.4-79.6) |

*All cases of coronavirus disease 2019 (Covid-19) were centrally confirmed unless otherwise indicated and occurred in participants who were considered to be at risk for Covid-19 (seronegative at baseline and negative on reverse-transcriptase–polymerase-chain-reaction [RT-PCR] testing before day 14 after vaccine or placebo administration). **The term "variant substitution" refers to all variants combined, except for the reference strain and strains not considered variants based on their mutations. The reference strain was the Wuhan-Hu1 variant including the D614G mutation (B.1 lineage).

CI, confidence interval; Covid-19, coronavirus disease 2019; SARS-CoV-2, severe acute respiratory syndrome coronavirus 2; yr, year.

**Table S6. Summary of Solicited Local and Systemic Adverse Events by Derived Term and Worst Severity Grade (Safety Subset)**

| | Ad26.COV2.S | Placebo Double-Blind |
|---|---|---|
| **Post-dose 1** | N=3015 | N=3052 |
| **AEs, n (%)** | | |
| Participants with ≥1 local AE | 1676 (55.6) | 653 (21.4) |
| Grade 1 | 1433 (47.5) | 590 (19.3) |
| Grade 2 | 234 (7.8) | 57 (1.9) |
| Grade 3 | 9 (0.3) | 6 (0.2) |
| Vaccination Site Erythema | 263 (8.7) | 142 (4.7) |
| Grade 1 | 196 (6.5) | 118 (3.9) |
| Grade 2 | 65 (2.2) | 23 (0.8) |
| Grade 3 | 2 (0.1) | 1 (<0.1) |
| Vaccination Site Pain | 1634 (54.2) | 556 (18.2) |
| Grade 1 | 1451 (48.1) | 522 (17.1) |
| Grade 2 | 180 (6.0) | 30 (1.0) |
| Grade 3 | 3 (0.1) | 4 (0.1) |
| Vaccination Site Swelling | 167 (5.5) | 52 (1.7) |
| Grade 1 | 130 (4.3) | 42 (1.4) |
| Grade 2 | 33 (1.1) | 9 (0.3) |
| Grade 3 | 4 (0.1) | 1 (<0.1) |
| Participants with ≥1 systemic AE | | |
| Any | 1764 (58.5) | 1138 (37.3) |
| Grade 1 | 1102 (36.6) | 833 (27.3) |
| Grade 2 | 607 (20.1) | 291 (9.5) |
| Grade 3 | 55 (1.8) | 14 (0.5) |
| Fatigue | 1355 (44.9) | 760 (24.9) |
| Grade 1 | 899 (29.8) | 582 (19.1) |
| Grade 2 | 430 (14.3) | 171 (5.6) |
| Grade 3 | 26 (0.9) | 7 (0.2) |
| Headache | 1291 (42.8) | 749 (24.5) |
| Grade 1 | 919 (30.5) | 599 (19.6) |
| Grade 2 | 349 (11.6) | 145 (4.8) |
| Grade 3 | 23 (0.8) | 5 (0.2) |
| Myalgia | 1172 (38.9) | 468 (15.3) |
| Grade 1 | 843 (28.0) | 389 (12.7) |
| Grade 2 | 306 (10.1) | 75 (2.5) |
| Grade 3 | 23 (0.8) | 4 (0.1) |
| Nausea | 546 (18.1) | 316 (10.4) |
| Grade 1 | 435 (14.4) | 258 (8.5) |
| Grade 2 | 102 (3.4) | 53 (1.7) |
| Grade 3 | 9 (0.3) | 5 (0.2) |
| Pyrexia | 150 (5.0) | 14 (0.5) |
| Grade 1 | 111 (3.7) | 13 (0.4) |
| Grade 2 | 37 (1.2) | 1 (<0.1) |
| Grade 3 | 2 (0.1) | 0 |

| | Ad26.COV2.S | Placebo Double-Blind |
|---|---|---|
| **Post-Booster Dose** | N=1559 | N=1425 |
| **AE, n (%)** | | |
| Participants with ≥1 local AE | 896 (57.5) | 252 (17.7) |
| Grade 1 | 753 (48.3) | 228 (16.0) |
| Grade 2 | 133 (8.5) | 21 (1.5) |
| Grade 3 | 10 (0.6) | 3 (0.2) |
| Vaccination Site Erythema | 128 (8.2) | 56 (3.9) |
| Grade 1 | 100 (6.4) | 46 (3.2) |
| Grade 2 | 21 (1.3) | 8 (0.6) |
| Grade 3 | 7 (0.4) | 2 (0.1) |
| Vaccination Site Pain | 877 (56.3) | 225 (15.8) |
| Grade 1 | 759 (48.7) | 210 (14.7) |
| Grade 2 | 115 (7.4) | 14 (1.0) |
| Grade 3 | 3 (0.2) | 1 (0.1) |
| Vaccination Site Swelling | 88 (5.6) | 18 (1.3) |
| Grade 1 | 68 (4.4) | 13 (0.9) |
| Grade 2 | 18 (1.2) | 5 (0.4) |
| Grade 3 | 2 (0.1) | 0 |
| Participants with ≥1 systemic AE | 821 (52.7) | 442 (31.0) |
| Grade 1 | 497 (31.9) | 320 (22.5) |
| Grade 2 | 299 (19.2) | 117 (8.2) |
| Grade 3 | 25 (1.6) | 5 (0.4) |
| Fatigue | 641 (41.1) | 293 (20.6) |
| Grade 1 | 412 (26.4) | 215 (15.1) |
| Grade 2 | 215 (13.8) | 76 (5.3) |
| Grade 3 | 14 (0.9) | 2 (0.1) |
| Headache | 558 (35.8) | 270 (18.9) |
| Grade 1 | 388 (24.9) | 225 (15.8) |
| Grade 2 | 160 (10.3) | 42 (2.9) |
| Grade 3 | 10 (0.6) | 3 (0.2) |
| Myalgia | 541 (34.7) | 186 (13.1) |
| Grade 1 | 396 (25.4) | 163 (11.4) |
| Grade 2 | 136 (8.7) | 22 (1.5) |
| Grade 3 | 9 (0.6) | 1 (0.1) |
| Nausea | 225 (14.4) | 100 (7.0) |
| Grade 1 | 173 (11.1) | 81 (5.7) |
| Grade 2 | 49 (3.1) | 19 (1.3) |
| Grade 3 | 3 (0.2) | 0 |
| Pyrexia | 38 (2.4) | 4 (0.3) |
| Grade 1 | 27 (1.7) | 3 (0.2) |
| Grade 2 | 10 (0.6) | 1 (0.1) |
| Grade 3 | 1 (0.1) | 0 |

The safety subset comprised participants who recorded solicited local and systemic adverse events in an electronic diary for 7 days post-vaccination and unsolicited AEs for 28 days post-vaccination.
Participants were counted only once within the period for any given event, regardless of the number of times they experienced the event within the period. AE, adverse event.

**Table S7. Unsolicited Adverse Events of at Least Grade 3 Severity (Safety Subset)**

| | Ad26.COV2.S | Placebo |
|---|---|---|
| **Post-Dose 1** | (N=3,015) | (N=3052) |
| **AE, n (%)** | | |
| Participants with ≥1 AE of grade ≥3 | 21 (0.7) | 16 (0.5) |
| Nervous system disorders | 8 (0.3) | 3 (0.1) |
| Headache | 8 (0.3) | 3 (0.1) |
| Musculoskeletal and connective tissue disorders | 5 (0.2) | 2 (0.1) |
| Myalgia | 2 (0.1) | 1 (<0.1) |
| Osteonecrosis | 1 (<0.1) | 0 |
| Rotator cuff syndrome | 1 (<0.1) | 0 |
| Torticollis | 1 (<0.1) | 0 |
| Arthritis | 0 | 1 (<0.1) |
| Gastrointestinal disorders | 3 (0.1) | 3 (0.1) |
| Diarrhea | 2 (0.1) | 0 |
| Nausea | 2 (0.1) | 2 (0.1) |
| Esophageal obstruction | 0 | 1 (<0.1) |
| General disorders and administration site conditions | 3 (0.1) | 3 (0.1) |
| Fatigue | 2 (0.1) | 2 (0.1) |
| Injection site pain | 1 (<0.1) | 0 |
| Pyrexia | 1 (<0.1) | 0 |
| Injection site erythema | 0 | 1 (<0.1) |
| Tenderness | 0 | 1 (<0.1) |
| Vaccination site erythema | 0 | 1 (<0.1) |
| Infections and infestations | 3 (0.1) | 6 (0.2) |
| Covid-19 pneumonia | 2 (0.1) | 1 (<0.1) |
| Escherichia urinary tract infection | 1 (<0.1) | 0 |
| Covid-19 | 0 | 2 (0.1) |
| Diabetic foot infection | 0 | 1 (<0.1) |
| Gangrene | 0 | 1 (<0.1) |
| Localized infection | 0 | 2 (0.1) |
| Sepsis | 0 | 1 (<0.1) |
| Tooth abscess | 0 | 1 (<0.1) |
| Cardiac disorders | 2 (0.1) | 0 |
| Angina pectoris | 1 (<0.1) | 0 |
| Pericarditis | 1 (<0.1) | 0 |
| Injury, poisoning and procedural complications | 1 (<0.1) | 0 |
| Wrist fracture | 1 (<0.1) | 0 |
| Psychiatric disorders | 1 (<0.1) | 0 |
| Depression | 1 (<0.1) | 0 |
| Reproductive system and breast disorders | 1 (<0.1) | 0 |
| Hemorrhagic ovarian cyst | 1 (<0.1) | 0 |
| Skin and subcutaneous tissue disorders | 1 (<0.1) | 0 |

| | Ad26.COV2.S | Placebo |
|---|---|---|
| Urticaria | 1 (<0.1) | 0 |
| Metabolism and nutrition disorders | 0 | 1 (<0.1) |
| Diabetes mellitus inadequate control | 0 | 1 (<0.1) |
| Neoplasms benign, malignant and unspecified (incl cysts and polyps) | 0 | 1 (<0.1) |
| Squamous cell carcinoma of skin | 0 | 1 (<0.1) |
| Respiratory, thoracic and mediastinal disorders | 0 | 1 (<0.1) |
| Acute respiratory failure | 0 | 1 (<0.1) |
| **Post-Booster Dose** | N=1,559 | N=1,425 |
| **AE, n (%)** | | |
| Subjects with 1 or more AEs of at least grade 3 | 12 (0.8) | 7 (0.5) |
| Gastrointestinal disorders | 2 (0.1) | 0 |
| Nausea | 2 (0.1) | 0 |
| General disorders and administration site conditions | 2 (0.1) | 1 (0.1) |
| Fatigue | 1 (0.1) | 1 (0.1) |
| Vaccination site erythema | 1 (0.1) | 0 |
| Infections and infestations | 2 (0.1) | 0 |
| Acute sinusitis | 1 (0.1) | 0 |
| Localized infection | 1 (0.1) | 0 |
| Nervous system disorders | 2 (0.1) | 0 |
| Cerebrovascular accident | 1 (0.1) | 0 |
| Headache | 1 (0.1) | 0 |
| Ear and labyrinth disorders | 1 (0.1) | 0 |
| Vertigo | 1 (0.1) | 0 |
| Injury, poisoning and procedural complications | 1 (0.1) | 1 (0.1) |
| Tendon rupture | 1 (0.1) | 0 |
| Animal attack | 0 | 1 (0.1) |
| Musculoskeletal and connective tissue disorders | 1 (0.1) | 3 (0.2) |
| Myalgia | 1 (0.1) | 0 |
| Arthritis | 0 | 1 (0.1) |
| Musculoskeletal pain | 0 | 1 (0.1) |
| Pain in extremity | 0 | 1 (0.1) |
| Renal and urinary disorders | 1 (0.1) | 0 |
| Urethral hemorrhage | 1 (0.1) | 0 |
| Reproductive system and breast disorders | 1 (0.1) | 0 |
| Testicular mass | 1 (0.1) | 0 |
| Skin and subcutaneous tissue disorders | 1 (0.1) | 0 |
| Rash | 1 (0.1) | 0 |
| Cardiac disorders | 0 | 1 (0.1) |
| Myocardial infarction | 0 | 1 (0.1) |
| Myocardial ischemia | 0 | 1 (0.1) |
| Metabolism and nutrition disorders | 0 | 1 (0.1) |
| Dyslipidemia | 0 | 1 (0.1) |

| | Ad26.COV2.S | Placebo |
|---|---|---|
| Psychiatric disorders | 0 | 1 (0.1) |
| Anxiety | 0 | 1 (0.1) |

The safety subset comprised approximately 6,000 participants who recorded solicited local and systemic adverse events in an electronic diary for 7 days post-vaccination and unsolicited AEs for 28 days post-vaccination. Participants were counted only once within the period for any given event, regardless of the number of times they experienced the event within the period. Adverse events were coded using MedDRA Version 24.0.
AE, adverse event.

**Table S8. Unsolicited Adverse Events of Considered Related to Vaccination (Safety Subset)**

| | Ad26.COV2.S | Placebo |
|---|---|---|
| **Post-Dose 1** | N=3,015 | N=3052 |
| **AE, n (%)** | | |
| Participants with ≥1 related AE | 283 (9.4) | 179 (5.9) |
| General disorders and administration site conditions | 200 (6.6) | 105 (3.4) |
| Fatigue | 80 (2.7) | 73 (2.4) |
| Vaccination site pain | 63 (2.1) | 17 (0.6) |
| Injection site pain | 26 (0.9) | 7 (0.2) |
| Vaccination site erythema | 26 (0.9) | 13 (0.4) |
| Chills | 19 (0.6) | 2 (0.1) |
| Pyrexia | 10 (0.3) | 1 (<0.1) |
| Vaccination site swelling | 7 (0.2) | 2 (0.1) |
| Injection site swelling | 5 (0.2) | 0 |
| Feeling abnormal | 4 (0.1) | 1 (<0.1) |
| Injection site erythema | 4 (0.1) | 1 (<0.1) |
| Malaise | 4 (0.1) | 0 |
| Influenza like illness | 3 (0.1) | 3 (0.1) |
| Pain | 3 (0.1) | 0 |
| Injection site bruising | 1 (<0.1) | 0 |
| Injection site vesicles | 1 (<0.1) | 0 |
| Swelling | 1 (<0.1) | 0 |
| Tenderness | 0 | 1 (<0.1) |
| Nervous system disorders | 87 (2.9) | 68 (2.2) |
| Headache | 79 (2.6) | 66 (2.2) |
| Tremor | 3 (0.1) | 0 |
| Ageusia | 1 (<0.1) | 1 (<0.1) |
| Anosmia | 1 (<0.1) | 0 |
| Dizziness | 1 (<0.1) | 1 (<0.1) |
| Hypogeusia | 1 (<0.1) | 0 |
| Presyncope | 1 (<0.1) | 0 |
| Syncope | 1 (<0.1) | 0 |
| Musculoskeletal and connective tissue disorders | 76 (2.5) | 55 (1.8) |
| Myalgia | 65 (2.2) | 53 (1.7) |
| Arthralgia | 6 (0.2) | 1 (<0.1) |
| Muscular weakness | 6 (0.2) | 1 (<0.1) |
| Pain in extremity | 3 (0.1) | 1 (<0.1) |
| Musculoskeletal stiffness | 1 (<0.1) | 0 |
| Musculoskeletal pain | 0 | 1 (<0.1) |
| Gastrointestinal disorders | 34 (1.1) | 30 (1.0) |
| Nausea | 21 (0.7) | 25 (0.8) |
| Diarrhea | 10 (0.3) | 5 (0.2) |
| Abdominal pain | 4 (0.1) | 0 |

| | Ad26.COV2.S | Placebo |
|---|---|---|
| Irritable bowel syndrome | 1 (<0.1) | 0 |
| Respiratory, thoracic and mediastinal disorders | 10 (0.3) | 9 (0.3) |
| Nasal congestion | 5 (0.2) | 4 (0.1) |
| Rhinorrhea | 4 (0.1) | 5 (0.2) |
| Cough | 3 (0.1) | 2 (0.1) |
| Sneezing | 2 (0.1) | 4 (0.1) |
| Dyspnea | 1 (<0.1) | 2 (0.1) |
| Oropharyngeal pain | 1 (<0.1) | 4 (0.1) |
| Respiratory tract congestion | 1 (<0.1) | 1 (<0.1) |
| Skin and subcutaneous tissue disorders | 7 (0.2) | 2 (0.1) |
| Erythema | 2 (0.1) | 2 (0.1) |
| Cold sweat | 1 (<0.1) | 0 |
| Hyperhidrosis | 1 (<0.1) | 0 |
| Miliaria | 1 (<0.1) | 0 |
| Rash | 1 (<0.1) | 0 |
| Urticaria | 1 (<0.1) | 0 |
| Injury, poisoning and procedural complications | 4 (0.1) | 0 |
| Vaccination complication | 3 (0.1) | 0 |
| Procedural dizziness | 1 (<0.1) | 0 |
| Metabolism and nutrition disorders | 4 (0.1) | 3 (0.1) |
| Decreased appetite | 4 (0.1) | 2 (0.1) |
| Type 2 diabetes mellitus | 0 | 1 (<0.1) |
| Cardiac disorders | 2 (0.1) | 1 (<0.1) |
| Arrhythmia supraventricular | 1 (<0.1) | 0 |
| Pericarditis | 1 (<0.1) | 0 |
| Palpitations | 0 | 1 (<0.1) |
| Ear and labyrinth disorders | 1 (<0.1) | 0 |
| Vertigo | 1 (<0.1) | 0 |
| Eye disorders | 1 (<0.1) | 3 (0.1) |
| Eye discharge | 1 (<0.1) | 0 |
| Dry eye | 0 | 1 (<0.1) |
| Eye irritation | 0 | 2 (0.1) |
| Eye pruritus | 0 | 1 (<0.1) |
| Infections and infestations | 1 (<0.1) | 0 |
| Viral upper respiratory tract infection | 1 (<0.1) | 0 |
| Psychiatric disorders | 1 (<0.1) | 1 (<0.1) |
| Depressed mood | 1 (<0.1) | 0 |
| Anxiety | 0 | 1 (<0.1) |
| Vascular disorders | 1 (<0.1) | 1 (<0.1) |
| Hematoma | 1 (<0.1) | 0 |
| Peripheral coldness | 0 | 1 (<0.1) |
| Immune system disorders | 0 | 1 (<0.1) |
| Hypersensitivity | 0 | 1 (<0.1) |
| Reproductive system and breast disorders | 0 | 1 (<0.1) |
| Priapism | 0 | 1 (<0.1) |

| | Ad26.COV2.S | Placebo |
|---|---|---|
| **Post-Booster Dose** | **N=1559** | **N=1425** |
| Participants with ≥1 related AE | 79 (5.1) | 49 (3.4) |
| General disorders and administration site conditions | 52 (3.3) | 33 (2.3) |
| Fatigue | 22 (1.4) | 21 (1.5) |
| Vaccination site pain | 11 (0.7) | 4 (0.3) |
| Injection site pain | 7 (0.4) | 3 (0.2) |
| Chills | 6 (0.4) | 1 (0.1) |
| Vaccination site erythema | 6 (0.4) | 4 (0.3) |
| Vaccination site swelling | 4 (0.3) | 2 (0.1) |
| Pyrexia | 2 (0.1) | 2 (0.1) |
| Feeling abnormal | 1 (0.1) | 0 |
| Injection site swelling | 1 (0.1) | 0 |
| Malaise | 1 (0.1) | 0 |
| Pain | 1 (0.1) | 0 |
| Peripheral swelling | 1 (0.1) | 0 |
| Injection site erythema | 0 | 1 (0.1) |
| Nervous system disorders | 23 (1.5) | 17 (1.2) |
| Headache | 22 (1.4) | 17 (1.2) |
| Loss of consciousness | 1 (0.1) | 0 |
| Musculoskeletal and connective tissue disorders | 21 (1.3) | 16 (1.1) |
| Myalgia | 18 (1.2) | 16 (1.1) |
| Pain in extremity | 2 (0.1) | 0 |
| Muscular weakness | 1 (0.1) | 0 |
| Arthralgia | 0 | 1 (0.1) |
| Gastrointestinal disorders | 11 (0.7) | 3 (0.2) |
| Nausea | 8 (0.5) | 2 (0.1) |
| Abdominal pain upper | 1 (0.1) | 0 |
| Diarrhea | 1 (0.1) | 1 (0.1) |
| Vomiting | 1 (0.1) | 0 |
| Skin and subcutaneous tissue disorders | 5 (0.3) | 0 |
| Rash | 2 (0.1) | 0 |
| Hyperhidrosis | 1 (0.1) | 0 |
| Keloid scar | 1 (0.1) | 0 |
| Rash papular | 1 (0.1) | 0 |
| Respiratory, thoracic and mediastinal disorders | 3 (0.2) | 0 |
| Cough | 2 (0.1) | 0 |
| Rhinorrhea | 2 (0.1) | 0 |
| Nasal congestion | 1 (0.1) | 0 |
| Sneezing | 1 (0.1) | 0 |
| Metabolism and nutrition disorders | 1 (0.1) | 0 |
| Type 2 diabetes mellitus | 1 (0.1) | 0 |
| Psychiatric disorders | 1 (0.1) | 0 |
| Poor quality sleep | 1 (0.1) | 0 |
| Infections and infestations | 0 | 1 (0.1) |

| | Ad26.COV2.S | Placebo |
|---|---|---|
| Upper respiratory tract infection | 0 | 1 (0.1) |

The safety subset comprised approximately 6,000 participants who recorded solicited local and systemic adverse events in an electronic diary for 7 days post-vaccination and unsolicited AEs for 28 days post-vaccination. Participants were counted only once within the period for any given event, regardless of the number of times they experienced the event within the period. Adverse events were coded using MedDRA Version 24.0.
AE, adverse event.

**Table S9. Summary of SAEs Considered Related to Vaccination and Deaths During the Double-Blind Phase (Full Analysis Set)**

| | Ad26.COV2.S | Placebo |
|---|---|---|
| | N=15,705 | N=15,588 |
| **SAEs, n (%)** | | |
| Participants with ≥1 related SAE | 8 (0.1) | 3 (<0.1) |
| General disorders and administration site conditions | 2 (<0.1) | 0 |
| Injection site swelling | 1 (<0.1) | 0 |
| Pyrexia | 1 (<0.1) | 0 |
| Nervous system disorders | 2 (<0.1) | 0 |
| Cerebrovascular accident | 1 (<0.1) | 0 |
| Facial paresis | 1 (<0.1) | 0 |
| Respiratory, thoracic, and mediastinal disorders | 2 (<0.1) | 2 (<0.1) |
| Hemoptysis | 1 (<0.1) | 0 |
| Pulmonary embolism | 1 (<0.1) | 1 (<0.1) |
| Respiratory distress | 0 | 1 (<0.1) |
| Cardiac disorders | 1 (<0.1) | 0 |
| Pericarditis | 1 (<0.1) | 0 |
| Ear and labyrinth disorders | 1 (<0.1) | 0 |
| Vertigo | 1 (<0.1) | 0 |
| Immune system disorders | 1 (<0.1) | 0 |
| Allergy to vaccine | 1 (<0.1) | 0 |
| Investigations | 1 (<0.1) | 0 |
| Increased myocardial necrosis marker | 1 (<0.1) | 0 |
| Gastrointestinal disorders | 0 | 1 (<0.1) |
| Acute pancreatitis | 0 | 1 (<0.1) |
| **Deaths** | | |
| Deaths | 4 | 13 |
| Covid-19 related deaths* | 0 | 1 |

*With onset ≥14 days after booster.

Covid-19, coronavirus disease 2019; SAE, serious adverse event.

**Table S10. Summary of Adverse Events of Clinical Interest and Special Interest During the Double-Blind Phase (Full Analysis Set)**

| | Post-Dose 1 | | Post-Booster Dose | |
|---|---|---|---|---|
| | **Ad26.COV2.S** N=15,705 | **Placebo** N=15,588 | **Ad26.COV2.S** N=8,646 | **Placebo** N=8,043 |
| **AEs of clinical interest, n (%)*** | | | | |
| Embolic and thrombotic events, any | 2 (<0.1) | 6 (<0.1) | 3 (<0.1) | 3 (<0.1) |
| Embolic and thrombotic events, arterial | 1 (<0.1) | 4 (<0.1) | 0 | 2 (<0.1) |
| Acute myocardial infarction | 1 (<0.1) | 0 | 0 | 0 |
| Myocardial infarction | 0 | 2 (<0.1) | 0 | 2 (<0.1) |
| Transient ischemic attack | 0 | 2 (<0.1) | 0 | 0 |
| Embolic and thrombotic events, venous | 0 | 2 (<0.1) | 1 (<0.1) | 1 (<0.1) |
| Pulmonary embolism | 0 | 1 (<0.1) | 1 (<0.1) | 0 |
| Deep vein thrombosis | 0 | 1 (<0.1) | 0 | 1 (<0.1) |
| Embolic and thrombotic events, vessel type unspecified and mixed arterial and venous | 1 (<0.1) | 0 | 2 (<0.1) | 0 |
| Cerebrovascular accident | 0 | 0 | 1 (<0.1) | 0 |
| Hemiparesis | 0 | 0 | 1 (<0.1) | 0 |
| Embolism | 1 (<0.1) | 0 | 0 | 0 |
| Hypersensitivity | 56 (0.36) | 35 (0.22) | 15 (0.17) | 9 (0.11) |
| Urticaria | 11 (<0.1) | 3 (<0.1) | 3 (<0.1) | 1 (<0.1) |
| Hemorrhagic disorders† | 24 (0.15) | 14 (<0.1) | 17 (0.2) | 7 (<0.01) |
| Arthritis | 24 (0.15) | 12 (<0.1) | 4 (<0.1) | 5 (<0.1) |
| Tinnitus | 4 (<0.1) | 2 (<0.1) | 2 (<0.1) | 2 (<0.1) |
| Thrombocytopenia | 1 (<0.1) | 1 (<0.1) | 1 (<0.1) | 1 (<0.1) |
| Facial Paralysis | 1 (<0.1) | 2 (<0.1) | 1 (<0.1) | 0 |
| Bell's Palsy | 1 (<0.1) | 0 | 0 | 0 |
| Pericarditis | 1 (<0.1) | 0 | 0 | 0 |

| | Post-Dose 1 | | Post-Booster Dose | |
|---|---|---|---|---|
| | **Ad26.COV2.S** N=15,705 | **Placebo** N=15,588 | **Ad26.COV2.S** N=8,646 | **Placebo** N=8,043 |
| Convulsions/seizures | 0 | 0 | 0 | 1 (<0.1) |
| **Adverse event of special interest** | | | | |
| Thrombosis with thrombocytopenia | 0 | 0 | 0 | 0 |

Post-dose 1=day 1 to day 29; Post-booster=day 57 to day 85.
*Other than events related to trauma, injury, or injection site AEs, no imbalances were seen for any system organ class level within 28 days after any vaccination.
†Four AEs were considered SAEs (1 in the vaccine group of worsening of hemorrhagic ovarian cyst versus 1 in the placebo group of lower gastrointestinal bleed after dose 1, and 2 of urethral bleeding and cerebral hemorrhage in the vaccine group versus 0 in the placebo group post-booster).

AE, adverse event; SMQ, Standardized MedDRA (Medical dictionary for regulatory activities) query.
